# Accurate phenotypic classification and exome sequencing allow identification of novel genes and variants associated with adult-onset hearing loss

**DOI:** 10.1101/2023.04.27.23289040

**Authors:** Morag A. Lewis, Jennifer Schulte, Lois Matthews, Kenneth I. Vaden, Claire J. Steves, Frances M.K. Williams, Bradley A. Schulte, Judy R. Dubno, Karen P. Steel

## Abstract

Adult-onset progressive hearing loss is a common, complex disease with a strong genetic component. Although to date over 150 genes have been identified as contributing to human hearing loss, many more remain to be discovered, as does most of the underlying genetic diversity. Many different variants have been found to underlie adult-onset hearing loss, but they tend to be rare variants with a high impact upon the gene product. It is likely that combinations of more common, lower impact variants also play a role in the prevalence of the disease.

Here we present our exome study of hearing loss in a cohort of 532 older adult volunteers with extensive phenotypic data, including 99 older adults with normal hearing, an important control set. Firstly, we carried out an outlier analysis to identify genes with a high variant load in older adults with hearing loss compared to those with normal hearing. Secondly, we used audiometric threshold data to identify individual variants which appear to contribute to different threshold values. We followed up these analyses in a second cohort. Using these approaches, we identified genes and variants linked to better hearing as well as those linked to worse hearing.

These analyses identified some known deafness genes, demonstrating proof of principle of our approach. However, most of the candidate genes are novel associations with hearing loss. While the results support the suggestion that genes responsible for severe deafness may also be involved in milder hearing loss, they also suggest that there are many more genes involved in hearing which remain to be identified. Our candidate gene lists may provide useful starting points for improved diagnosis and drug development.

## Introduction

Hearing loss is a common, complex condition with a strong genetic component. More than 700 genes have been found to underlie Mendelian hearing loss in humans and/or mice (reviewed in (Lewis et al. 2022)), but large-scale mouse studies suggest there may be as many as 1000 genes which alone can result in hearing impairment when mutated (Ingham et al. 2019). Identifying the genes and specific gene variants involved in age-related hearing loss may suggest genes or pathways that can be targeted therapeutically, as well as being useful for diagnosis.

Identifying genes and variants involved in hearing loss is challenging owing to the heterogeneity of the disease. The inner ear is a complex system, with multiple molecular components that need to function and interact correctly to enable normal hearing. Family studies have led to the identification of many variants involved in adult-onset Mendelian hearing loss (for example, *MIR96* (Mencia et al. 2009), *DMXL2* (Chen et al. 2017), reviewed in (Ahmadmehrabi et al. 2021)), but these tend to be very rare or even private variants, and are unlikely to explain all of the hearing loss seen in humans. Some loci have been identified through genome-wide association studies (GWAS) (Ivarsdottir et al. 2021; Kalra et al. 2020; Wells et al. 2019), but very large numbers of people are needed and GWAS chips are limited by their use of common, ancient variants. Whole exome and genome sequencing offer greater scope for identifying causative variants whatever their allele frequency and, indeed, recent studies using exome sequencing (Lewis et al. 2022; Praveen et al. 2022) suggest that intermediate frequency variants also play a role in hearing difficulty.

Another challenge in this field is the complexity of auditory phenotypes. Hearing loss does not have a single pathogenic mechanism, but can result from multiple inner ear pathologies. At present, with few exceptions, accurate diagnosis of the underlying hearing problem is not possible. In addition, many large-scale studies make use of self-reported questionnaires to explore hearing impairment. Although self-reported hearing difficulty is fairly well correlated with overall audiometric thresholds (Cherny et al. 2020; Davis and Research 1995; Nondahl et al. 1998), and hearing aid prescription is a surrogate for abnormal pure tone audiometry at least in the UK, these may be prone to subjective bias and offer no way to distinguish between different auditory phenotypes and underlying pathologies. Large cohorts with good audiometric phenotyping offer more objective classification of participants, which may allow more sensitive detection of causal genes and variants. This has been demonstrated by a recent study on such a cohort, which found an increased burden of predicted deleterious rare variants in known hearing loss genes in people with sensorineural hearing loss compared to controls with good hearing (assessed by pure-tone threshold) or no medical reports of hearing loss (Ahmadmehrabi et al. 2022). Furthermore, appropriate quality control is vital for genetic studies, and because adult-onset hearing loss is so common, a well-characterised age-matched group with audiometrically determined normal hearing provides a better control than volunteers reporting no hearing difficulty or younger adults with normal hearing.

Here we present our data from a cohort of older adult volunteers having extensive phenotype data, including 99 older adults with good hearing. We have carried out both gene-based and variant-based tests to identify candidate genes and variants, and prioritised those candidates using a variety of methods, including repeat analyses in a second, smaller cohort.

## Methods

### Ethics

All human subjects research was conducted in accordance with the Declaration of Helsinki. Informed consent was obtained in this study, which was approved by the Medical University of South Carolina (MUSC) Institutional Review Board (for the MUSC cohort) and Guys & St Thomas’ Trust (GSTT) Ethics Committee (for the TwinsUK cohort).

### Participants and audiometric measurements

The primary cohort consisted of 532 volunteers enrolled in an ongoing longitudinal study of age-related hearing loss at MUSC, dating from 1987, described in detail in Dubno et al, 2013 (Dubno et al. 2013). Notably, no participants exhibited any sign of conductive hearing loss or active otologic disease. The 532 individuals were aged 55 years or older. Pure tone thresholds (at 0.25, 0.5, 1.0, 2.0, 3.0, 4.0, 6.0 and 8.0 kHz) were obtained for each ear of each person, along with questionnaire responses concerning noise exposure history.

For the follow-up cohort, we selected 159 participants from the TwinsUK study based on age (55 years and older), self-reported ethnicity (“White”), and availability of both exome and pure-tone audiometry data. The pure-tone audiometry data collection has previously been described (Wolber et al. 2012); briefly, all participants underwent an otologic examination followed by an air-conduction pure-tone audiogram for each ear (0.125, 0.25, 0.5, 1.0, 2.0, 4.0, 6.0 and 8.0 kHz). Participants also answered a detailed questionnaire concerning medical history and environmental exposure to factors relevant to hearing.

### Classification of audiograms

Phenotype cohorts were formed based on selection criteria to define individuals with representative metabolic or sensory hearing losses, as well as normal hearing, to enable comparisons between specific phenotypes. Audiograms were classified into one of three main categories (Older-Normal, Metabolic, and Sensory) based on the estimated metabolic and sensory components of the observed hearing loss (Vaden et al. 2022). The typical audiogram in metabolic cases shows mildly elevated thresholds at low frequencies sloping gently downwards towards higher frequencies, while the shape of a typical sensory pattern has normal thresholds at low frequencies and steeply downwards-sloping thresholds at high frequencies (Dubno et al. 2013; Schmiedt 2010; Vaden et al. 2017). These typical profiles (obtained from 402 older adult audiograms (Vaden et al. 2022)) can be used to approximate the metabolic and sensory components of the hearing loss observed in an individual ear. It is then possible to calculate the contribution of each profile to this approximation, and the quality of the approximation itself is represented by the line-fit error.

To classify these cohorts, first, metabolic and sensory estimates and line-fit error were calculated for each of the right/left pairs of audiograms. Second, poorly fit audiograms were excluded from classification using the criterion of line-fit error ≥15 dB, which identifies audiograms with configurations inconsistent with age-related hearing loss (e.g., corner audiograms). These rejected audiograms are referred to as “Unselected” below. Third, a set of simple rules (below) using the metabolic and sensory estimates were applied to classify cases into the four remaining categories.

Using this approach, the Older-Normal category was defined by cases with summed metabolic + sensory estimates that were <20 dB HL, with <10 dB difference in the estimates between ears. The Metabolic category was selected from the remaining cases (i.e., not Older-Normal) with metabolic estimates ≥20 dB, ear asymmetries in the metabolic estimate ≤15 dB, sensory estimates <20 dB, and metabolic > sensory estimates. The Sensory category was selected from the remaining cases (i.e., not Older-Normal and not Metabolic) with sensory estimates ≥15 dB, ear asymmetries in the sensory estimate ≤20 dB, metabolic estimates <25 dB, and sensory > metabolic estimates. Finally, the remaining cases (i.e., not Older-Normal, not Metabolic, not Sensory) were less clearly representative of metabolic or sensory hearing loss, and were labelled Unclassified. After completing the rule-based selection, all audiograms in each category were reviewed by eye; a few anomalous cases were removed and a few cases were added based on consistency with a category. There were a total of 1-10 manual additions or removals for each category (Suppl. Figures 1,2).

### Exome sequencing and alignment

Libraries for exome sequencing of the MUSC cohort were prepared using the Agilent SureSelect X2 Target Enrichment System (version 5) and the Agilent SureSelect Human All Exon V5 kit, which included 5’ and 3’ UTRs. DNA was sheared using the Covaris S220 focused ultrasonicator. Libraries were sequenced on the Illumina HiSeq 2500.

The exome sequencing of the Twins UK cohort has been previously described (Williams et al. 2012). Briefly, DNA extracted from whole blood was hybridised to NimbleGen human exome arrays and sequenced using Illumina sequencing machines (NimbleGen 2.1M and the Illumina GAIIx for the first batch of sequencing, and NimbleGen EZ v2 and the HiSeq 2000 for the second).

For both cohorts, fastq files were aligned to GRCh38 using Hisat2.0 (Kim, Langmead, and Salzberg 2015), following quality control steps (Suppl. Table 1). Bam files were realigned to sex-corrected genomes using XYalign (Webster et al. 2019).

### Variant calling, filtering, annotation and confirmation

After read alignment, genomic variants were called using three callers; GATK HaplotypeCaller (McKenna et al. 2010; Poplin et al. 2018), BCFtools (Danecek et al. 2021) and Freebayes (Garrison 2012) (Suppl. Table 1). Combining calls from multiple callers has been shown to offer more accurate variant calling (Bao et al. 2014). HaplotypeCaller quality scores were recalibrated using the GATK Variant Quality Score Recalibrator (VQSR) tool (Van der Auwera et al. 2013), which annotates variants into tranches which represent subsequent levels of sensitivity versus specificity. Variants in the highest tranche are very high accuracy, very likely to be true but also incomplete (high specificity, lower sensitivity). The second tranche is less specific but more sensitive, and so on (https://gatk.broadinstitute.org/hc/en-us/sections/360007226651-Best-Practices-Workflows). BCFtools calls were filtered using vcfutils (Danecek et al. 2021), and Freebayes calls were filtered using vcftools (Danecek et al. 2011) (Suppl. Table 1).

After quality filtering each set of calls in the MUSC cohort, a selection of variants representing a range of quality scores was tested by Sanger sequencing to ascertain the best combination of filters. The resequenced variants were assessed based on whether the variant was correctly identified and also whether the sample genotypes were correctly called. We obtained 184 sequences from 66 variants, and the most accurate variant calls were those which had passed the GATK VQSR filters and had also passed at least one of the BCFtools or Freebayes filters. Those variants from the second GATK VQSR tranche which had passed the BCFtools filter were also found to be accurately called. The variant calls from all three callers were combined according to these requirements, and this combination filter was implemented for both the MUSC and the TwinsUK cohorts (Supp Table 2). Where there was a genotype disagreement (eg GATK and Freebayes called 0/1 and BCFtools called 1/1), the majority call was accepted. Calls like this, and calls with no disagreement, accounted for 99.8% (MUSC) and 85.5% (TwinsUK) of total calls. Where there were three different calls, one for homozygote alternate, one for homozygote reference and one for heterozygote, a heterozygous genotype was assigned (0.00025% of calls (MUSC); 0.00046% of calls (TwinsUK)). Other call combinations were considered missing (0.19% (MUSC); 14.5% (TwinsUK)). The reason for the TwinsUK sequencing having a higher call missing rate is due to the exome sequencing having been processed in two batches with different exome arrays (Williams et al. 2012), so some variants have only been called in half the participants.

941,165 (MUSC) and 281,261 (TwinsUK) variants passed these quality filters, and were then tested for excess heterozygosity using the R HardyWeinberg package (Graffelman 2015; Graffelman and Camarena 2008) to identify and remove misaligned variants (Fuentes Fajardo et al. 2012). Also excluded were variants which had a high allele frequency in their cohort (defined as variants with cohort allele frequency above minor allele frequency (MAF)+0.4), which are likely to be aligner miscalls in low-complexity regions (Maffucci et al. 2019). From this variant calling and quality filtering pipeline (Suppl. Figure 3, Suppl. Table 2) 938,008 (MUSC) and 279,434 (TwinsUK) high quality variants were obtained. Further Sanger sequencing was carried out on 113 variants in multiple samples from the MUSC cohort, and individual call accuracy was 94.7% (357 correct from 377 total). Only two variants were not validated; the remainder of the incorrect calls were errors in zygosity (eg a heterozygote call for an individual homozygous for the alternate allele).

Mitochondrial variants were called using GATK Mutect2 (Benjamin et al. 2019) and filtered using GATK FilterMutectCalls. Although none of the kits used (Agilent SureSelect All Exon v5, NimbleGen 2.1M and NimbleGen EZv2) include the mitochondrial chromosome, off-target reads have been found to map correctly (Picardi and Pesole 2012). Griffin et al (Griffin et al. 2014) tested this using three different exome kits (including the Agilent SureSelect Human All Exon 50Mb kit and the NimbleGen SeqCap EZ Exome Library v2.0) and conventional mitochondrial DNA sequence, and found that if the coverage was high enough (>30x), heteroplasmy over 40% could be reliably detected. The mitochondrial calls were therefore further filtered by read depth and variant allele fraction (Suppl. Table 2), resulting in 1174 (MUSC) and 142 (TwinsUK) variants, most of which were homoplasmic (with a variant allele fraction > 0.95). Mitochondrial variants were then annotated and filtered as the genomic variants were (Suppl. Figure 3), resulting in 226 (MUSC) and 16 (TwinsUK) high impact variants with MAF < 0.1 (Suppl. Figure 3). For the two analyses carried out (described below), homoplasmic variants were treated as homozygote calls and heteroplasmic variants as heterozygote calls.

Genomic and mitochondrial variants were annotated using the Ensembl Variant Effect Predictor (VEP) v100 (McLaren et al. 2016). Annotation sources included 5’UTR variant prediction (Sutr, (Pajusalu)), splice site variant prediction (SpliceAI, (Jaganathan et al. 2019)), pathogenicity prediction (CADD, (Rentzsch et al. 2019)) and minor allele frequency (gnomAD, TOPMED, ESP6500 and 1000Genomes (Fu et al. 2013; Genomes Project et al. 2015; Karczewski et al. 2020; Taliun et al. 2021)). Variants were filtered for high predicted impact and MAF < 0.1, based on our previous work (Suppl. Table 2, (Lewis et al. 2022)), resulting in 29,807 (MUSC) and 21,432 (Twins UK) high quality, high impact variants (Suppl. Figure 3).

Chosen variants from the MUSC cohort were resequenced using Sanger sequencing (carried out by Eurofins Genomics LLC, Kentucky, USA). Primers for Sanger sequencing were designed using primer3 (Untergasser et al. 2012), and sequence traces were checked using Gap4 (Bonfield, Smith, and Staden 1995).

### Regression analysis of number of variants per gene

Four comparisons were carried out: Older-Normal hearing to all hearing loss (including Unselected and Unclassified participants); Older-Normal hearing to Metabolic hearing loss; Older-Normal hearing to Sensory hearing loss; and Metabolic hearing loss to Sensory hearing loss. 12176 genes (including mitochondrial genes) had at least one variant called in one sample, and were assessed in each analysis. For each comparison, a linear regression was carried out on the total number of variants per gene per group. In the first three comparisons, the number of variants in the Older-Normal hearing group was used to predict the expected number of variants in the hearing loss group, and in the fourth comparison, the number of variants in the Metabolic hearing loss group was used to predict the expected number of variants in the Sensory hearing loss group. The residuals (the difference between the observed and predicted variant load for each gene) were used to determine the outlier genes. Briefly, the first (Q1) and third (Q3) quartile and the interquartile distance D (Q3-Q1) were calculated for each regression’s residuals, and outlier genes were defined as those with residuals > Q3 + 6D and those with residuals < Q1 – 6D (Vuckovic et al. 2018). Hypergeometric tests for enrichment were carried out using R.

### Compilation of the list of known deafness genes

The list of deafness genes consists of those genes known to underlie hearing impairment in humans or in mice, and was manually compiled and curated from the literature. It includes all the genes listed in the Hereditary Hearing Loss Homepage (hereditaryhearingloss.org/) and genes which, when mutated, result in altered hearing thresholds in mutant mice, as reported by the International Mouse Phenotyping Consortium (www.mousephenotype.org) (average thresholds were individually checked for shifts > 10dB and low variance between individuals). This list is an update of that reported in our previous study (Lewis et al. 2022); it consists of 515 genes linked to hearing impairment in mice, 72 genes linked to hearing impairment in humans, and 122 genes linked to hearing impairment in both mice and humans (Figure 1, Suppl. Table 3).

**Figure 1.**
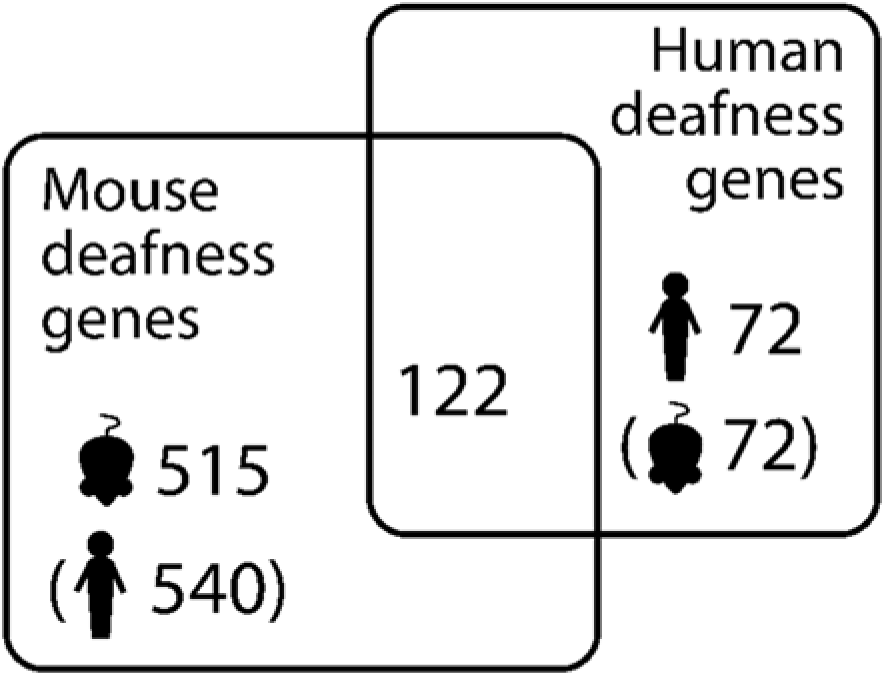
Numbers of known deafness genes in humans and mice. Brackets indicate orthologues (e.g. there are 540 human orthologues of the 515 mouse deafness genes).

**Figure 2.**
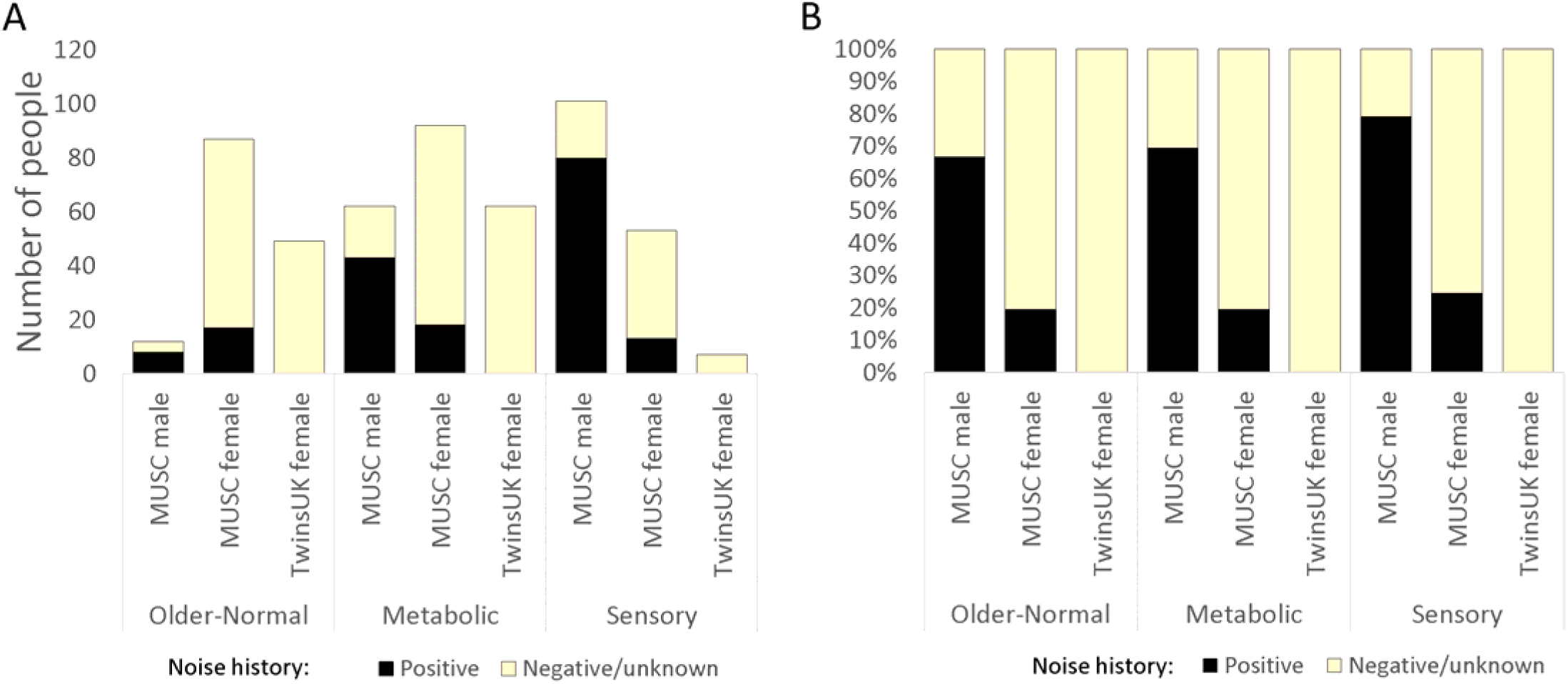
Bar charts showing the numbers of participants in each classification (not including Unclassified or Unselected cases) in the MUSC and TwinsUK cohorts (with twins removed from the latter). Black sections represent those participants reporting a positive noise history. A shows the numbers, and B shows the percentages reporting a positive noise history within each category.

### Expression analysis of outlier genes

Gene expression in the mouse inner ear was assessed using single cell RNAseq data obtained from the gEAR portal (https://umgear.org (Orvis et al. 2021)). Datasets were chosen to include multiple ages (embryonic day (E)16, postnatal day (P)1, P7 (Kolla et al. 2020), P15 (Ranum et al. 2019), P20 (Xue et al. 2021) and P30 (Korrapati et al. 2019)), and expression was normalised within each dataset and cell type to *Hprt* expression. Where a dataset had more than one set of measurements for a cell type (eg the E16 dataset has “OHC_1” and “OHC_2”, both representing outer hair cells), expression levels were averaged. The expression of each gene was plotted in 12 different cell types, as defined by the original experiments. Eleven marker genes were plotted for comparison (hair cells: *Myo7a*; inner hair cells: *Fgf8*; outer hair cells: *Slc26a5*; non-sensory cells: *Sox2*; inner pillar cells: *S100b*; Deiters’ cells: *Hes5*; marginal cells: *Kcne1*; intermediate cells: *Met*; basal cells: *Cldn11*; spindle and root cells: *Slc26a4*; fibrocytes: *Gm525*). These are known marker genes for their cell types, with the exception of *Gm525*, which was chosen based on its fibrocyte-specific expression at P30 (Korrapati et al. 2019).

### Threshold difference detection

To assess each individual variant, audiograms were plotted with participants separated into groups by genotype and sex. Variants with fewer than 5 people/group in all alternate allele groups were excluded. Each stimulus frequency was tested for a difference of 20 dB or more in average thresholds, and a maximum limit was imposed on standard deviation in the alternate allele group which differed by stimulus frequency (15 dB for 0.125-0.5 kHz, 20 dB for 1-2 kHz, 25 dB for 3-4 kHz, 30 dB for over 4 kHz) to prioritise variants associated with consistent threshold patterns. All variants where at least two stimulus frequencies in each ear passed this filter were put through to permutation testing. Permutations (20,000) were then carried out, with individuals from the cohort assigned randomly to groups of the same number and sex, to assess the likelihood that those stimulus frequencies passing the filter were observed by chance. If more than 1000 random shufflings produced a similar result, the variant was rejected. This was carried out automatically, and the scripts can be found at github.com/moraglewis/ThreADD.

## Results

### MUSC cohort classification

Our primary cohort consisted of 532 participants; 292 female and 240 male participants, with an overall average age of 72.25 years (71.96 years for women, 72.60 for men). 62 women and 182 men reported a positive noise history (Table 1, Figure 2). 99 participants were classified into the Older-Normal audiogram category; 87 women and 12 men. In the Metabolic category, there were 92 women and 62 men, while in the Sensory category, there were 53 women and 101 men. PLINK v2 (Chang et al. 2015) was used to check for relatedness using common variants in linkage equilibrium; none of the cohort were related. We used the 2504 individuals from the 1000 Genomes study (Genomes Project et al. 2015) to plot out the ancestry of this cohort and found it to be largely non-Finnish European, most similar to the “British in England and Scotland” and “Utah residents with Northern and Western European ancestry” sub-populations (Suppl. Figure 4).

**Table 1.** Details of the cohorts used in this study.

|  | *Entire MUSC cohort* |  | *Older-Normal* |  | *Metabolic* |  | *Sensory* |  | *Unclassified* |  | *Unselected* |  |
| --- | --- | --- | --- | --- | --- | --- | --- | --- | --- | --- | --- | --- |
|  | Male | Female | Male | Female | Male | Female | Male | Female | Male | Female | Male | Female |
| **Number** | 240 | 292 | 12 | 87 | 62 | 92 | 101 | 53 | 48 | 47 | 17 | 13 |
| **Average age** | 72.6 | 71.96 | 66.46 | 67.71 | 74.51 | 74.29 | 72.32 | 73.39 | 72.36 | 73.57 | 72.27 | 71.64 |
| **Positive noise history** | 189 | 62 | 8 | 17 | 43 | 18 | 80 | 13 | 42 | 9 | 14 | 5 |
| **Unknown noise history** |  |  |  |  | 1 |  | 1 | 1 |  |  |  |  |
|  | *Entire TwinsUK cohort* |  | *Older-Normal* |  | *Metabolic* |  | *Sensory* |  | *Unclassified* |  | *Unselected* |  |
|  | All | Twins removed | All | Twins removed | All | Twins removed | All | Twins removed | All | Twins removed | All | Twins removed |
| **Number** | 159 | 149 | 53 | 49 | 66 | 62 | 7 | 7 | 19 | 17 | 14 | 13 |
| **Average age** | 64.82 | 64.69 | 61.3 | 61.37 | 66.48 | 66.26 | 68.14 | 68.14 | 67.32 | 66.65 | 65.29 | 65.15 |

**Table 2.** The number of genes, known deafness genes and highly variable genes in the high variant load lists from the outlier regression analyses comparing different phenotypes in the MUSC and TwinsUK cohorts. Red outlines indicate significant enrichment of deafness or highly variable genes in the high variant load list. Genes are listed in Supp Tables 4, 5.

| Cohort | Participants | Comparison | Variant load in metabolic hearing loss |  |  | Variant load in Sensory hearing loss |  |  |
| --- | --- | --- | --- | --- | --- | --- | --- | --- |
|  |  |  | Genes | Deafness genes | Variable genes | Genes | Deafness genes | Variable genes |
| MUSC | All | Older-Normal vs all others | 31 | 0 (adj.p=1) | 2 (adj.p=0.40) | 96 | 7 (adj.p=0.31) | 16 (adj.p=2.95x10<sup>-6</sup>) |
| MUSC | All | Older-Normal vs Metabolic | 10 | 0 (adj.p=1) | 0 (adj.p=1) | 40 | 7 (adj.p=0.020) | 6 (adj.p=0.0068) |
| MUSC | All | Older-Normal vs Sensory | 7 | 0 (adj.p=1) | 0 (adj.p=1) | 58 | 4 (adj.p=0.49) | 10 (adj.p=0.00018) |
| MUSC | Female | Older-Normal vs all others | 35 | 0 (adj.p=1) | 2 (adj.p=0.44) | 107 | 9 (adj.p=0.20) | 18 (adj.p=9.32x10<sup>-7</sup>) |
| MUSC | Female | Older-Normal vs Metabolic | 4 | 0 (adj.p=1) | 1 (adj.p=0.23) | 16 | 2 (adj.p=0.36) | 5 (adj.p=0.00070) |
| MUSC | Female | Older-Normal vs Sensory | 6 | 0 (adj.p=1) | 2 (adj.p=0.036) | 51 | 5 (adj.p=0.26) | 7 (adj.p=0.0060) |
| TwinsUK | Female | Older-Normal vs all others | 147 | 4 (p=0.86) | 10 (p=0.046) | 269 | 17 (p=0.053) | 32 (p=4.27x10<sup>-9</sup>) |
| TwinsUK | Female | Older-Normal vs Metabolic | 23 | 1 (p=0.62) | 1 (p=0.58) | 55 | 3 (p=0.40) | 5 (p=0.052) |
|  |  |  | Variant load in metabolic hearing loss |  |  | Variant load in Sensory hearing loss |  |  |
|  |  |  | Genes | Deafness genes | Variable genes | Genes | Deafness genes | Variable genes |
| MUSC | All | Metabolic vs Sensory | 13 | 2 (adj.p=0.36) | 1 (adj.p=0.43) | 34 | 0 (adj.p=1) | 6 (adj.p=0.0038) |
| MUSC | Male | Metabolic vs Sensory | 23 | 5 (adj.p=0.019) | 2 (adj.p=0.28) | 89 | 5 (adj.p=0.62) | 15 (adj.p=3.96x10<sup>-6</sup>) |
| MUSC | Female | Metabolic vs Sensory | 10 | 1 (adj.p=0.62) | 2 (adj.p=0.088) | 40 | 1 (adj.p=1) | 3 (adj.p=0.27) |

### TwinsUK cohort classification

There were 159 female participants from the TwinsUK cohort meeting our requirements, including ten dizygotic twin pairs. There were no monozygotic twin pairs, and no other relatedness was reported. The overall mean age was 64.82 years. There were few positive responses to questions about noise exposure in work or leisure activities, so no participants were classified as having a positive noise history. We carried out the same ancestry analysis on these 159 participants, and found they also had a non-Finnish European ancestry, and like the MUSC cohort, they were most similar to the “British in England and Scotland” and “Utah residents with Northern and Western European ancestry” sub-populations from the 1000 Genomes (Suppl. Figure 4).

One twin of each pair was removed from the cohort; where twins were classified into the same category, the removed twin was chosen at random (6 pairs). For 3 pairs, one twin was classified as Older-Normal or Metabolic, with the other either Unclassified or Unselected; in those cases, the Unclassified or Unselected twin was removed. The last pair consisted of one twin classified as Older-Normal and one classified as Metabolic; both were removed for the outlier analysis but for the threshold analysis, one (the Older-Normal-classified twin) was chosen at random for removal. After twin removal, there were 49 in the Older-Normal category (average age 61.37 years), 62 in the Metabolic category (average age 66.26 years) and 7 in the Sensory category (average age 68.14 years) (Table 1, Figure 2).

### Outlier analysis

To investigate variant load in hearing loss, and in and between the specific phenotypes, the number of variants per gene in participants belonging to one group (eg Older-Normal) were compared to the number of variants in the same gene in participants belonging to another group (eg Metabolic). The participants were also compared segregated by sex, because the genetic contribution to adult-onset hearing loss differs by sex (Lewis et al. 2022); however, because there were only 12 men classified as having Older-Normal hearing, comparisons which required that group were not carried out, resulting in 9 comparisons from the MUSC cohort (Figure 3,A-I; Table 2). Two lists of genes were obtained from each comparison; one with an exceptionally high variant load in the first group and one with an exceptionally high variant load in the second group (Suppl. Tables 4, 5).

**Figure 3.**
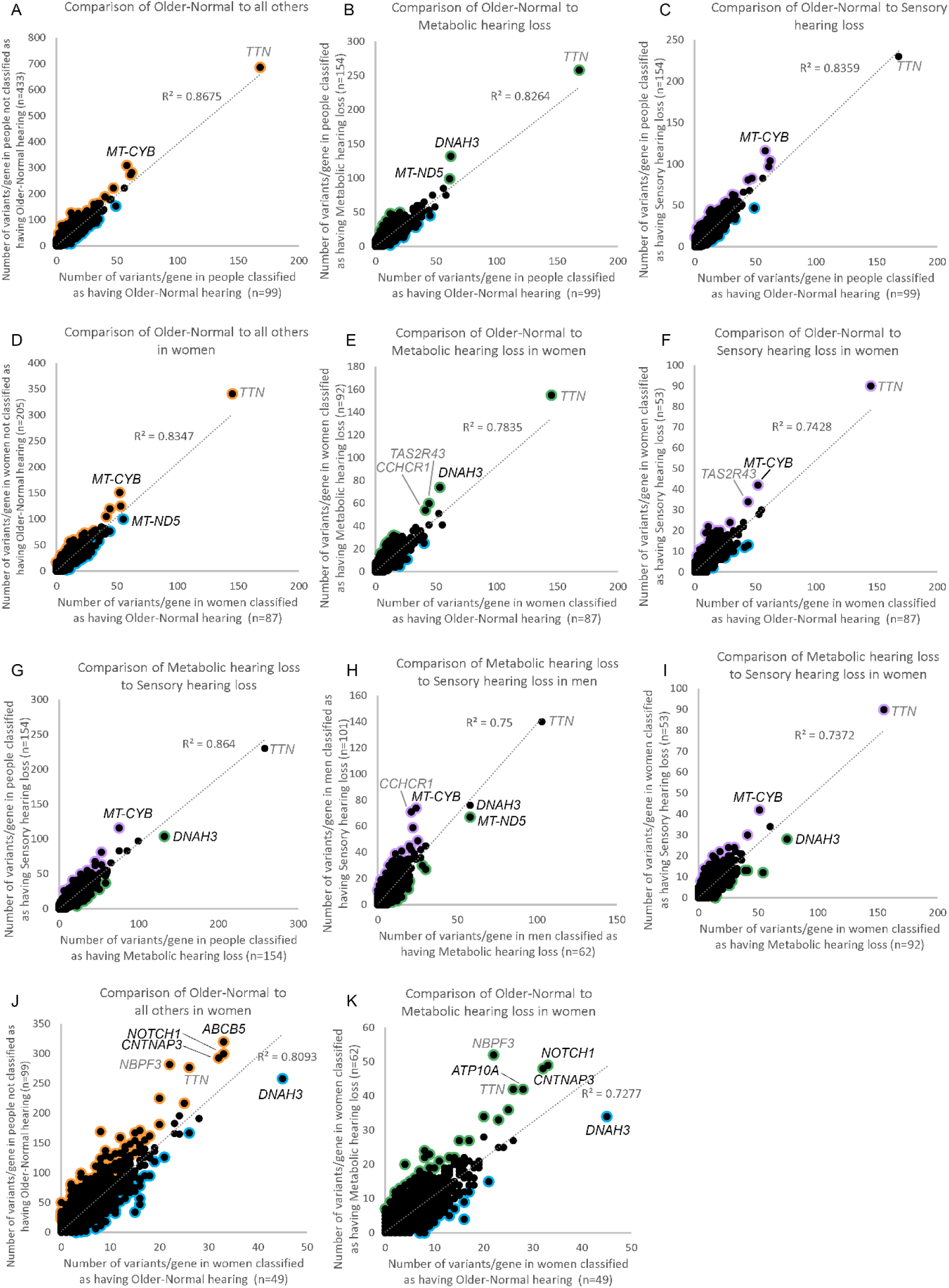
Comparison of variant load per gene between different classifications. Each point represents a gene. Outliers (Supp Tables 4, 5) are marked in orange (for higher load in participants not classified as Older-Normal), blue (for higher load in participants classified as having Older-Normal hearing), purple (for higher load in participants classified as having Sensory hearing loss) or green (for higher load in participants classified as having Metabolic hearing loss). A-I show comparisons in the MUSC cohort; A,B,C,G show all participants, D,E,F,I show female participants and H shows male participants. A,D show a comparison of variant load in people in the Older-Normal category to all others in the cohort, B,E show a comparison of variant load in people in the Older-Normal category to people in the Metabolic category, C,F show a comparison of variant load in people in the Older-Normal category to people in the Sensory category, and G,H,I show a comparison of variant load in people in the Metabolic category to people in the Sensory category. J, K show comparison of variant load in the TwinsUK cohort (which is all female); J shows a comparison of variant load in people in the Older-Normal category to all others in the cohort and K shows a comparison of variant load in people in the Older-Normal category to people in the Metabolic category. Genes with a lot of variants in, at the top right of each plot, are labelled; in some cases these are highly variable genes (*TTN*, *CCHCR1*, *NBPF3*, *TAS2R43*, shown in grey).

To investigate these gene lists, findings were compared to a list of 734 genes which are known to underlie hearing loss in humans and/or mice (Suppl. Table 3; this includes 540 human orthologues of the 515 deafness genes known only from mouse studies (Figure 1)). These are good candidates for adult-onset hearing loss, and we suggest that enrichment in these genes supports the relevance to hearing loss. Only two lists showed a significant enrichment for hearing genes; the list of genes with high variant load in Metabolic hearing loss (male and female participants together, comparing Older-Normal to Metabolic hearing loss), and the list of genes with high variant load in Metabolic hearing loss (male participants, comparing Metabolic hearing loss to Sensory hearing loss) (Table 2). The gene lists were also tested for enrichment in 1213 highly variable genes, which are genes frequently reported to carry variants in multiple exome sequencing projects (Lewis et al. 2022). A significant enrichment of highly variable genes was found in multiple gene lists (Table 2), suggesting that some of the genes included are present for reasons unrelated to hearing. The outlier lists were combined to obtain a final candidate list of 291 genes, 18 of which were known deafness genes and 37 of which were highly variable genes (Suppl. Table 4). 107 of these genes were also identified in our previous study of self-reported hearing difficulty in the UK BioBank cohort (Lewis et al. 2022), 11 of which were known deafness genes (*ELMO3*, *CDH23*, *UBE3B*, *ADGRV1*, *COL9A3*, *NAV2*, *DMD*, *AFAP1L2*, *MPDZ*, *LOXHD1*, and *CELSR1*).

To prioritise the list of candidate genes, a third list of outlier genes was obtained from the TwinsUK cohort. In this case there were not sufficient participants classified as having Sensory hearing loss, and so only two comparisons were carried out (Figure 3 J,K; Table 2), resulting in a final candidate list of 436 genes, including 23 known deafness genes and 43 highly variable genes (Suppl. Table 5).

Thirty-eight genes were common to all three analyses (Figure 4), one of which was a known deafness gene (*PKHD1L1*) (Figure 4). Of these 38 genes, 32 had good quality mouse orthologues. The expression of these 32 genes in the mouse inner ear was investigated using publicly available single cell RNAseq data from the gEAR public expression resource (Orvis et al. 2021). Eleven genes had no expression reported in the chosen ages and cell types, and a further eleven genes were expressed at low levels (up to and including the expression level of *Hprt1*, to which all expression was normalised). The remaining 10 genes were strongly expressed in at least one cell type and age (Suppl. Figure 5). Based on this analysis, among the most interesting novel candidate genes were *FKBP2* and *SYNE2*, which have strong expression in multiple cochlear and lateral wall cell types, and *ABCB8*, which shows similar expression to the hair cell marker *Myo7a* (Figures 5, 6).

**Figure 4.**
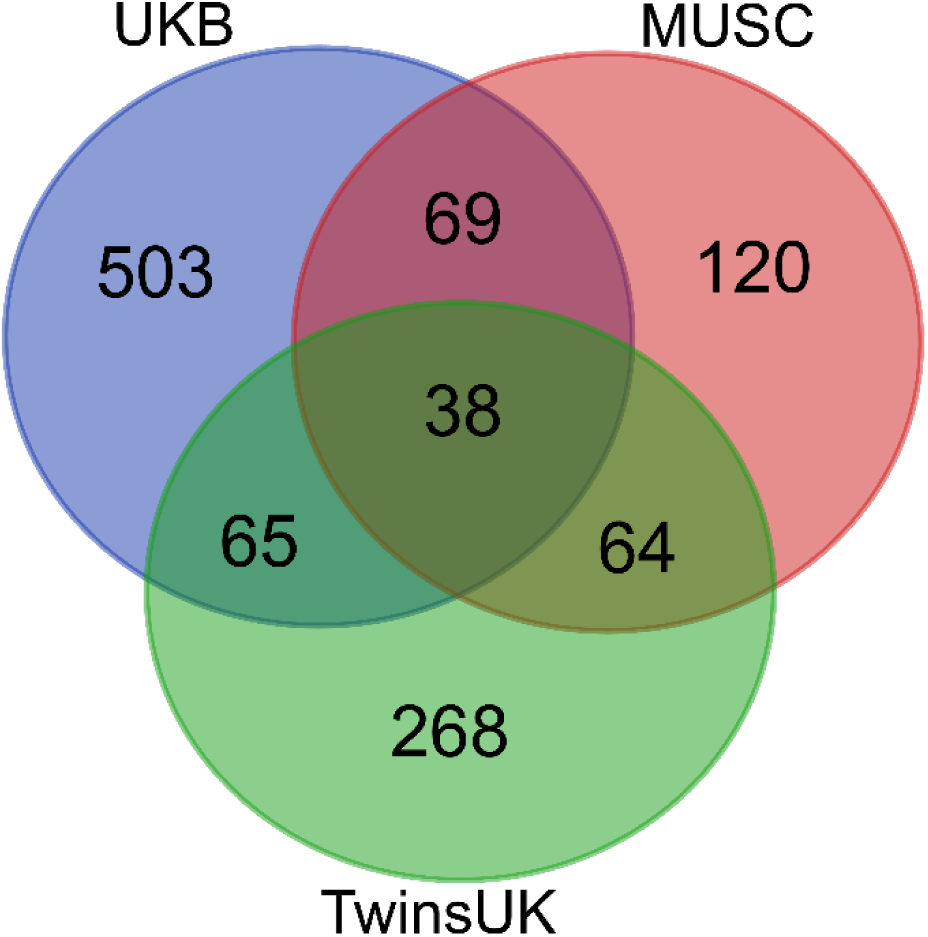
Overlap in gene lists from the two cohorts described in this study (genes listed in Supp Tables 4, 5) and the gene list from our previous study on the UK Biobank (Lewis et al. 2022).

**Figure 5.**
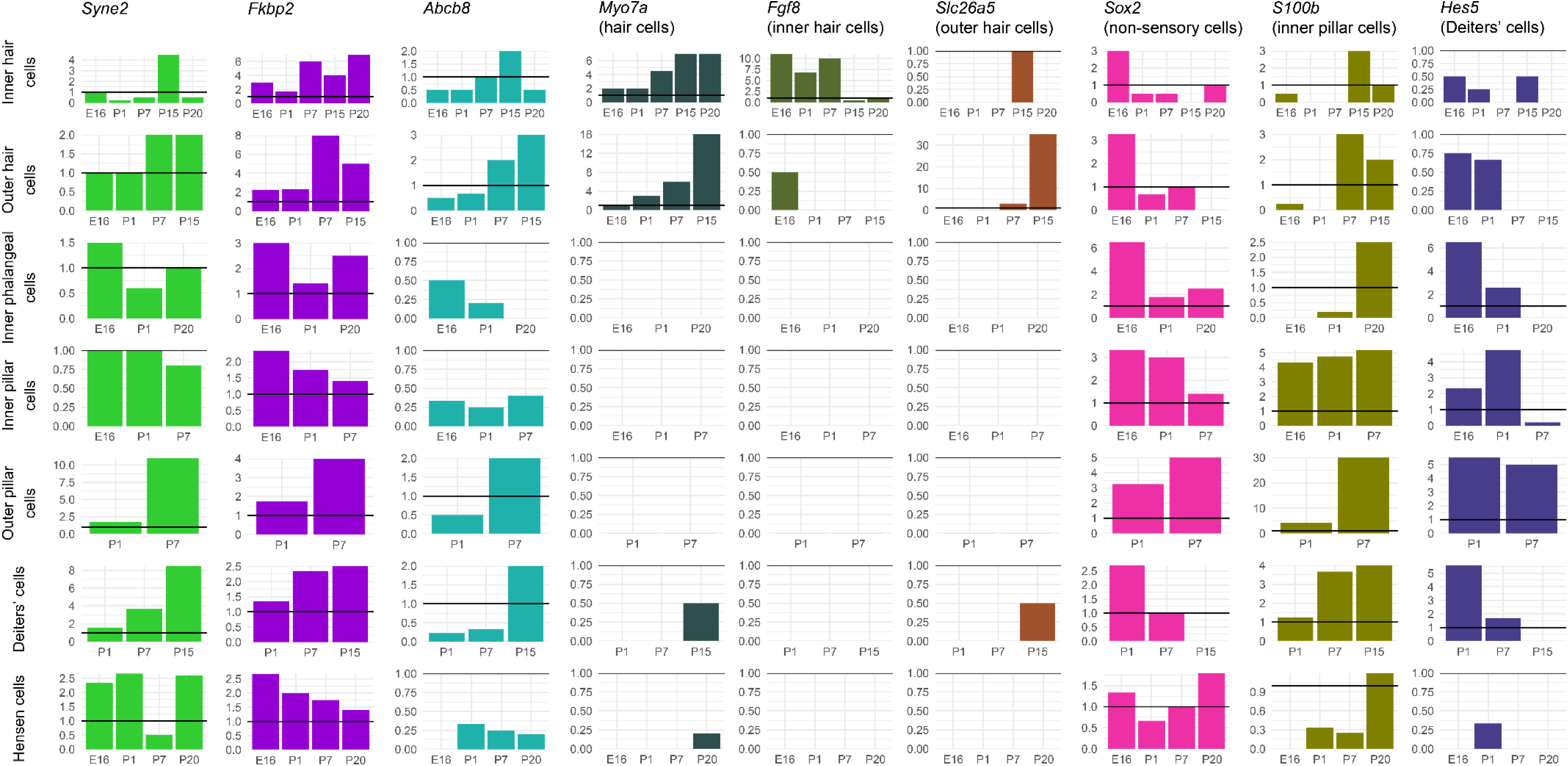
Expression levels at different developmental stages of key cell type marker genes and genes of interest from the exome sequence analysis, based on single cell RNAseq data from the gEAR (http://umgear.org). Expression was normalised to *Hprt* (represented by a horizontal line at y=1 on each plot). Marker genes included for comparison are *Myo7a* (hair cells), *Fgf8* (inner hair cells), *Slc26a5* (outer hair cells), *Sox2* (non-sensory cells), *S100b* (inner pillar cells) and *Hes5* (Deiters’ cells). *Syne2*, *Fkbp2* and *Abcb8* show interesting expression across different ages and cell types in the mouse organ of Corti; *Syne2* and *Fkbp2* are strongly expressed in multiple cochlear cell types, particularly supporting cells, and the expression pattern of *Abcb8* resembles that of *Myo7a*, both temporally and spatially.

In order to investigate genes associated with specific phenotypes, we also plotted the expression of genes identified only in the phenotype-specific analyses. There were 18 genes linked only to Metabolic hearing loss (including four deafness genes: *DMD*, *DUOX2*, *CELSR1* and *ELMO3*) and 54 genes linked only to Sensory hearing loss (including four deafness genes: *ARHGAP21*, *LMO7*, *UBE3B* and *ADGRV1*) (Suppl. Tables 4, 5). After removing genes without a good quality mouse orthologue and with low or no expression in the chosen inner ear datasets, we plotted the expression of 12 Metabolic-linked genes and 17 Sensory-linked genes (Suppl. Figure 6). The four Metabolic-linked genes most strongly expressed in the lateral wall are *MT-CO1*, *TLN2*, *DPP4* and *CHMP4C*, and are also expressed in several organ of Corti cell types (Suppl. Figure 6). The Sensory-linked genes most strongly expressed in the organ of Corti are *MADD*, *UBE3B* and *LMO7*, but they have low or no expression in the lateral wall (Suppl. Figure 6).

**Figure 6.**
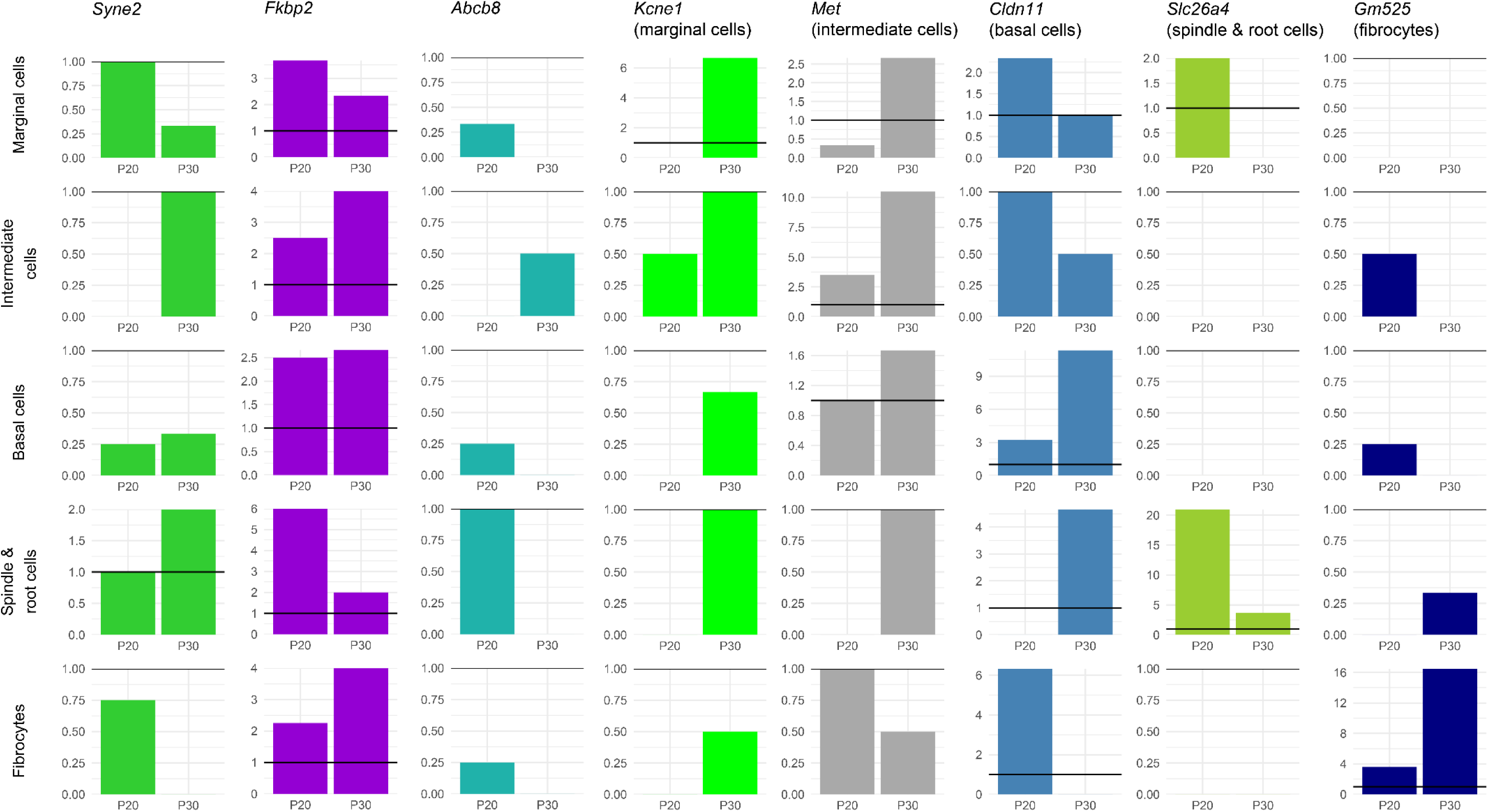
Expression of *Syne2*, *Fkbp2* and *Abcb8* in the mouse lateral wall, based on single cell RNAseq data from the gEAR (http://umgear.org). Expression was normalised to *Hprt* (represented by a horizontal line at y=1 on each plot). Marker genes have been included for comparison (*Kcne1* (marginal cells), *Met*(intermediate cells), *Cldn11* (basal cells), *Slc26a4* (spindle and root cells) and *Gm525* (fibrocytes)).

### Threshold difference detection

We compared the thresholds of carriers of each individual variant to those of non-carriers in order to assess the contribution of each variant to threshold differences. Forty of the 29,807 high impact variants in the nuclear exome passed the filters and permutation testing. In two cases (*KIRREL1* and *CCDC171*), both the non-segregated alternate allele group and one of the sex-segregated groups exhibited a significant difference in thresholds. In the remaining 38 cases, only one group exhibited a significant threshold difference. One mitochondrial variant (rs41518645, in *MT-CYB*) also was found to pass the filter and permutation tests, and was associated with better thresholds in male participants (Figure 7). There were no instances of multiple variants being identified in the same gene, and only one was in a known deafness gene, *S1PR2* (Table 3, Figure 7). Sixteen of the 41 variants were associated with better thresholds than the sex-matched reference group (eg *TCEANC2*, Figure 7), and 25 with worse thresholds than the reference group (eg *CLDN3*, Figure 7). Fifteen variants exhibited a significant difference in thresholds in only one sex (eg *S1PR2*, *HADH*, Figure 7), not including those instances where there were too few carriers of the opposite sex to determine if their thresholds were similarly affected, eg *CAPN9* (Figure 7, Table 3; all audiograms are shown in Suppl. Figure 7).

**Figure 7.**
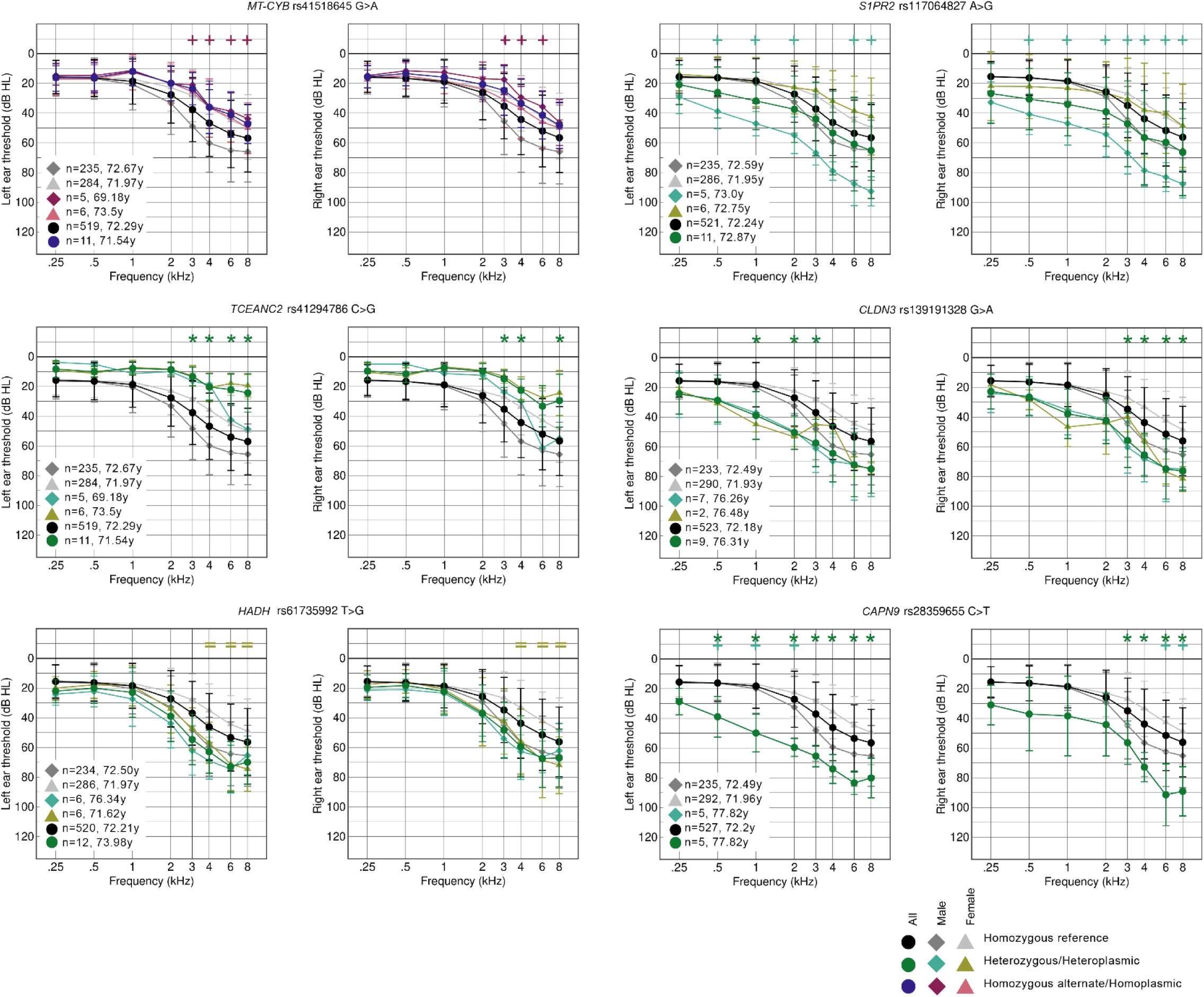
Average audiograms from the MUSC cohort plotted in groups by sex and genotype for six different variants (chosen as examples from the full list of 41; see Suppl. Figure 5). Two audiograms are shown for each variant; the thresholds from the left ear are shown on the left, and those from the right ear on the right. Numbers and average ages of each group are listed on the graph. The symbols at the top of each graph mark which groups passed the criteria for each stimulus frequency compared to the relevant reference group (+ for male, = for female, and * for all participants). Carriers of the *MT-CYB* and *TCEANC2* variants have better thresholds than non-carriers, and carriers of the *MMS19*, *S1PR2*, *CLDN3* and *CAPN9* variants have worse thresholds than non-carriers. The variants in *S1PR2* and *HADH* are linked to worse thresholds only in male and female carriers respectively, and there are no female carriers of the *CAPN9* variant so it is unknown whether they would be similarly affected to male carriers.

**Table 3.**
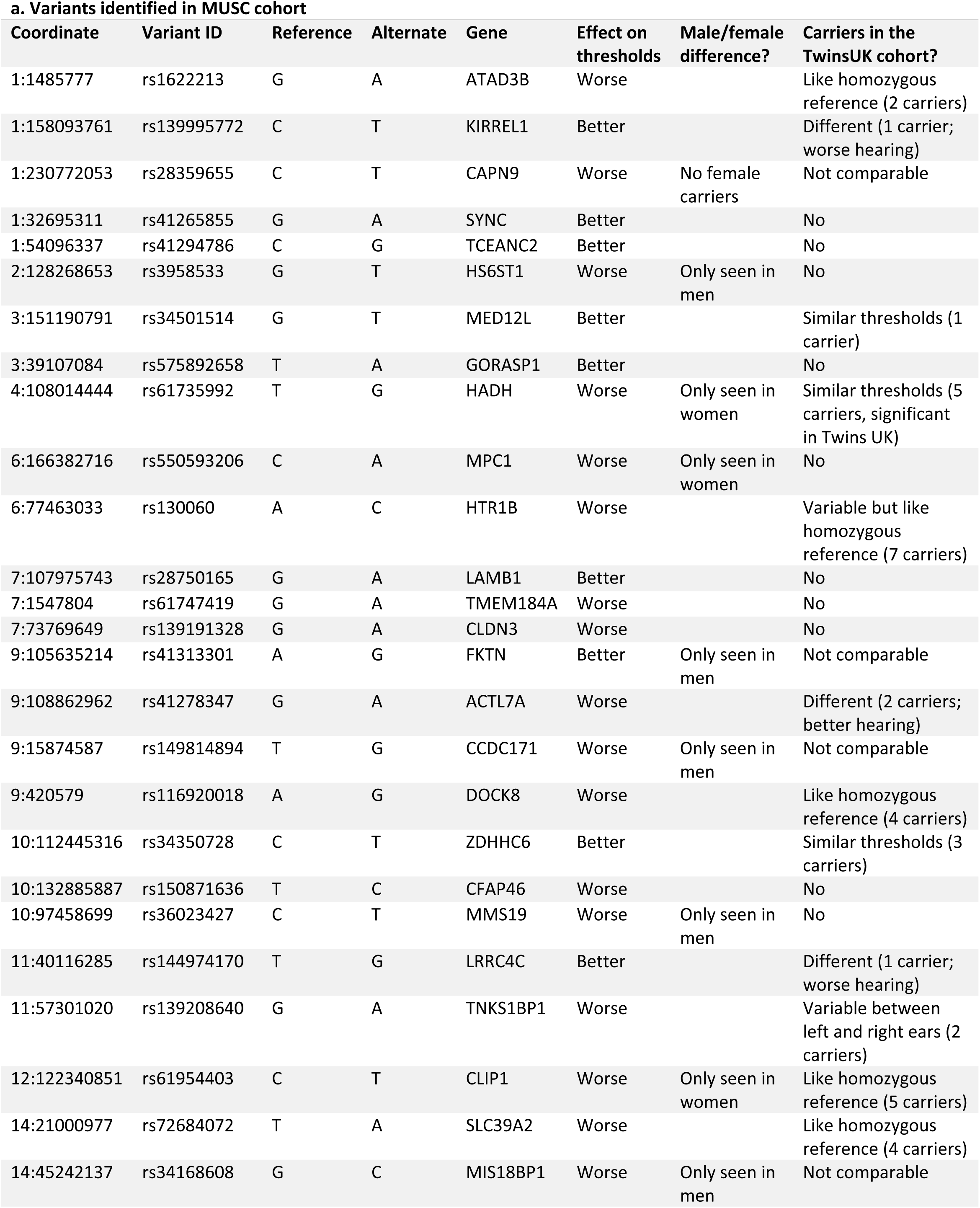
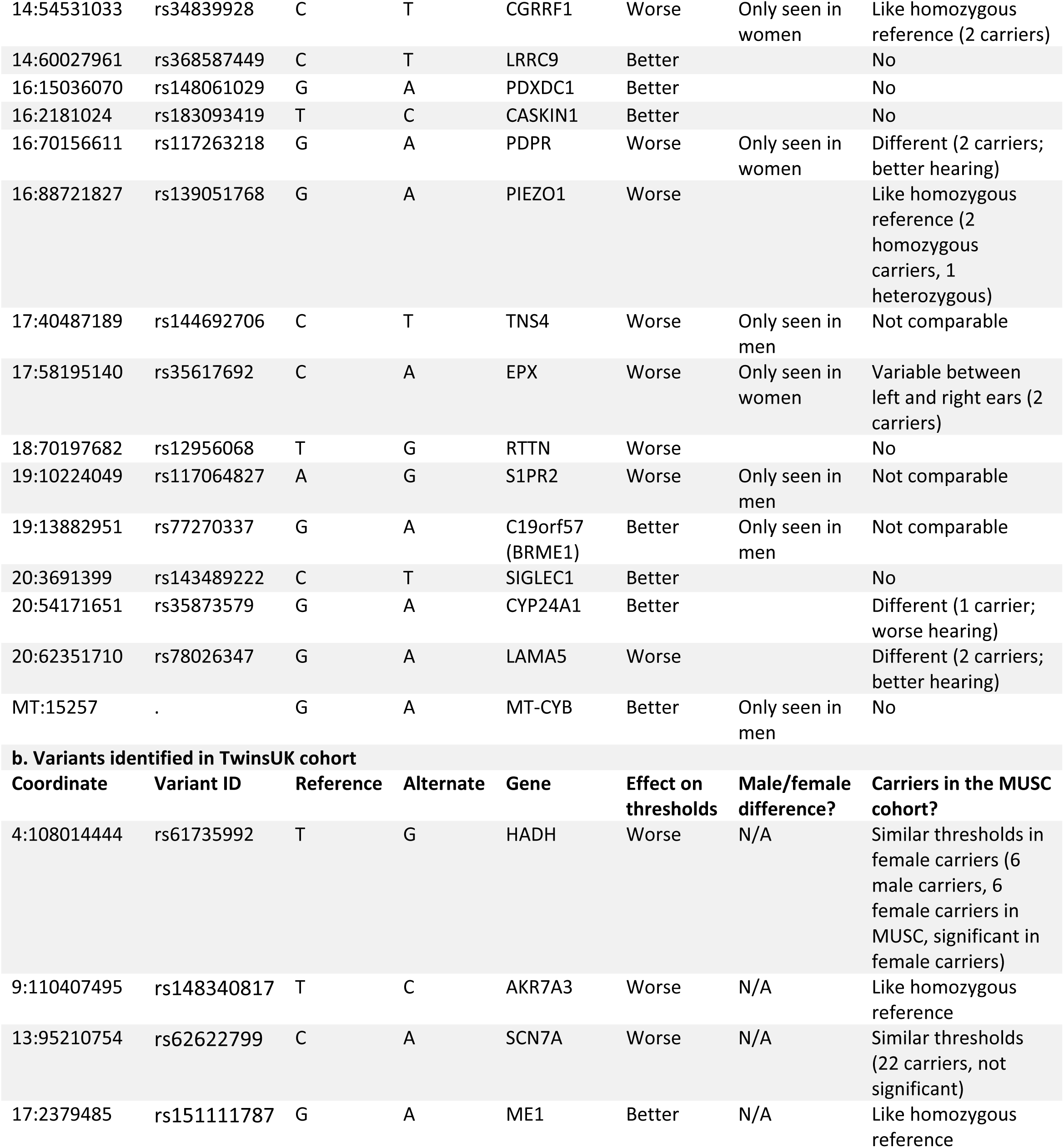
Details of variants identified by associated threshold differences in the MUSC (a) and TwinsUK (b) cohorts. Variants are ordered by genomic location.

To further investigate the contribution of these 41 variants to threshold differences, carriers of each variant in the TwinsUK cohort were identified, and their audiograms plotted compared to homozygous carriers of the reference allele. Sixteen of the 41 variants were not found in any of the TwinsUK participants (Table 3), and for a further seven variants, the threshold difference was only seen in male MUSC carriers, not in female participants, so a comparison was not possible with the all-female TwinsUK cohort (Table 3). However, five carriers from the TwinsUK cohort were found to have similar audiograms to those in the MUSC cohort for the variant in *HADH*, and this was also found independently when the same filter and permutation testing was carried out on the TwinsUK cohort. Carriers of variants in *MED12L* (n=2), and *ZDHHC6* (n=3) also had a similar average threshold shape to that seen in the MUSC cohort carriers (Table 3, Suppl. Figure 7), supporting the suggestion of a potential role for these variants in contributing to the hearing loss seen in carriers. We examined the MUSC carriers of the variants in *HADH*, *MED12L* and *ZDHHC6* to check for any variants in 50 known dominant deafness genes (https://hereditaryhearingloss.org, accessed March 2023 (Van Camp and Smith)) but did not find any dominant gene consistently affected within each group.

From the TwinsUK cohort alone, only four variants passed the filters and permutation testing, one of which was the variant in *HADH*, also identified in the MUSC cohort. The other three genes were *AKR7A3*, *SCN7A* and *ME1* (Suppl. Figure 7). There were many carriers of each of these three variants in the MUSC cohort, but for *ME1* and *AKR7A3*, the average thresholds of carriers did not show any obvious difference to non-carriers, suggesting that if these variants do contribute to hearing loss, the impact is not reflected in audiogram shape (Suppl. Figure 5). MUSC carriers of the variant in *SCN7A* (n=22; 9 female, 13 male) had, on average, slightly worse thresholds than homozygous carriers of the reference allele, resembling the thresholds of carriers in the TwinsUK cohort, but the difference was not significant (Suppl. Figure 7).

## Discussion

From the outlier analysis, we identified 38 candidate genes that may contribute to overall hearing status, 18 genes linked to Metabolic hearing loss alone, and 54 genes linked to Sensory hearing loss alone. The threshold analysis revealed 41 candidate genes including one known deafness gene (*S1PR2*). One gene, *GORASP1*, was identified from both analyses, since MUSC carriers of the rs575892658 missense variant had improved thresholds (Suppl. Fig 7), and *GORASP1* was linked specifically to Metabolic hearing loss through the TwinsUK outlier analysis (Suppl. Table 5).

### Known deafness genes from the candidate gene lists

Our candidate gene lists include 10 deafness genes; *S1PR2*, *PKHD1L1*, *DMD*, *DUOX2*, *CELSR1*, *ELMO3*, *ARHGAP21*, *LMO7*, *UBE3B* and *ADGRV1*. Only 3 of these have been identified in humans; *ADGRV1*, which is an Usher syndrome type II gene (Weston et al. 2004), *DMD*, which has been associated with congenital hearing impairment as well as muscular dystrophy (Pfister et al. 1998), and *S1PR2,* mutations in which lead to congenital profound hearing impairment (Santos-Cortez et al. 2016), although a point mutation in *S1pr2* in mice results in early-onset progressive hearing loss (Ingham et al. 2016). These phenotypes are more severe than the late-onset progressive hearing loss in our human subject cohorts, which supports the theory that genes responsible for severe deafness may also be involved in milder forms of hearing loss.

*PKHD1L1*, *DUOX2*, *CELSR1*, *ELMO3*, *ARHGAP21*, *LMO7* and *UBE3B* are all defined as deafness genes through work on the mouse orthologues. Of these, hearing loss caused by mutant alleles of *Arhgap21* and *Elmo3* have only been reported by the IMPC large-scale phenotyping screen (www.mousephenotype.org (Dickinson et al. 2016; Groza et al. 2023)); *Elmo3* homozygous mutants have raised thresholds at low frequencies (https://www.mousephenotype.org/data/genes/MGI:2679007) and *Arhgap21* heterozygous mutants exhibit variably raised thresholds across most frequencies tested (https://www.mousephenotype.org/data/genes/MGI:1918685). Mice with a disrupted *Ube3b* gene display mild hearing impairment at all frequencies at 3 months old, and this impairment was more severe when tested at 6 months old (Basel-Vanagaite et al. 2012). *Pkhd1l1* mutants show early-onset progressive hearing loss (Wu et al. 2019), while abolishing *Lmo7* expression in mice results in late-onset progressive hearing impairment (Du et al. 2019). All these are comparatively mild effects, but mice carrying a missense mutation in *Duox2* have severely raised thresholds (Johnson et al. 2007), and mice carrying mutations in the planar cell polarity gene *Celsr1* exhibit vestibular defects and misoriented outer hair cells (Curtin et al. 2003).

### Expression analysis of novel candidate genes

Interestingly, most of our newly associated genes have not previously been reported with hearing loss, suggesting that there are many more genes involved in hearing which remain to be identified. Our expression analysis results suggest some promising genes for further investigation, such as *SYNE2*, *FKBP2*, and *ABCB8* from the main analysis, and *MADD* and *CHMP4C* from the phenotype-specific analysis. SYNE2 forms part of the LINC (Linker of

Nucleoskeleton and Cytoskeleton) Complex, which is part of the nuclear envelope and is essential for maintenance of normal hearing (Horn et al. 2013). *FKBP2* encodes FKBP13, a luminal endoplasmic reticulum (ER) protein which is upregulated in response to cellular stress such as heat shock, or the accumulation of unfolded protein precursors in the ER (Nigam et al. 1993; Partaledis and Berlin 1993), and *ABCB8* is a mitochondrial ABC transporter which plays a role in cellular viability and is protective against oxidative stress (Ardehali, O’Rourke, and Marban 2005); mutations in either may contribute to the vulnerability of inner ear cells to damage and age-related deterioration. *MADD*, which was associated with Sensory hearing loss and has an expression pattern resembling that of *Myo7a* (Suppl. Figure 6), is an activator of the Rab3 small GTP-binding protein family, and has been shown to be critical for neurotransmitter release in neuromuscular junctions and in hippocampal neurons (Tanaka et al. 2001; Yamaguchi et al. 2002); it also may play a role in inner ear synapses but that has yet to be determined. *CHMP4C*, which was associated with Metabolic hearing loss in the TwinsUK cohort, is expressed in the marginal and basal cells of the stria vascularis, as well as several cell types in the organ of Corti. Previous whole exome sequencing and genome-wide association studies have also linked *CHMP4C* to hearing impairment, suggesting it is a good candidate for further study (Ivarsdottir et al. 2021; Kalra et al. 2020; Lewis et al. 2022; Wells et al. 2019). However, it should be noted that gene expression in a particular cell type is not a guarantee of a critical role in that cell type, and the absence of expression in inner ear cells at the times and stages studied does not preclude a gene from having a role in hearing. It may be needed at a later time in development, or elsewhere in the auditory pathway, or may only be needed only at very low quantities, making it difficult to detect by single cell RNAseq. Also, given the limited data available from expression studies, a role for the other candidate genes and variants in age-related hearing loss should not be discounted.

### Novel candidate genes from the threshold analyses

From our threshold analyses on both cohorts, we identified a variant in the gene *HADH* as a candidate associated with worse hearing, and variants in the genes *ZDHHC6* and *MED12L* were associated with better hearing (Suppl. Figure 7). HADH (hydroxyacyl-Coenzyme A dehydrogenase) localises to the mitochondrial matrix where it plays a role in the beta-oxidation pathway, breaking down fatty acid molecules to generate acetyl-coA. Mutations in other genes in the same pathway have been shown to result in mitochondrial dysfunction (Foomani et al. 2021), suggesting a potential mechanism for *HADH* variants to affect hearing. ZDHHC6 is a palmitoyltransferase located in the ER, and defects in palmitoylation have been linked to hearing impairment (Steinke et al. 2015). MED12L is a subunit of the Mediator protein complex which is part of the basal transcriptional apparatus; post-natal deletion of the Med12 subunit of the same complex in mice results in rapid loss of basal cell organisation and disruption of the stria vascularis leading to hearing loss (Huang et al. 2021).

### The genetic contribution to hearing differences between sexes

The MUSC cohort has a slight excess of female participants over male, but the difference in classification of their hearing is marked, with too few male participants classified as “Older-Normal” to carry out a robust regression analysis on men alone using that category (Table 1, Figure 2). This difference has been previously described in multiple studies (Cruickshanks et al. 1998; Davis and Research 1995; Dubno et al. 2008; Dubno et al. 1997; Helzner et al. 2005; Lee et al. 2005; Pearson et al. 1995), with hearing in women tending to be better than in men and declining later in life, generally around the onset of menopause (Davis and Research 1995; Hederstierna et al. 2010). However, the average age of the participants in the MUSC cohort is over 60, suggesting that there is also a genetic contribution to the difference in hearing impairment between the sexes, as observed in our previous study (Lewis et al. 2022).

The other clear difference in auditory phenotype between the sexes can be seen in the number of men classified as having Sensory hearing loss (101, 42% of male participants) compared to women (53, 18% of female participants) (Table 1). The proportions in the Metabolic hearing loss group are the inverse, although not so extreme (62 men, 26% of male participants, and 92 women, 32% of female participants). However, in the all-female TwinsUK cohort, there are only 7 participants classified as having Sensory hearing loss (5%, not including the twins who were removed; Table 1). Sensory hearing loss has been attributed to noise exposure, among other factors, and most of the men in the MUSC cohort had a positive noise exposure history (189, 79% of all male participants). However, the proportion of men reporting a history of noise exposure across the three classified groups was broadly similar (Older-Normal: 67%; Sensory: 79%; and Metabolic: 69%) (Table 1, Figure 2). The proportion of women in the MUSC cohort reporting a positive noise history in the different classifications was also very similar (Table 1, Figure 2). Self-reported noise history alone thus does not explain the excess of male participants classified as having Sensory hearing loss in the MUSC cohort. There may be a sex-specific genetic contribution to this observation, but more data are needed for further exploration. A more objective, quantifiable measure of noise exposure would also help in this, since noise history questionnaires can be an unreliable measure of an individual’s noise exposure.

The only regression analysis which could be performed using male participants alone from the MUSC cohort was the comparison of variant counts in men classified with Sensory hearing loss versus those classified with Metabolic hearing loss (Figure 3). Some of the genes with a high variant load in Sensory hearing loss are found in both the male and female lists, but none of the genes with a high variant load in Metabolic hearing loss are shared between the sexes (Figure 8). It is possible that there is a higher sex-specific genetic contribution to Metabolic hearing loss, but more data from larger cohorts are needed to explore this further. We also found multiple variants which appear to contribute to differences in thresholds in a sex-specific manner (Table 3), although the lack of audiograms and exome sequencing from male participants in the TwinsUK cohort means that we have not been able to follow up on those variants linked to threshold differences visible only in men from the MUSC cohort. Similarly well-characterised cohorts are necessary for further investigating the differing genetic contribution to hearing loss between the sexes.

**Figure 8.**
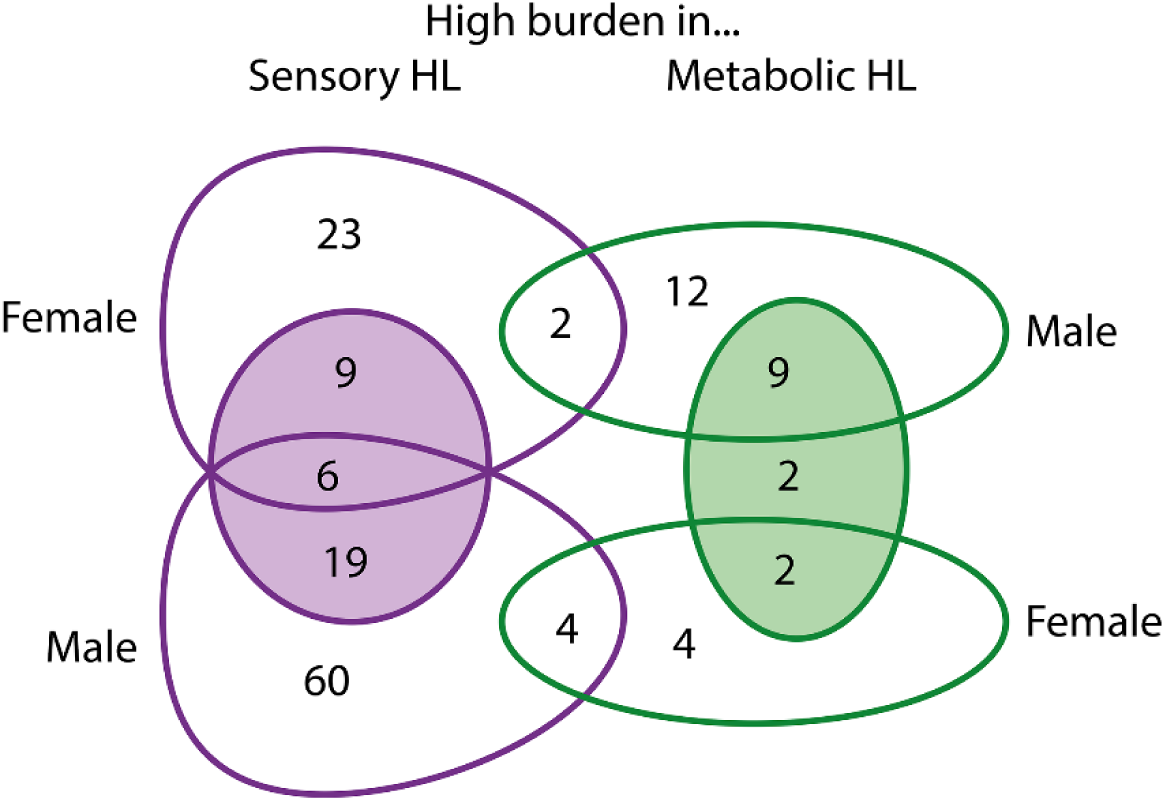
Venn diagram showing the overlap of genes identified as having a high variant load in Metabolic or Sensory hearing loss in all, male and female participants in the MUSC cohort. The shaded circles show the high variant load gene counts identified in all participants.

### Candidate genes and variants associated with better hearing in older adults

Intriguingly, a subset of candidates from both outlier and threshold analyses are associated with better hearing, suggesting that some variants may lead to protection against age-related hearing loss and/or protection against damage from noise exposure. This is not the first report of protective mutations. Examples of other protective variants include the N352S variant in *B4GALT1* which is protective against cardiovascular disease (Montasser et al. 2021) and protein-truncating variants in *GPR75* which reduce the risk of obesity (Akbari et al. 2021), as well as the A88V variant in *Gjb6*, which protects against hearing loss in mice (Kelly et al. 2019). Identification of genes and variants which protect against hearing loss could be a useful starting point for developing therapeutic treatments to do the same.

## Data Availability

Data availability: Access to the TwinsUK data used in this study may be requested through the TwinsUK registry at https://twinsuk.ac.uk/. The mouse single cell RNAseq data is publicly available at https://umgear.org/. De-identified datasets from the MUSC cohort are available to researchers upon request and completion of institutional data use agreements, as required by the Medical University of South Carolina.

https://umgear.org/

## Acknowledgements

We thank Elysia James for help with annotation. We are grateful to the participants in each cohort for contributing their data.

## Data Availability

Access to the TwinsUK data used in this study may be requested through the TwinsUK registry at https://twinsuk.ac.uk/. The mouse single cell RNAseq data is publicly available at https://umgear.org/. De-identified datasets from the MUSC cohort are available to researchers upon request and completion of institutional data use agreements, as required by the Medical University of South Carolina.

