## Supplementary Figures for "Accurate phenotypic classification and exome sequencing allow identification of novel genes and variants associated with adult-onset hearing loss"

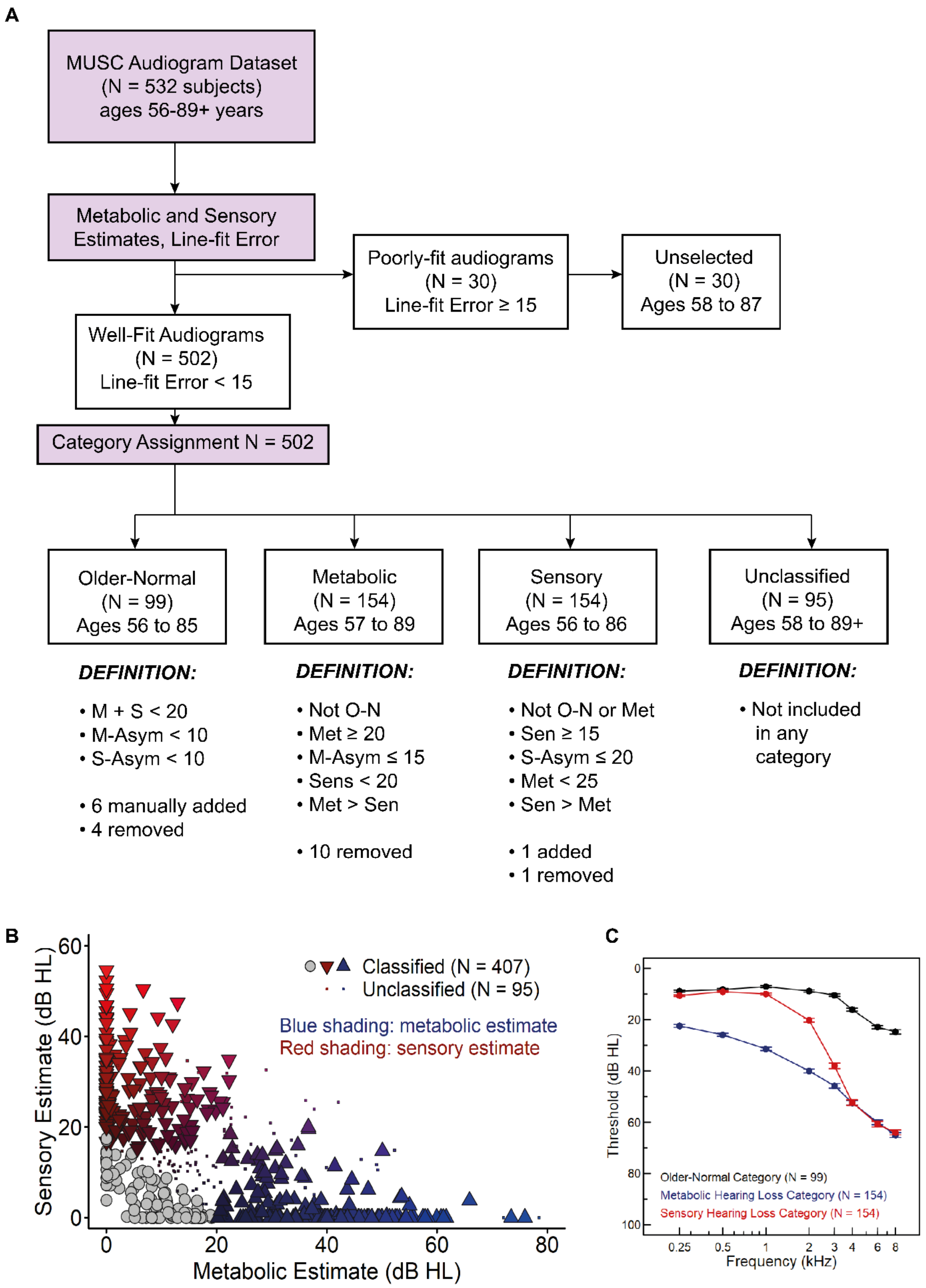

**Suppl. Figure 1.** A, Flow chart showing the audiogram classification procedure for the MUSC cohort. B, plot showing the sensory estimate (how much of the hearing loss observed can be attributed to sensory hearing loss) against the metabolic estimate (how much of the hearing loss observed can be attributed to metabolic hearing loss) for each case, with the blue/red shading indicating the magnitude of each estimate (metabolic/sensory respectively). The small dots are the Unclassified cases; Unselected cases are not shown. The grey circles near the origin are the Older-Normal participants, with very small contributions of both Sensory and Metabolic hearing loss. The red-shaded triangles aligned along the Y axis are those people with purely sensory contributions to their hearing loss, and the blue-shaded triangles along the X axis are those people with purely metabolic contributions to their hearing loss. However, there are a substantial number of people with mixed contributions, and these are the mixed shades in the middle of the graph. C, mean audiograms for the selected cases assigned to each category. Error bars are SEM.

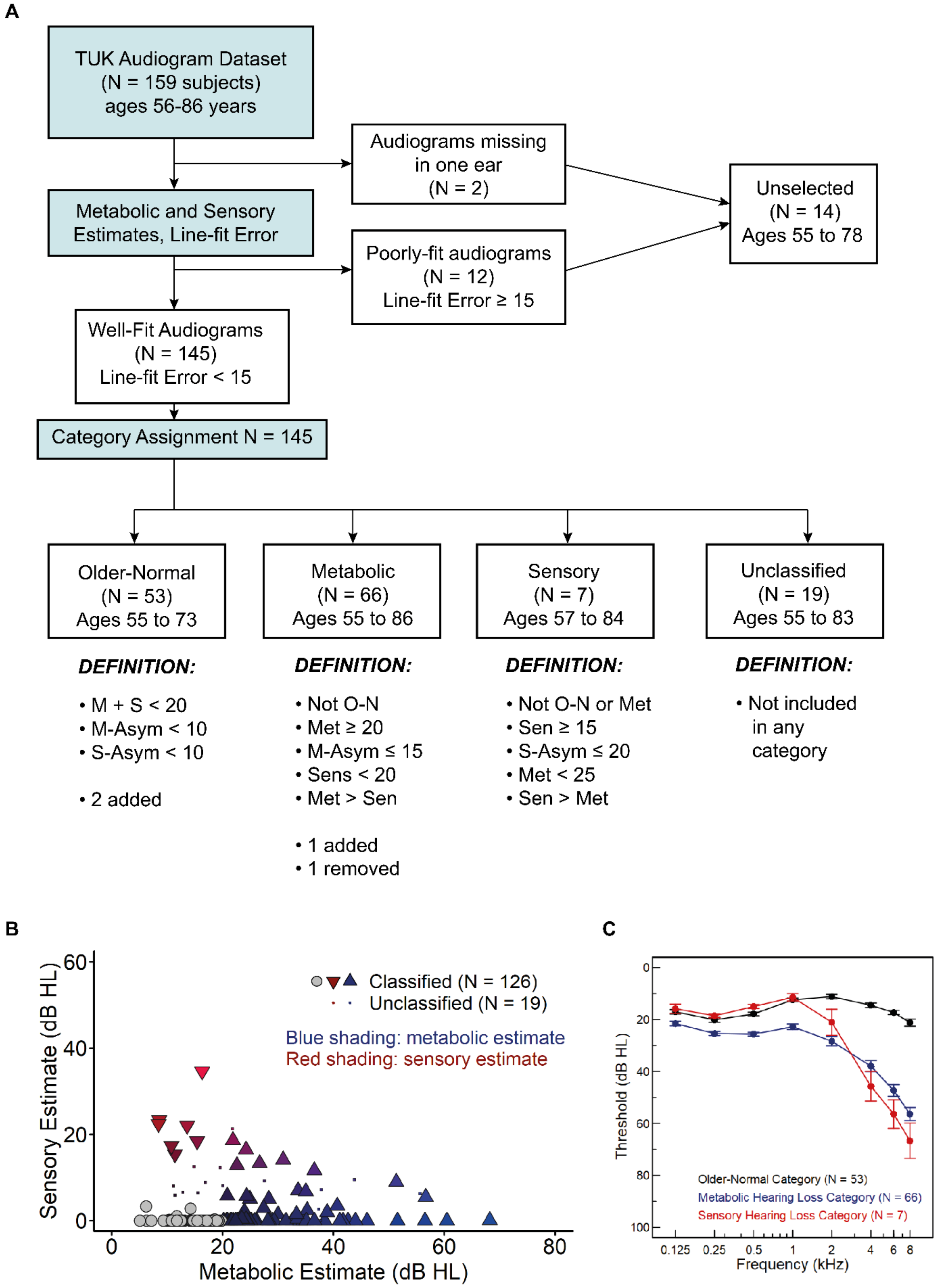
A
TUK Audiogram Dataset
(N = 159 subjects)
ages 56-86 years
Audiograms missing
in one ear
(N = 2)
Unselected
(N = 14)
Ages 55 to 78
Metabolic and Sensory
Estimates, Line-fit Error
Poorly-fit audiograms
(N = 12)
Line-fit Error ≥ 15
Well-Fit Audiograms
(N = 145)
Line-fit Error < 15
Category Assignment N = 145
Older-Normal
(N = 53)
Ages 55 to 73
Metabolic
(N = 66)
Ages 55 to 86
Sensory
(N = 7)
Ages 57 to 84
Unclassified
(N = 19)
Ages 55 to 83
DEFINITION:
• M + S < 20
• M-Asym < 10
• S-Asym < 10
• 2 added
DEFINITION:
• Not O-N
• Met ≥ 20
• M-Asym ≤ 15
• Sens < 20
• Met > Sen
• 1 added
• 1 removed
DEFINITION:
• Not O-N or Met
• Sen ≥ 15
• S-Asym ≤ 20
• Met < 25
• Sen > Met
DEFINITION:
• Not included in any category
B
Classified (N = 126)
Unclassified (N = 19)
Blue shading: metabolic estimate
Red shading: sensory estimate
Sensory Estimate (dB HL)
Metabolic Estimate (dB HL)
C
Threshold (dB HL)
Frequency (kHz)
Older-Normal Category (N = 53)
Metabolic Hearing Loss Category (N = 66)
Sensory Hearing Loss Category (N = 7)

**Suppl. Figure 2.** A, Flow chart showing the audiogram classification procedure for the TwinsUK cohort. B, plot showing the sensory estimate (how much of the hearing loss observed can be attributed to sensory hearing loss) against the metabolic estimate (how much of the hearing loss observed can be attributed to metabolic hearing loss) for each case, with the blue/red shading indicating the magnitude of each estimate (metabolic/sensory respectively). The small dots are the Unclassified cases; Unselected cases are not shown. The grey circles near the origin are the Older-Normal participants, with very small contributions of both Sensory and Metabolic hearing loss. The blue-shaded triangles along the X axis are those people with purely metabolic contributions to their hearing loss. There are no participants with a purely sensory contribution to their hearing loss, but there are a few with a Sensory classification, shown by redder shading and their high y coordinate. Most people with mixed contributions have a comparatively small Sensory component, and are classified as Metabolic (the blue-purple triangles on the right of the plot). C, mean audiograms for the selected cases assigned to each category. Error bars are SEM.

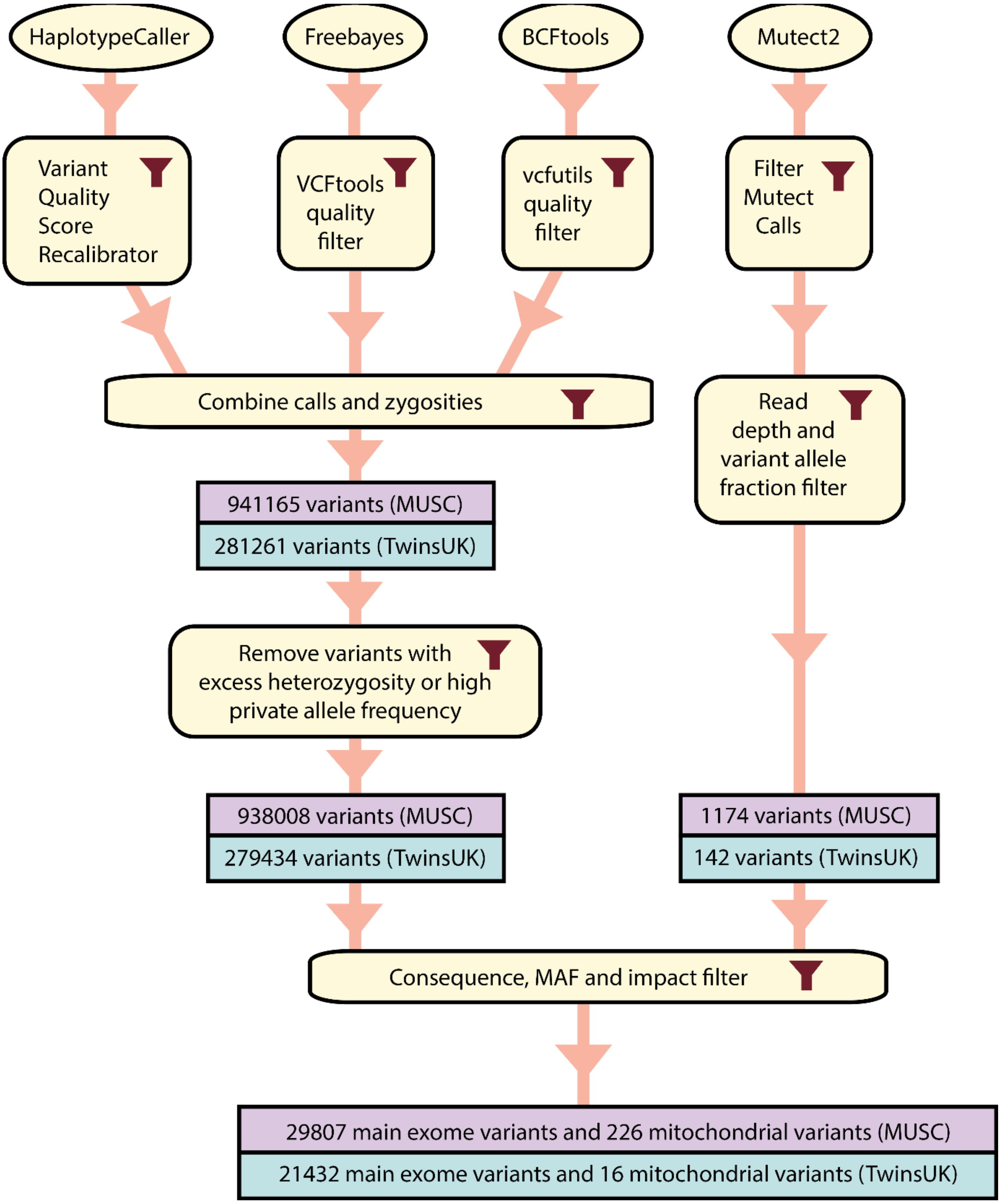


**Suppl. Figure 3.** Flow chart showing the variant filtering pipeline. See Suppl. Table 2 for details of the filter settings.

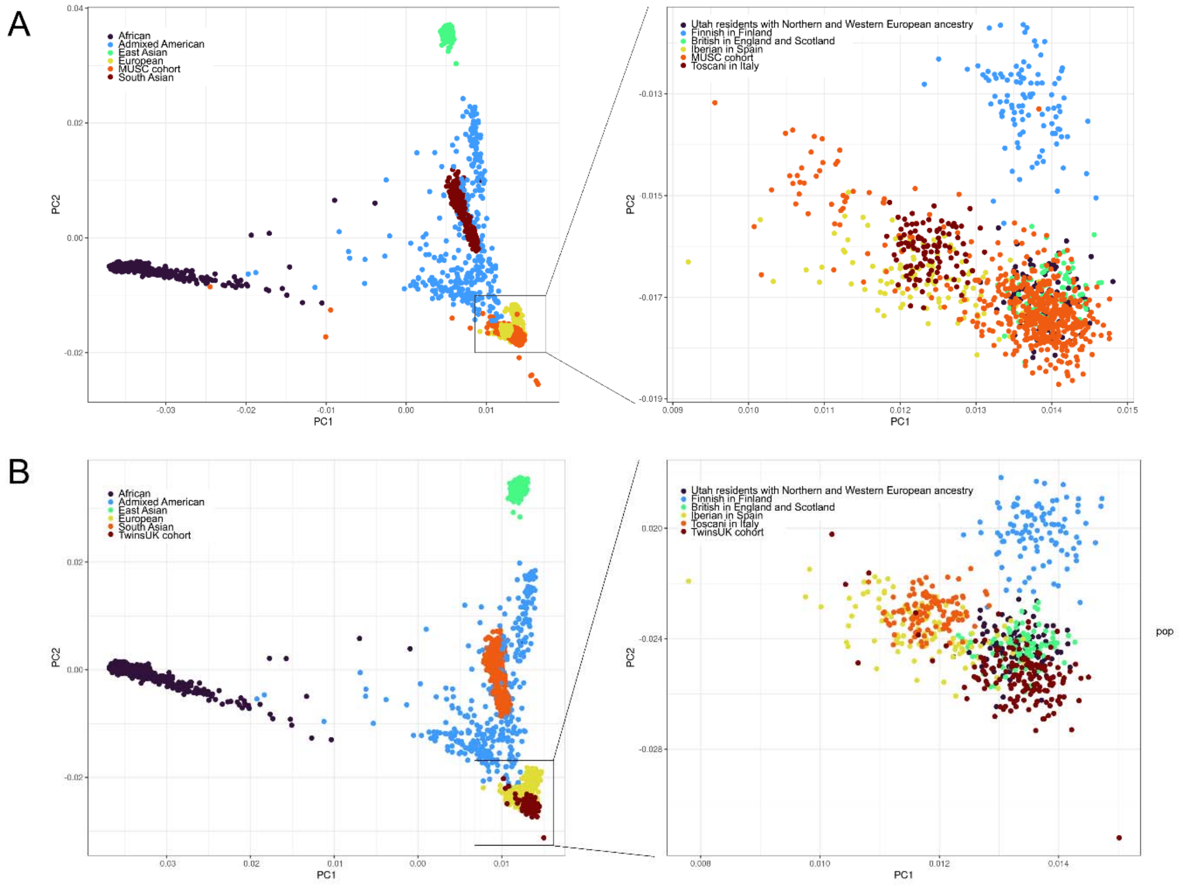


**Suppl. Figure 4.** Principal Component Analysis plots showing the genetic ancestry of the two cohorts used in this study compared to 2504 individuals from the 1000 Genomes study (Genomes Project et al. 2015). A shows the MUSC cohort, and B the TwinsUK cohort. On the left-hand plot, 1000 Genomes samples are coloured by their superpopulation, while the right-hand plot is a close-up showing just the European subpopulations.

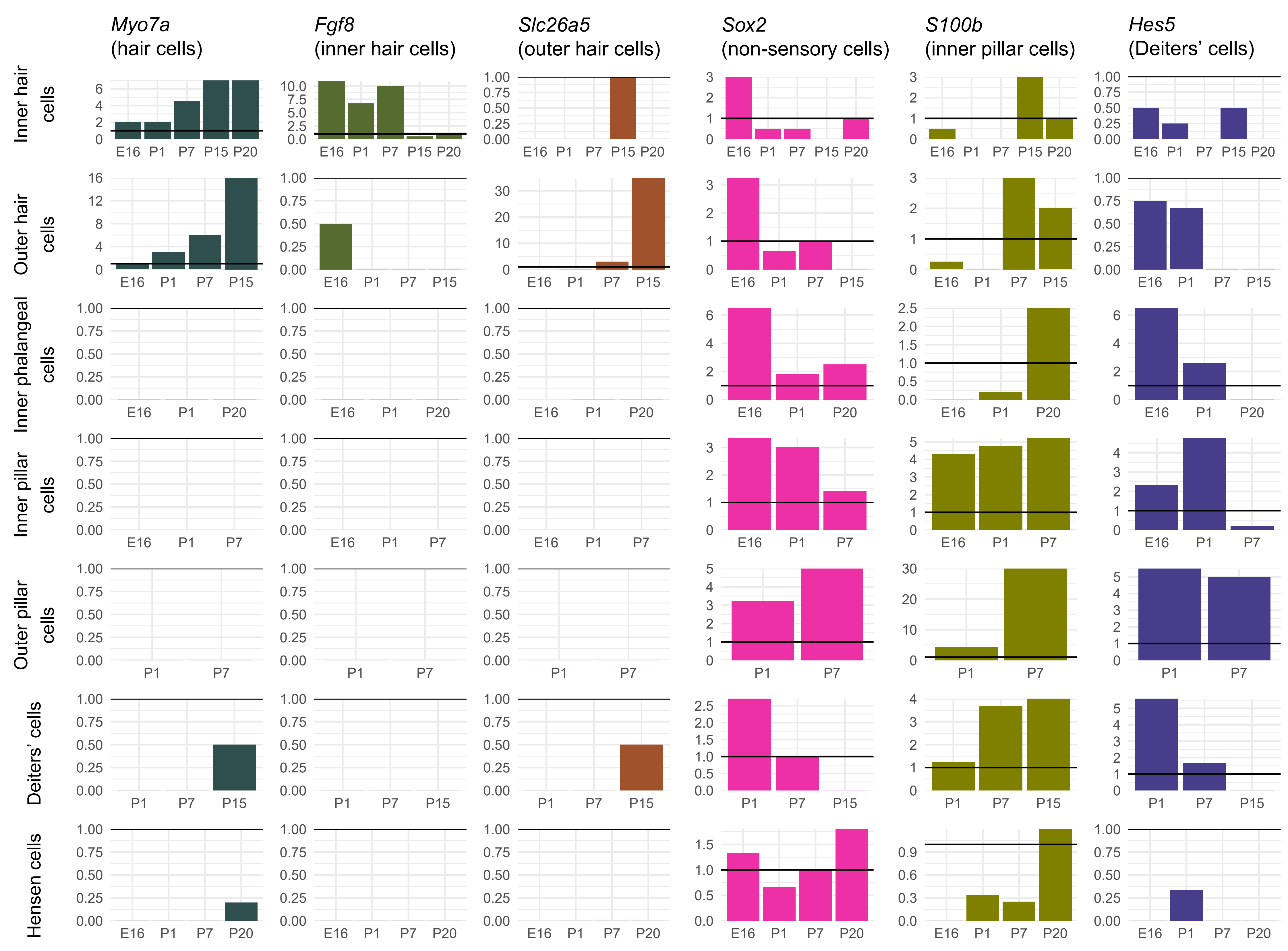
Myo7a
(hair cells)
Fgf8
(inner hair cells)
Slc26a5
(outer hair cells)
Sox2
(non-sensory cells)
S100b
(inner pillar cells)
Hes5
(Deiters' cells)
Inner hair cells
Outer hair cells
Inner phalangeal cells
Inner pillar cells
Outer pillar cells
Deiters' cells
Hensen cells
E16
P1
P7
P15
P20

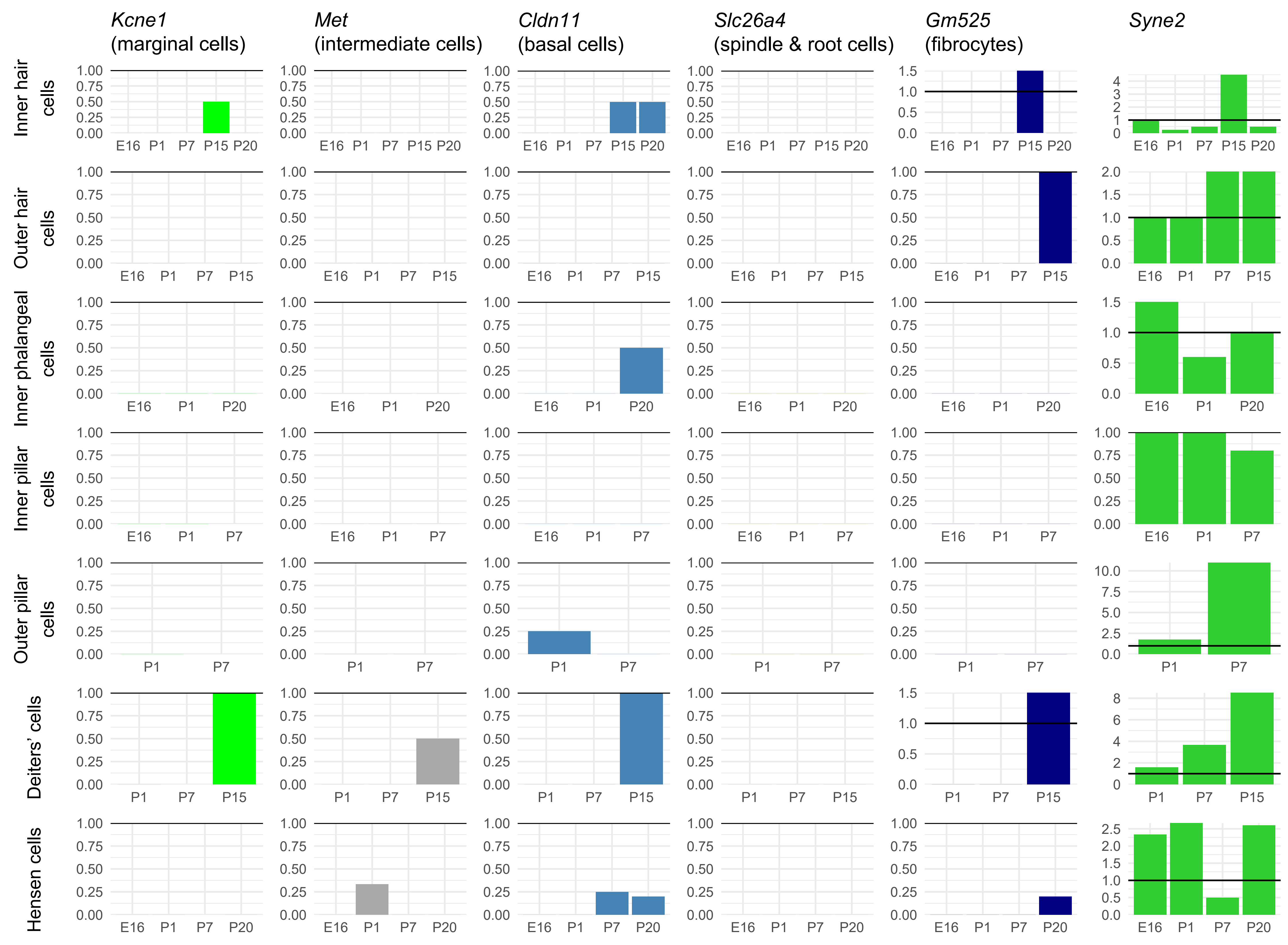

Kcne1 (marginal cells)
Met (intermediate cells)
Cldn11 (basal cells)
Slc26a4 (spindle & root cells)
Gm525 (fibrocytes)
Syne2
Inner hair cells
Outer hair cells
Inner phalangeal cells
Inner pillar cells
Outer pillar cells
Deiters' cells
Hensen cells
E16
P1
P7
P15
P20

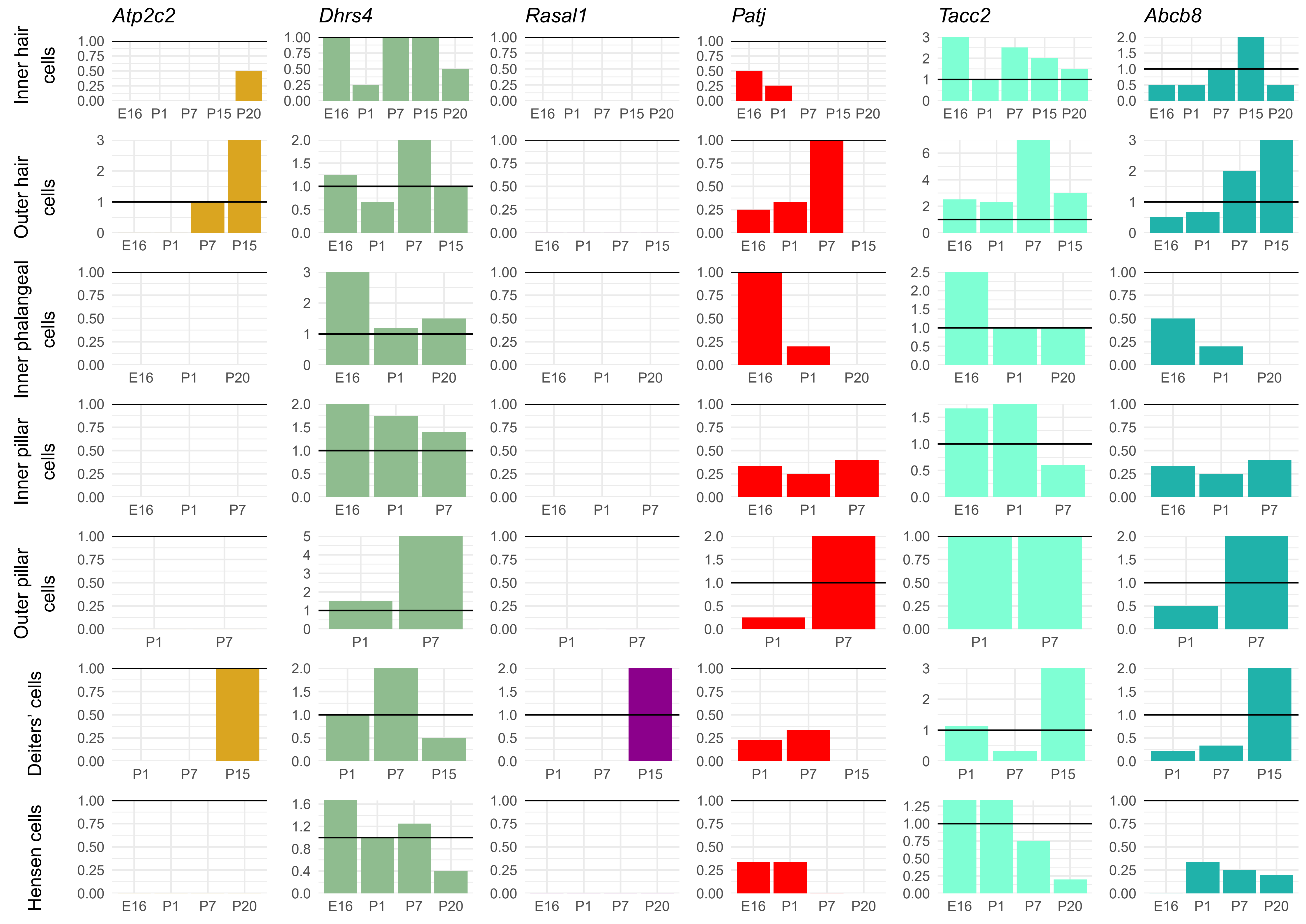

Atp2c2
Dhrs4
Rasal1
Patj
Tacc2
Abcb8
Inner hair cells
Outer hair cells
Inner phalangeal cells
Inner pillar cells
Outer pillar cells
Deiters' cells
Hensen cells
E16
P1
P7
P15
P20

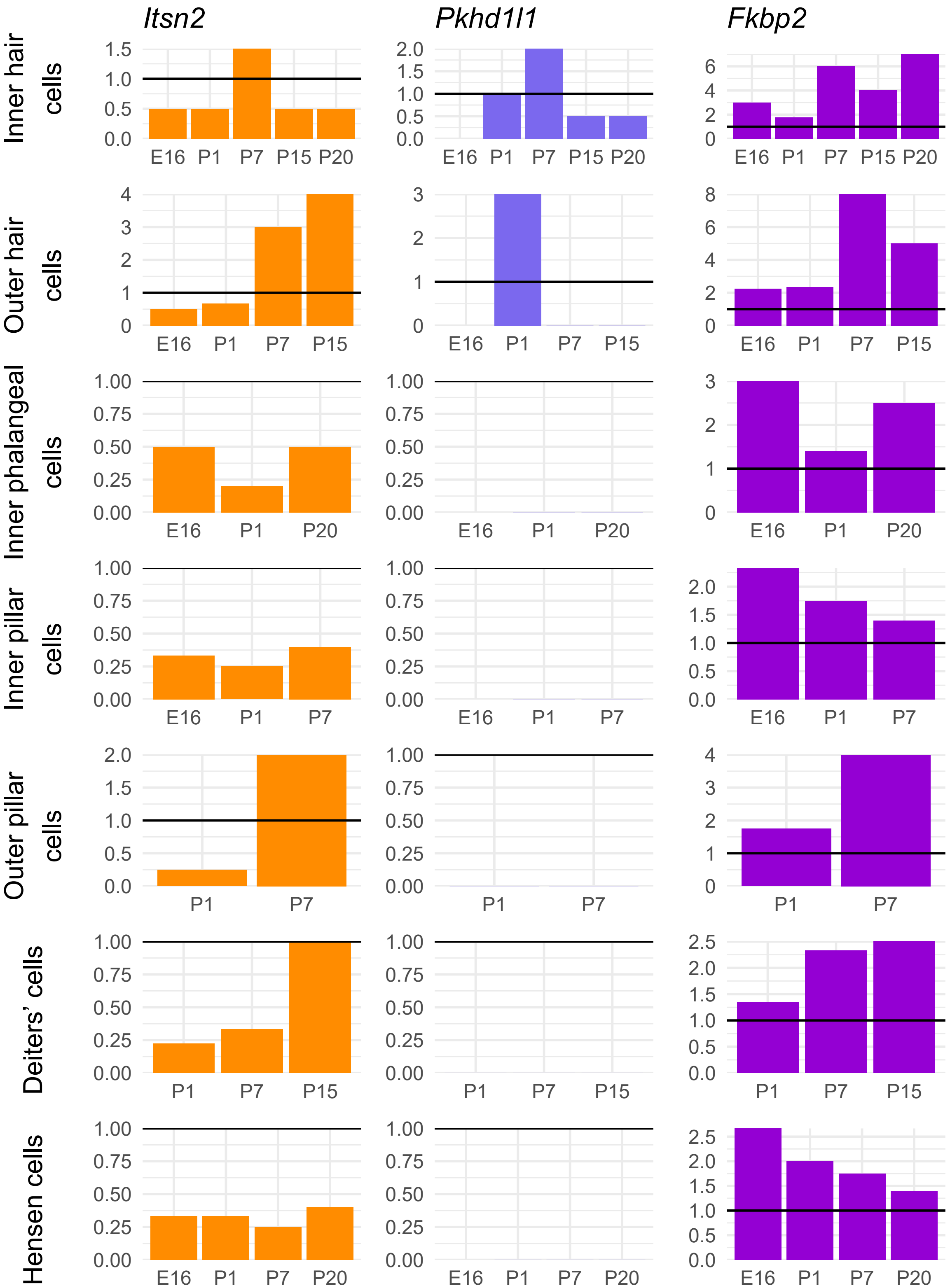
Itsn2
Pkhd1l1
Fkbp2
Inner hair cells
Outer hair cells
Inner phalangeal cells
Inner pillar cells
Outer pillar cells
Deiters' cells
Hensen cells
E16
P1
P7
P15
P20

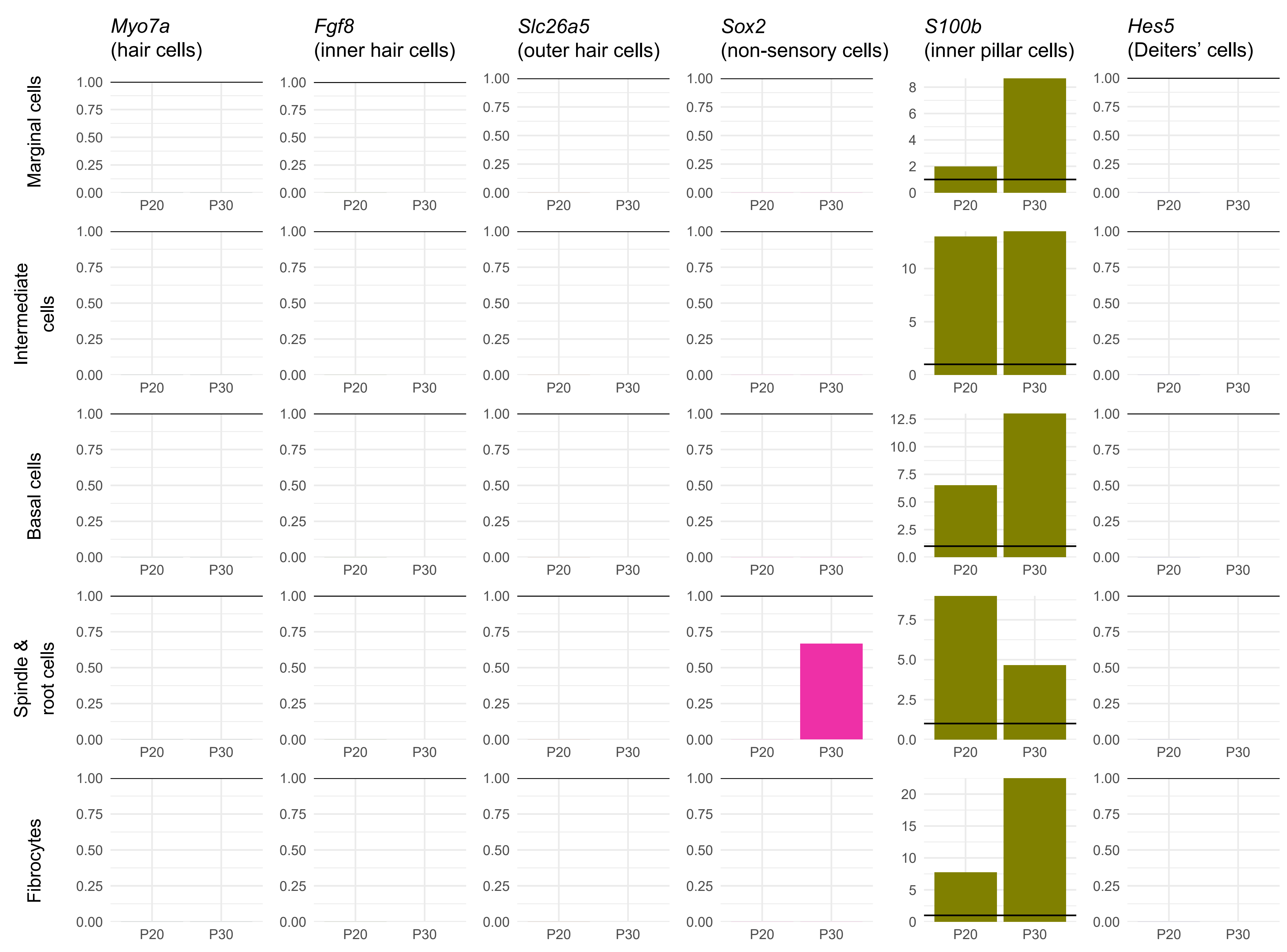

Myo7a
(hair cells)
Fgf8
(inner hair cells)
Slc26a5
(outer hair cells)
Sox2
(non-sensory cells)
S100b
(inner pillar cells)
Hes5
(Deiters' cells)
Marginal cells
Intermediate cells
Basal cells
Spindle & root cells
Fibrocytes
P20
P30
0.00
0.25
0.50
0.75
1.00

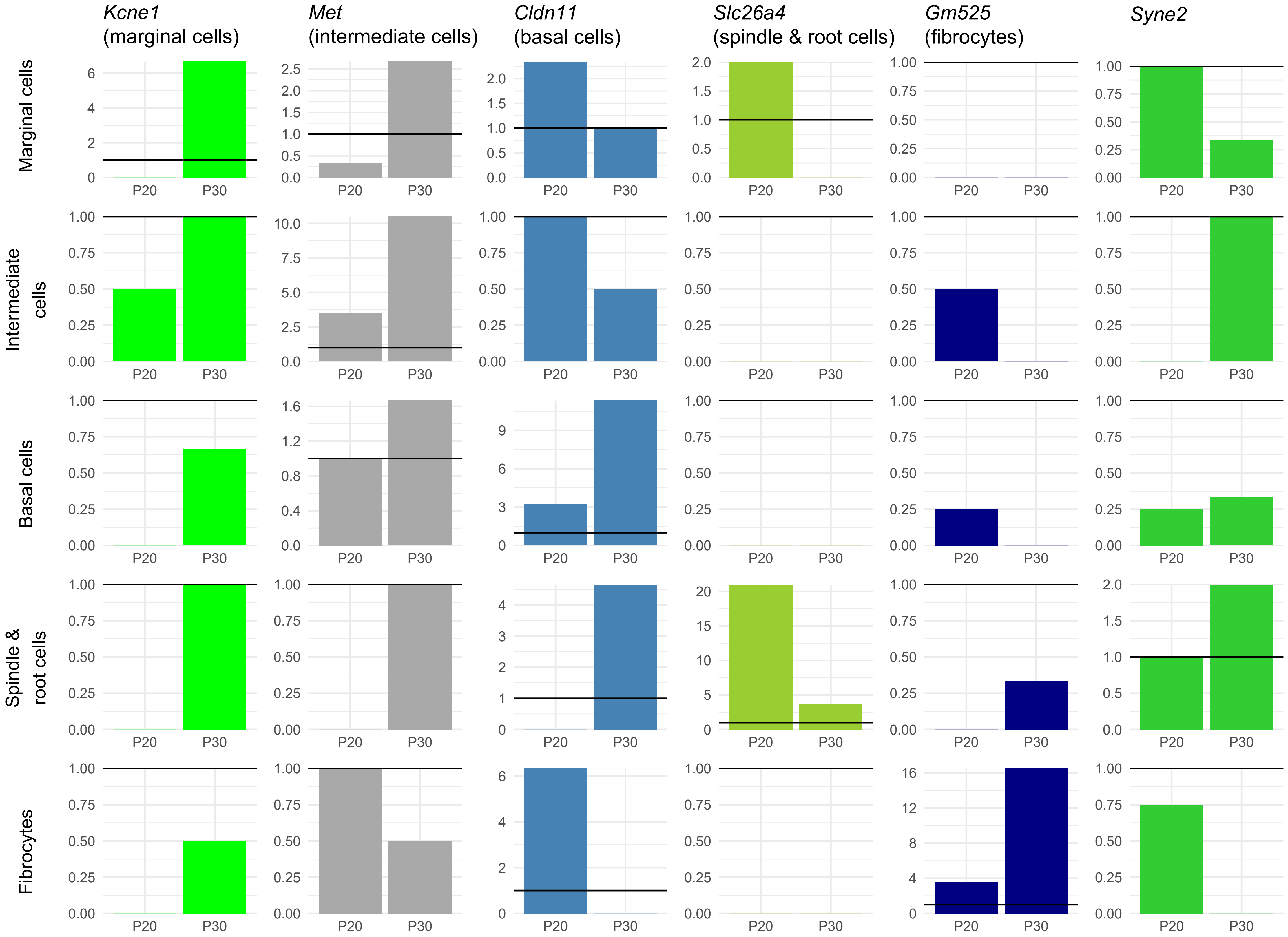

Kcne1 (marginal cells)
Met (intermediate cells)
Cldn11 (basal cells)
Slc26a4 (spindle & root cells)
Gm525 (fibrocytes)
Syne2
Marginal cells
Intermediate cells
Basal cells
Spindle & root cells
Fibrocytes
P20
P30

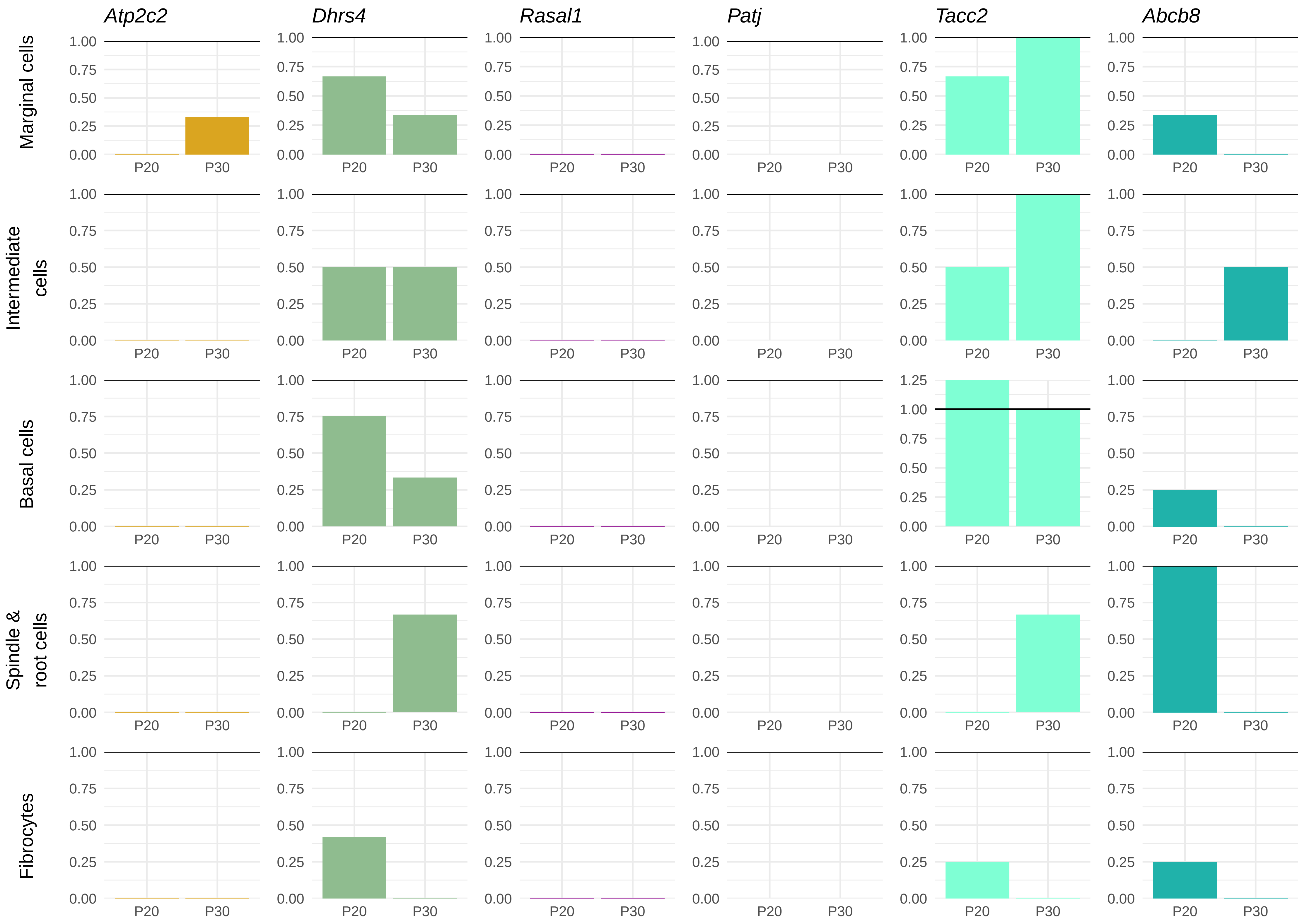
Atp2c2
Dhrs4
Rasal1
Patj
Tacc2
Abcb8
Marginal cells
Intermediate cells
Basal cells
Spindle & root cells
Fibrocytes
0.00
0.25
0.50
0.75
1.00
1.25
P20
P30

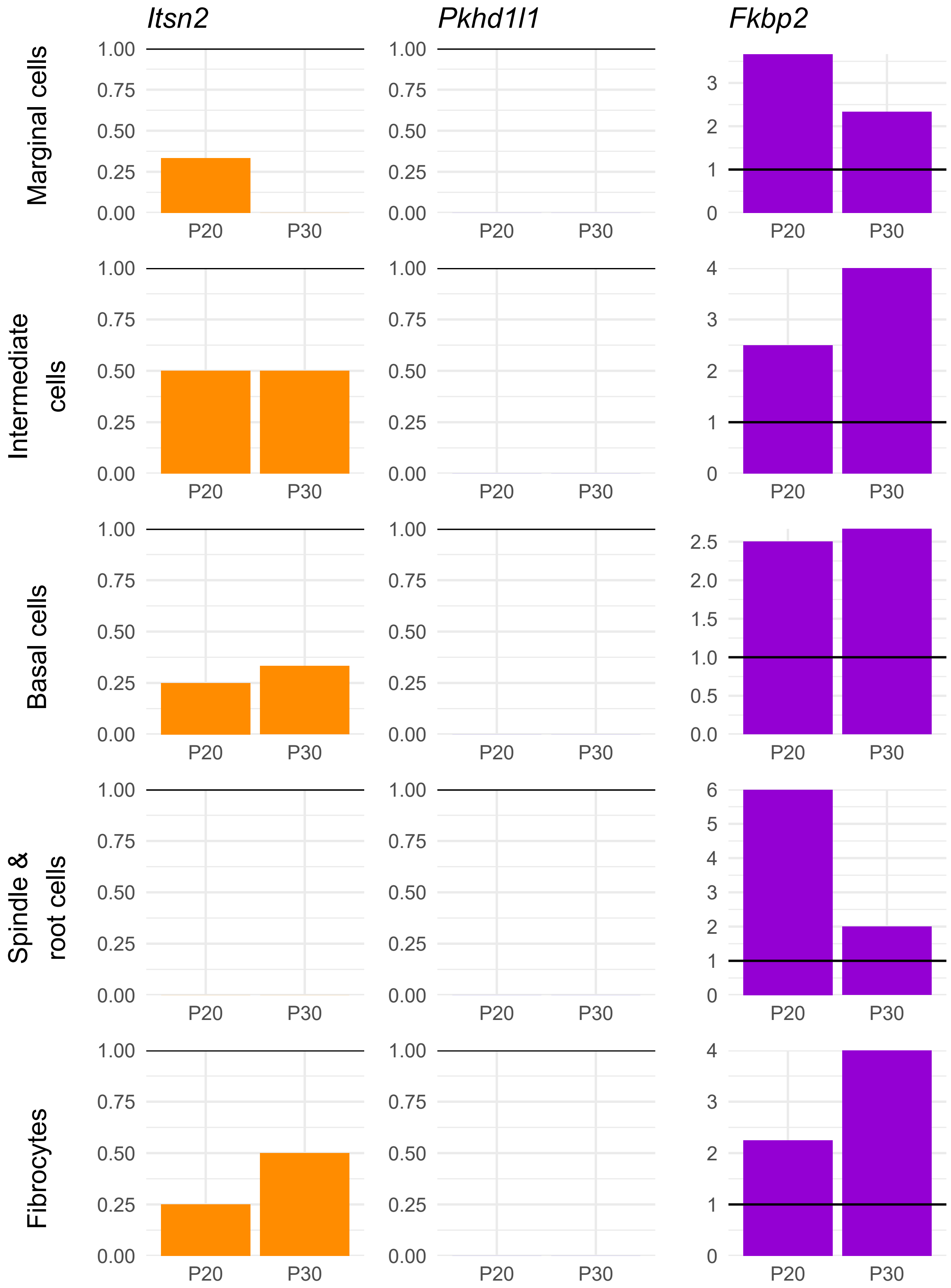

Itsn2
Pkhd1l1
Fkbp2
Marginal cells
Intermediate cells
Basal cells
Spindle & root cells
Fibrocytes
P20
P30

**Suppl. Figure 5.** Plots of gene expression in different cell types of the mouse inner ear, from single cell RNAseq data. These are genes identified in 3 cohorts (MUSC, TwinsUK and UK Biobank (Lewis et al. 2022)). Expression was normalised to *Hprt* (represented by a horizontal line at y=1 on each plot). Marker genes have been included for comparison (*Myo7a* (hair cells), *Fgf8* (inner hair cells), *Slc26a5* (outer hair cells), *Sox2* (non-sensory cells), *S100b* (inner pillar cells), *Hes5* (Deiters' cells), *Kcne1* (marginal cells), *Met* (intermediate cells), *Cldn11* (basal cells), *Slc26a4* (spindle and root cells), *Gm525* (fibrocytes)). Datasets were obtained from the gEAR (http://umgear.org) (Kolla et al. 2020; Korrapati et al. 2019; Ranum et al. 2019; Xue et al. 2021).

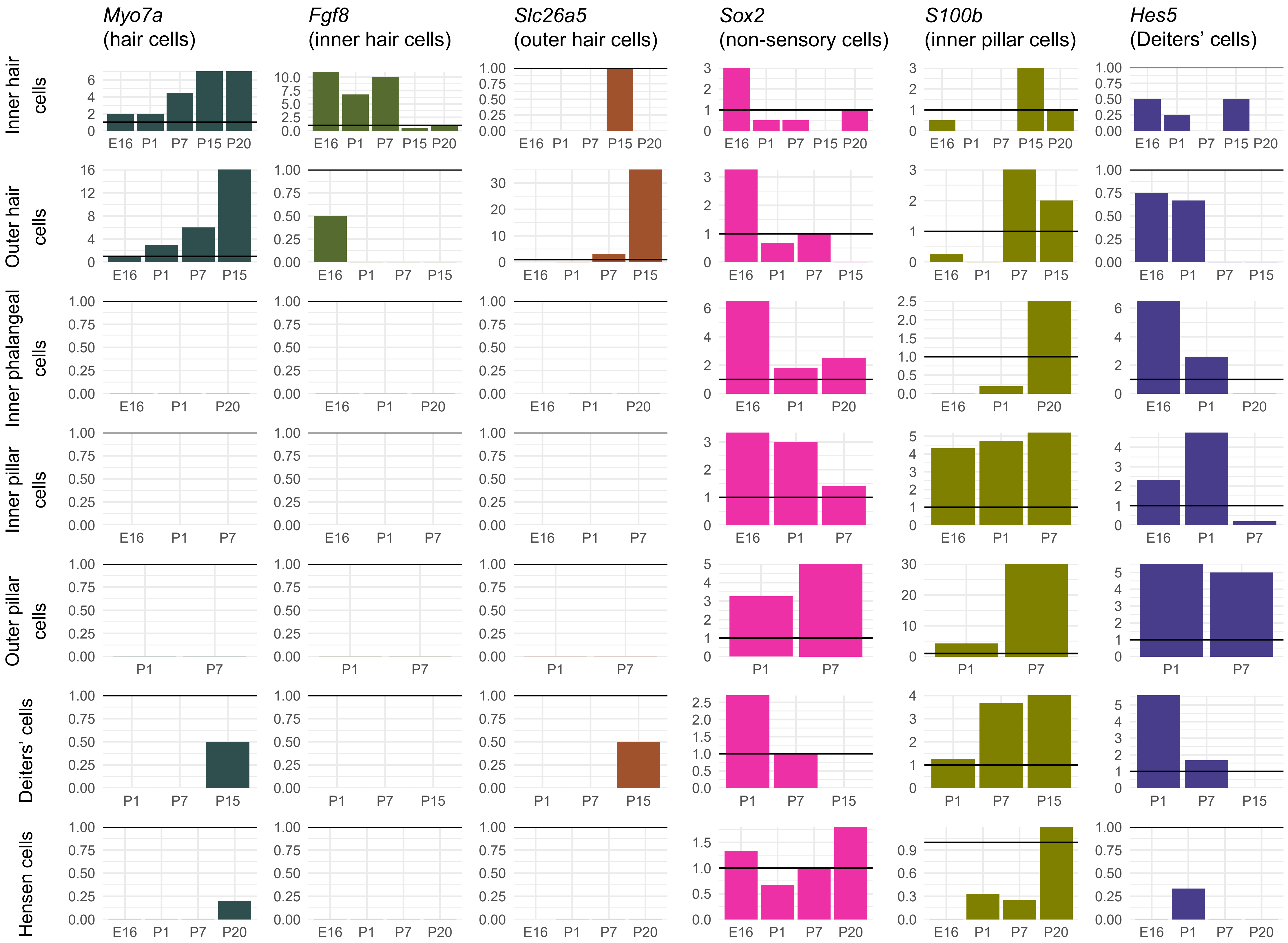

Myo7a (hair cells)
Fgf8 (inner hair cells)
Slc26a5 (outer hair cells)
Sox2 (non-sensory cells)
S100b (inner pillar cells)
Hes5 (Deiters' cells)
Inner hair cells
Outer hair cells
Inner phalangeal cells
Inner pillar cells
Outer pillar cells
Deiters' cells
Hensen cells
E16
P1
P7
P15
P20

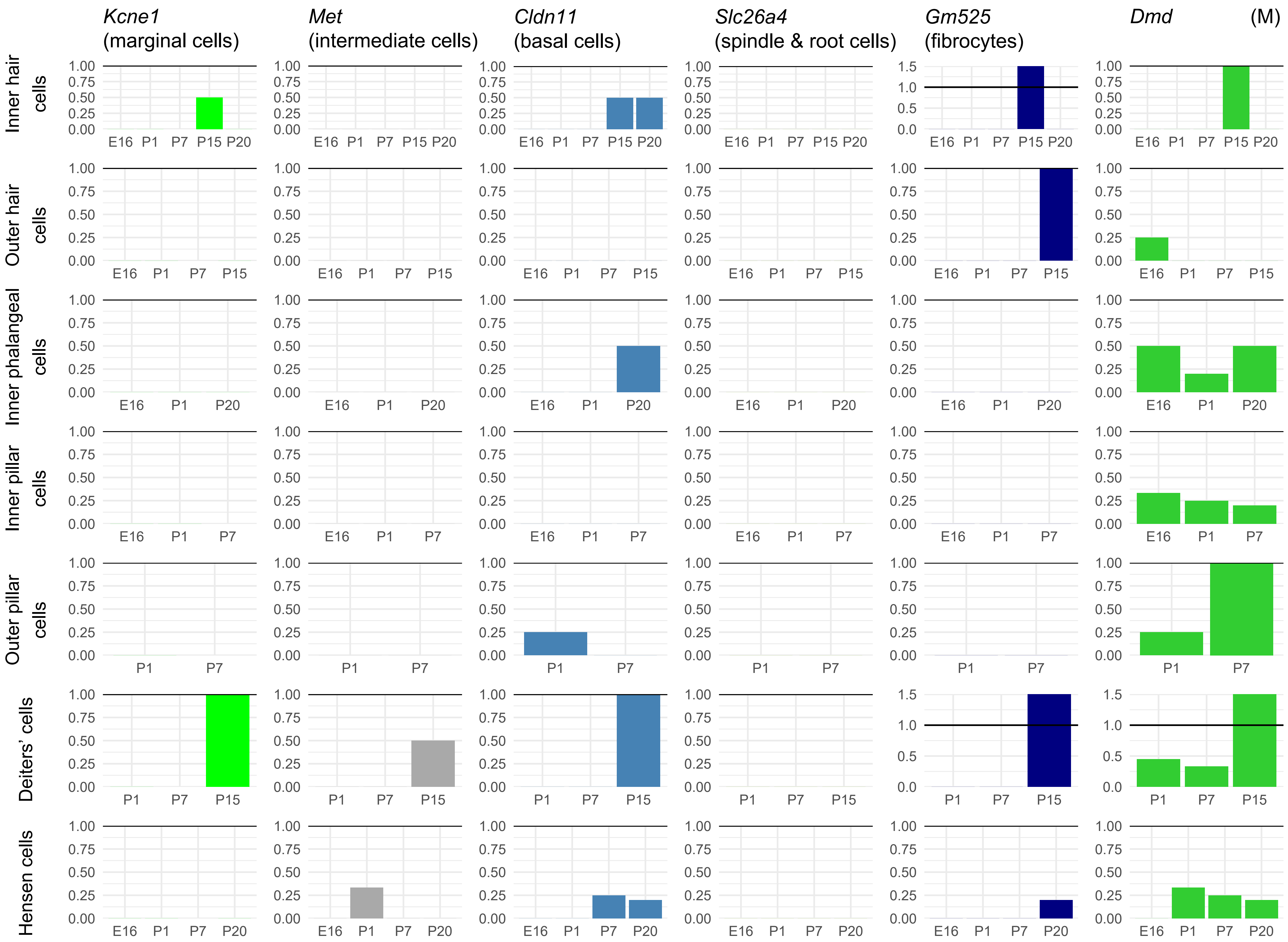

(M)
Kcne1 (marginal cells)
Met (intermediate cells)
Cldn11 (basal cells)
Slc26a4 (spindle & root cells)
Gm525 (fibrocytes)
Dmd
Inner hair cells
Outer hair cells
Inner phalangeal cells
Inner pillar cells
Outer pillar cells
Deiters' cells
Hensen cells
E16
P1
P7
P15
P20
0.00
0.25
0.50
0.75
1.00
0.0
0.5
1.0
1.5

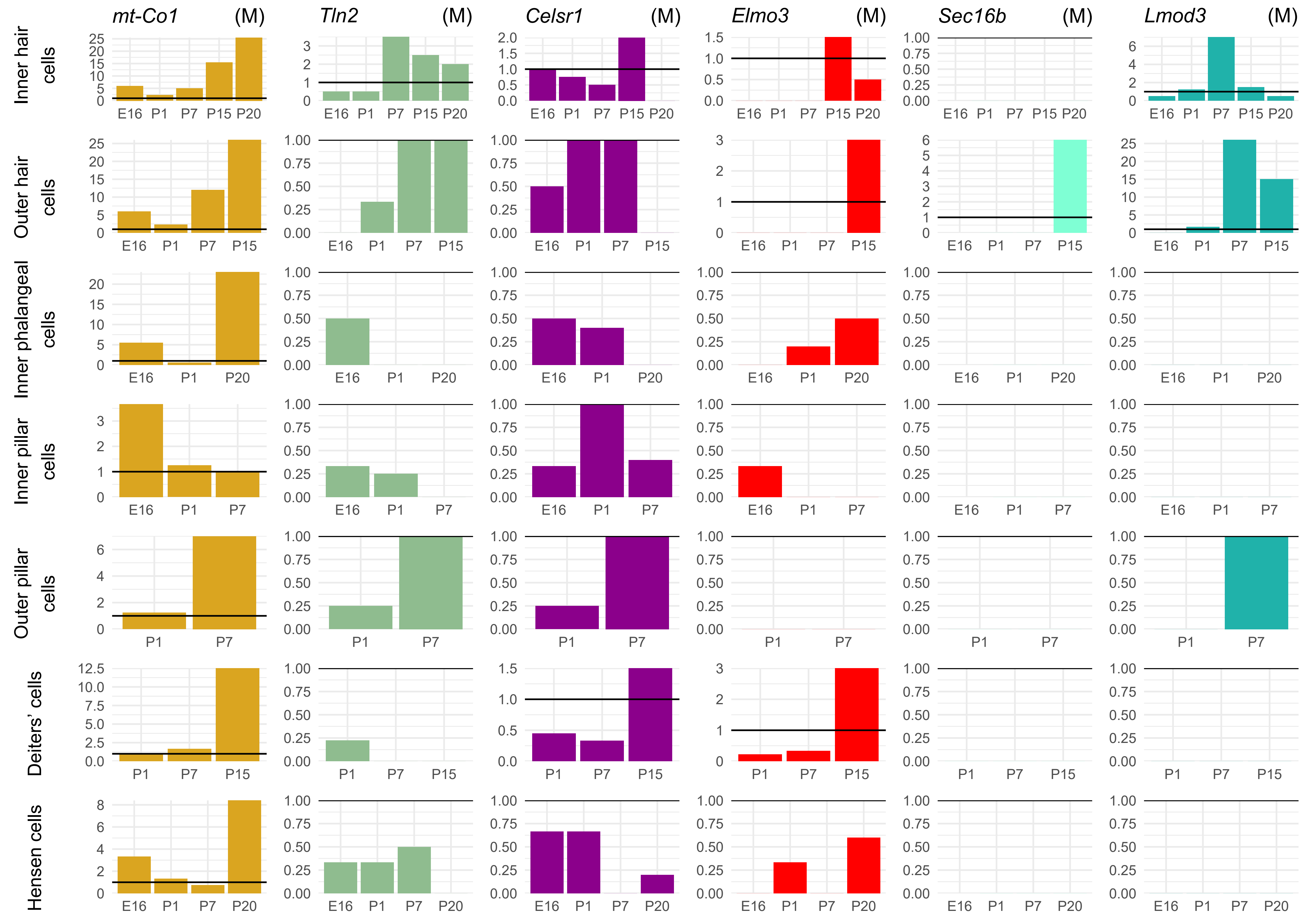

mt-Co1 (M)
Tln2 (M)
Celsr1 (M)
Elmo3 (M)
Sec16b (M)
Lmod3 (M)
Inner hair cells
Outer hair cells
Inner phalangeal cells
Inner pillar cells
Outer pillar cells
Deiters' cells
Hensen cells
E16
P1
P7
P15
P20

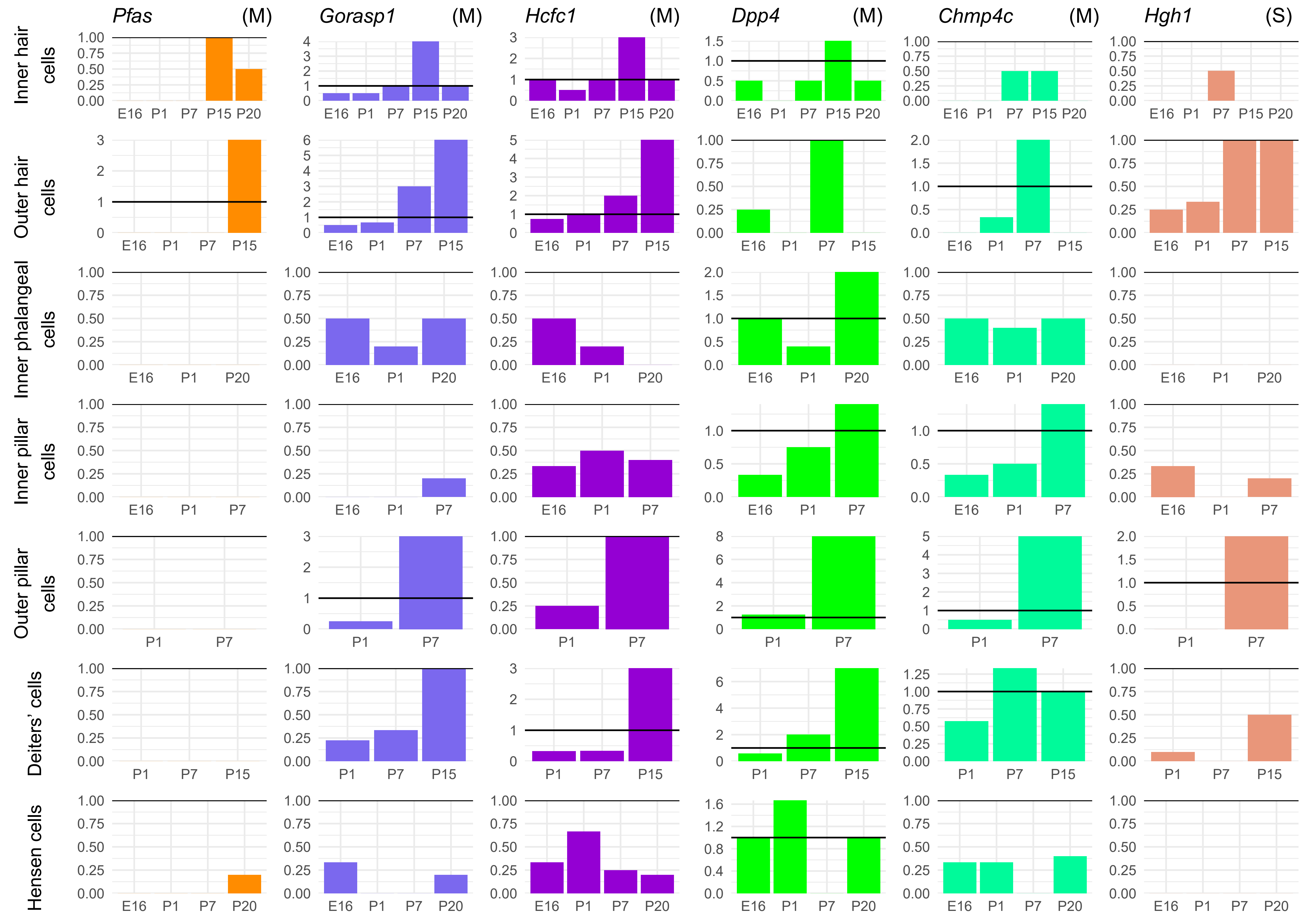

Pfas (M)
Gorasp1 (M)
Hcfc1 (M)
Dpp4 (M)
Chmp4c (M)
Hgh1 (S)
Inner hair cells
Outer hair cells
Inner phalangeal cells
Inner pillar cells
Outer pillar cells
Deiters' cells
Hensen cells
E16
P1
P7
P15
P20

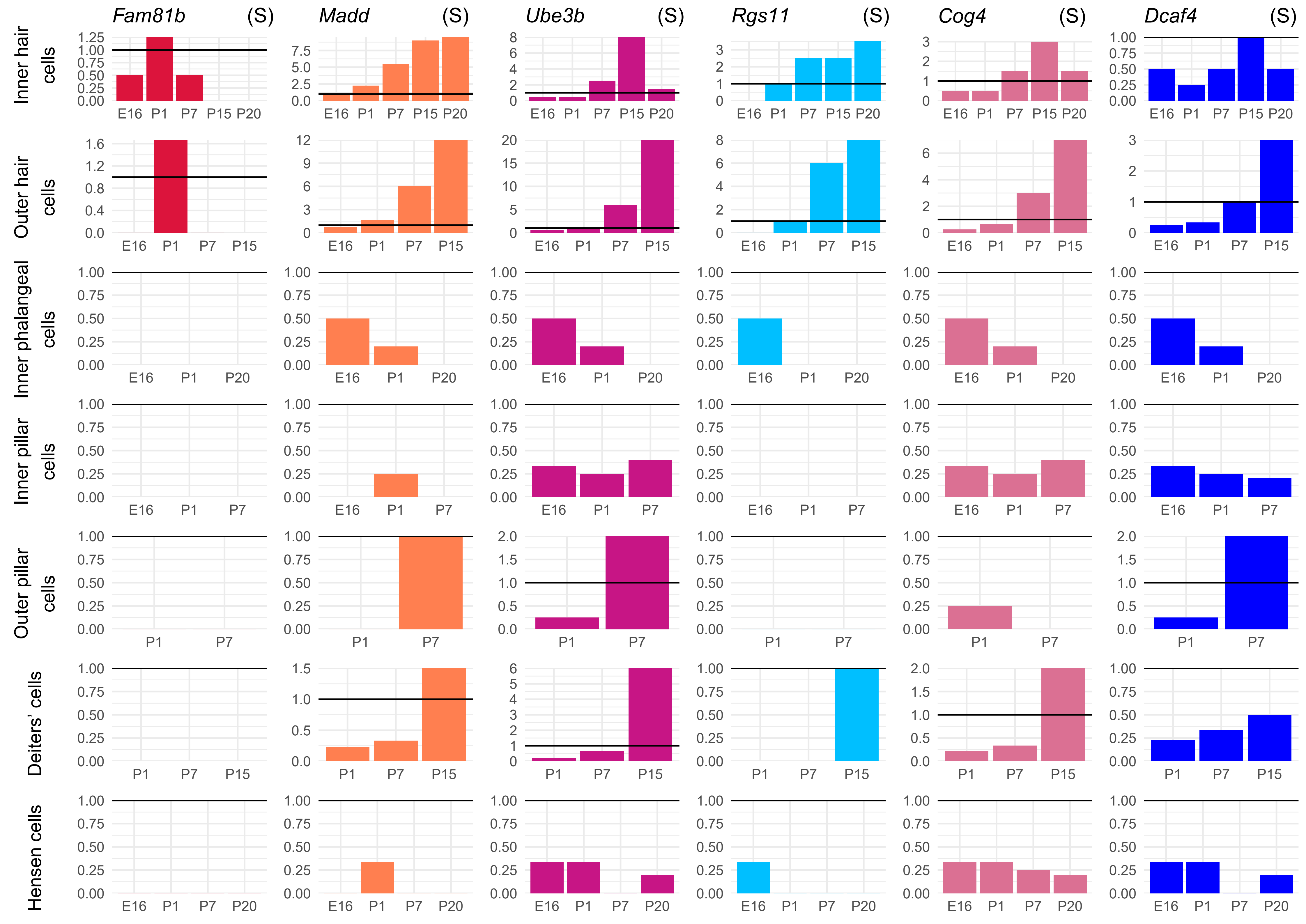

Fam81b (S)
Madd (S)
Ube3b (S)
Rgs11 (S)
Cog4 (S)
Dcaf4 (S)
Inner hair cells
Outer hair cells
Inner phalangeal cells
Inner pillar cells
Outer pillar cells
Deiters' cells
Hensen cells
E16 P1 P7 P15 P20

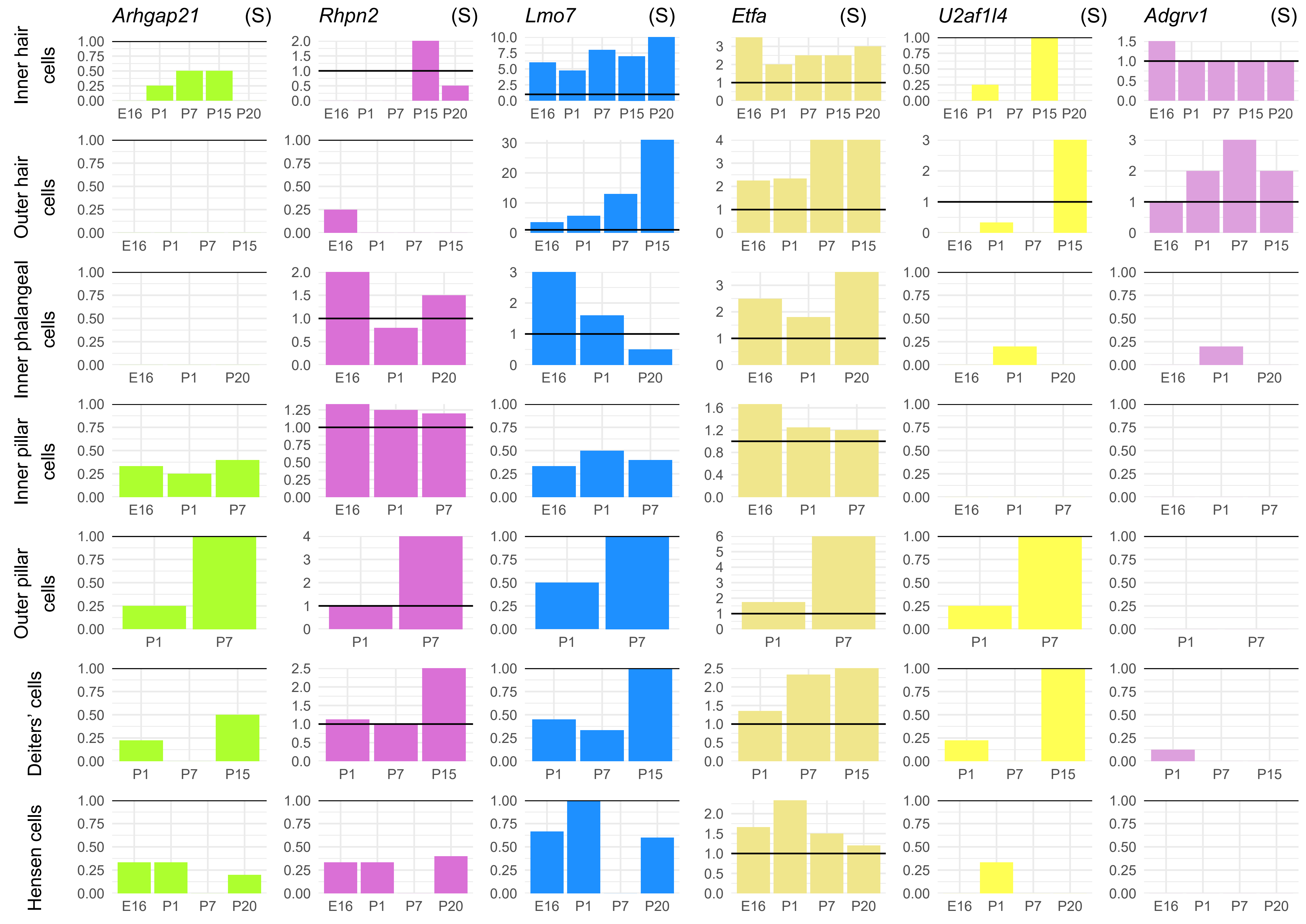
Arhgap21 (S)
Rhpn2 (S)
Lmo7 (S)
Etfa (S)
U2af1l4 (S)
Adgrv1 (S)
Inner hair cells
Outer hair cells
Inner phalangeal cells
Inner pillar cells
Outer pillar cells
Deiters' cells
Hensen cells
E16
P1
P7
P15
P20

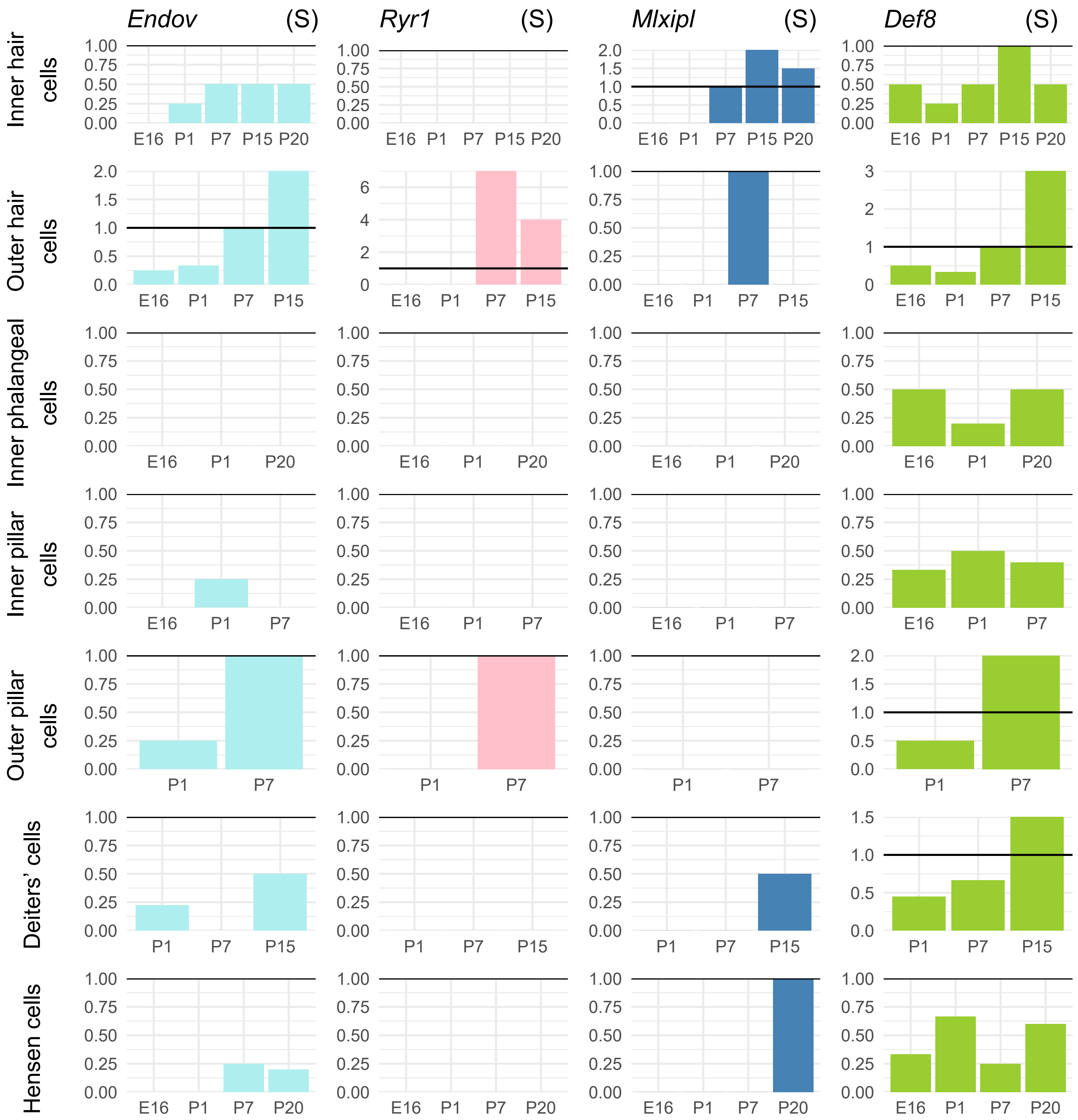

Endov (S)
Ryr1 (S)
Mlxipl (S)
Def8 (S)
Inner hair cells
Outer hair cells
Inner phalangeal cells
Inner pillar cells
Outer pillar cells
Deiters' cells
Hensen cells
E16 P1 P7 P15 P20

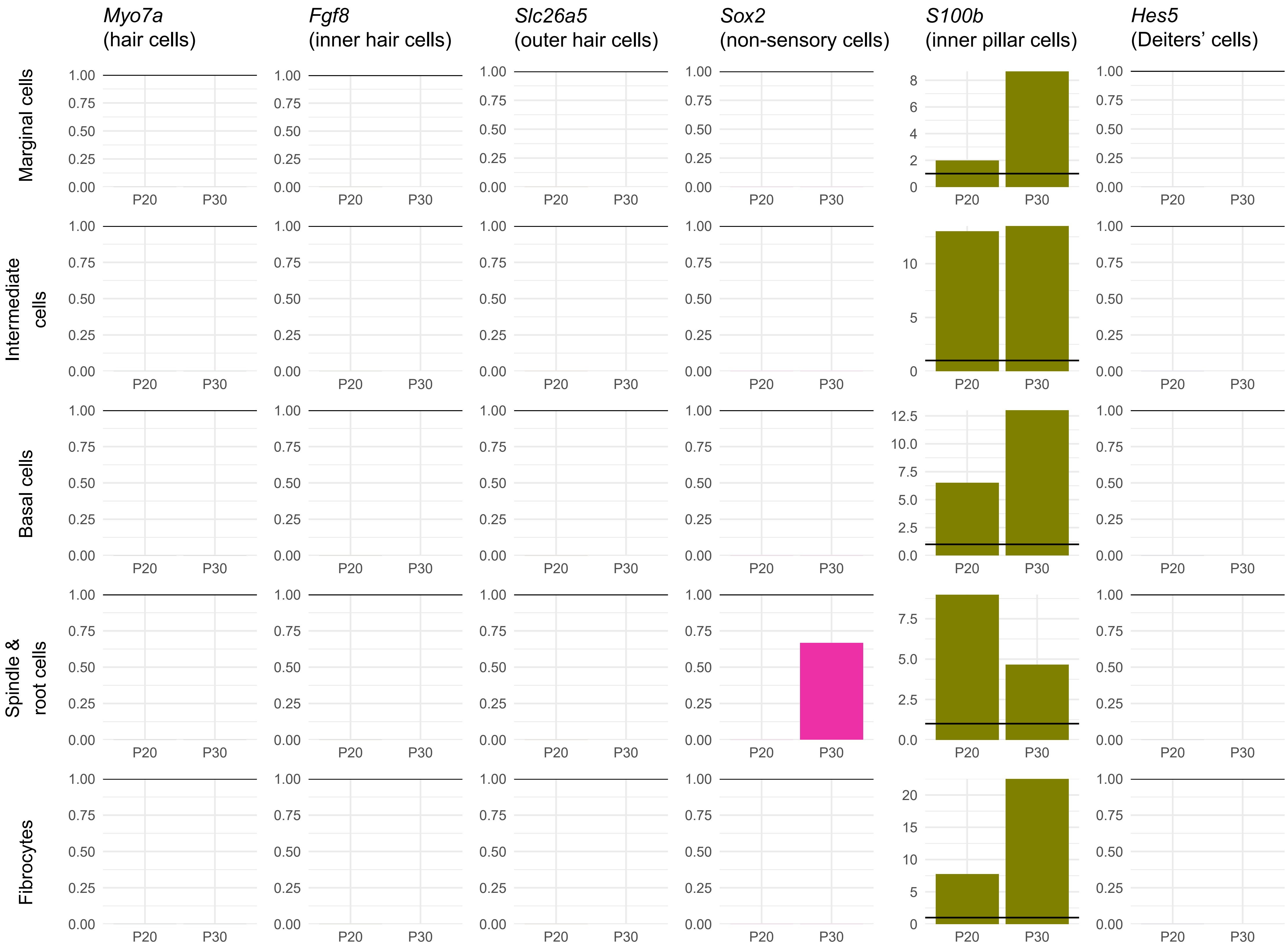

Myo7a (hair cells)
Fgf8 (inner hair cells)
Slc26a5 (outer hair cells)
Sox2 (non-sensory cells)
S100b (inner pillar cells)
Hes5 (Deiters' cells)
Marginal cells
Intermediate cells
Basal cells
Spindle & root cells
Fibrocytes
P20
P30

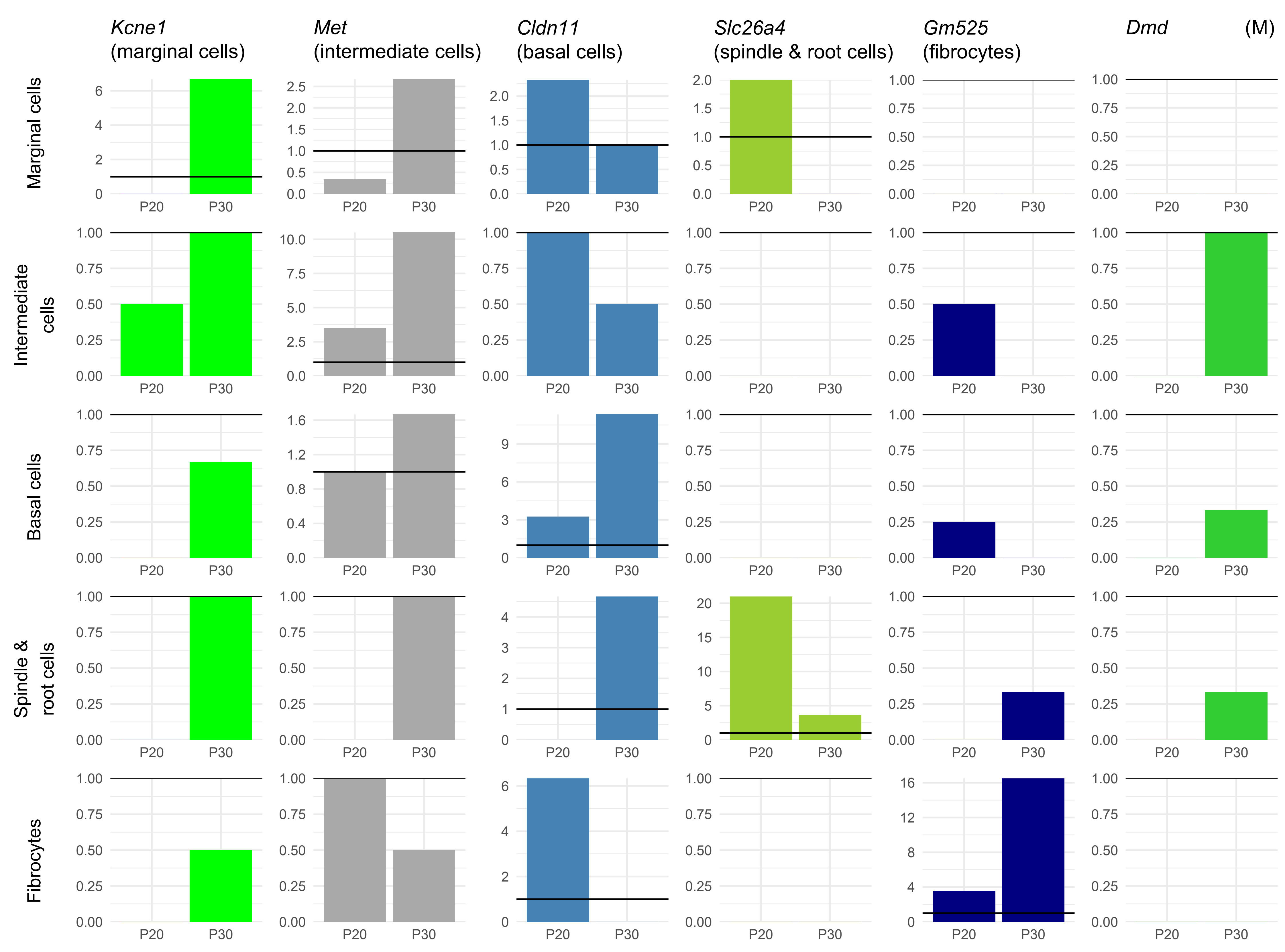

Kcne1
(marginal cells)
Met
(intermediate cells)
Cldn11
(basal cells)
Slc26a4
(spindle & root cells)
Gm525
(fibrocytes)
Dmd
(M)
Marginal cells
Intermediate cells
Basal cells
Spindle & root cells
Fibrocytes
P20
P30

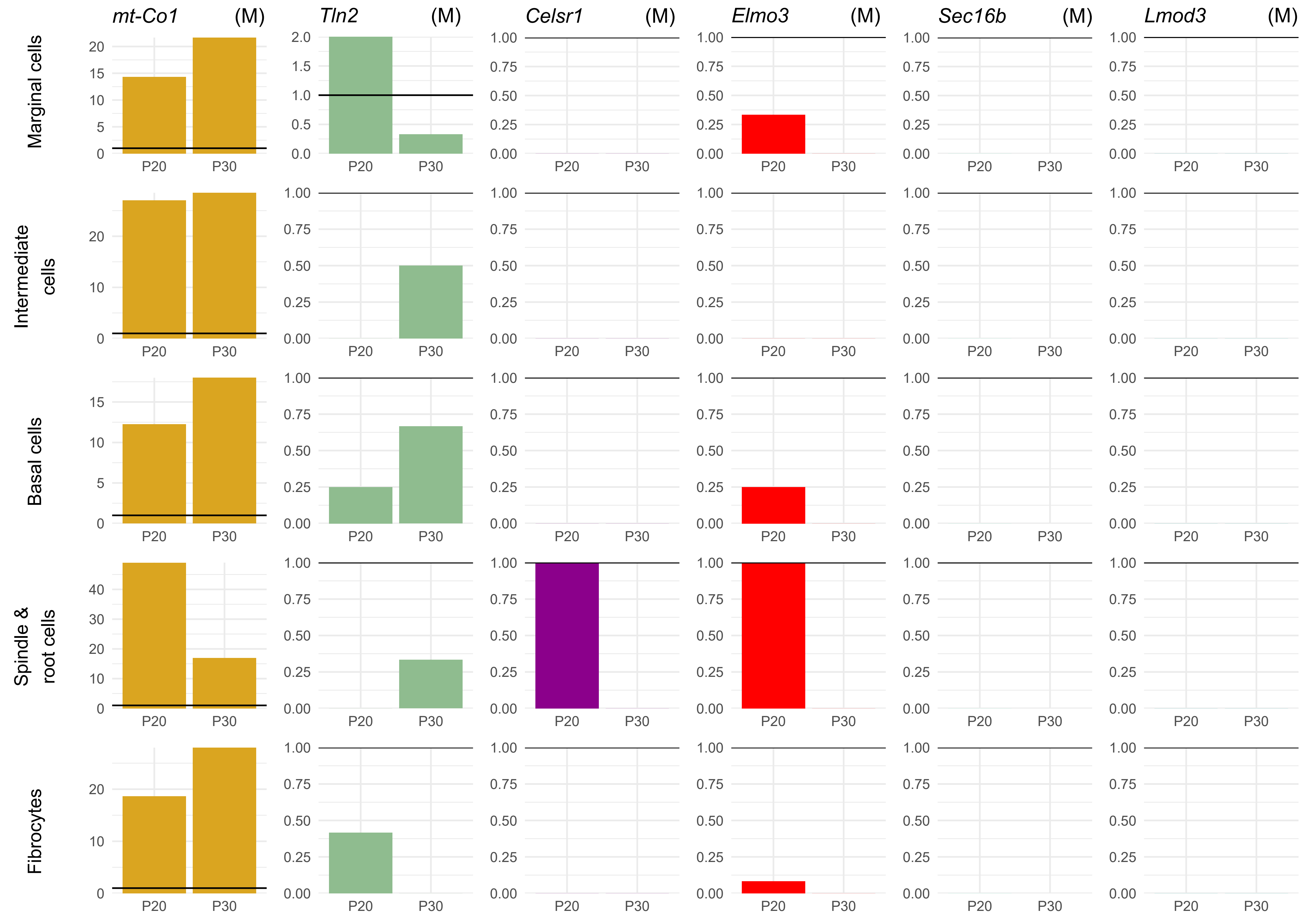

mt-Co1 (M)
Tln2 (M)
Celsr1 (M)
Elmo3 (M)
Sec16b (M)
Lmod3 (M)
Marginal cells
Intermediate cells
Basal cells
Spindle & root cells
Fibrocytes
P20
P30

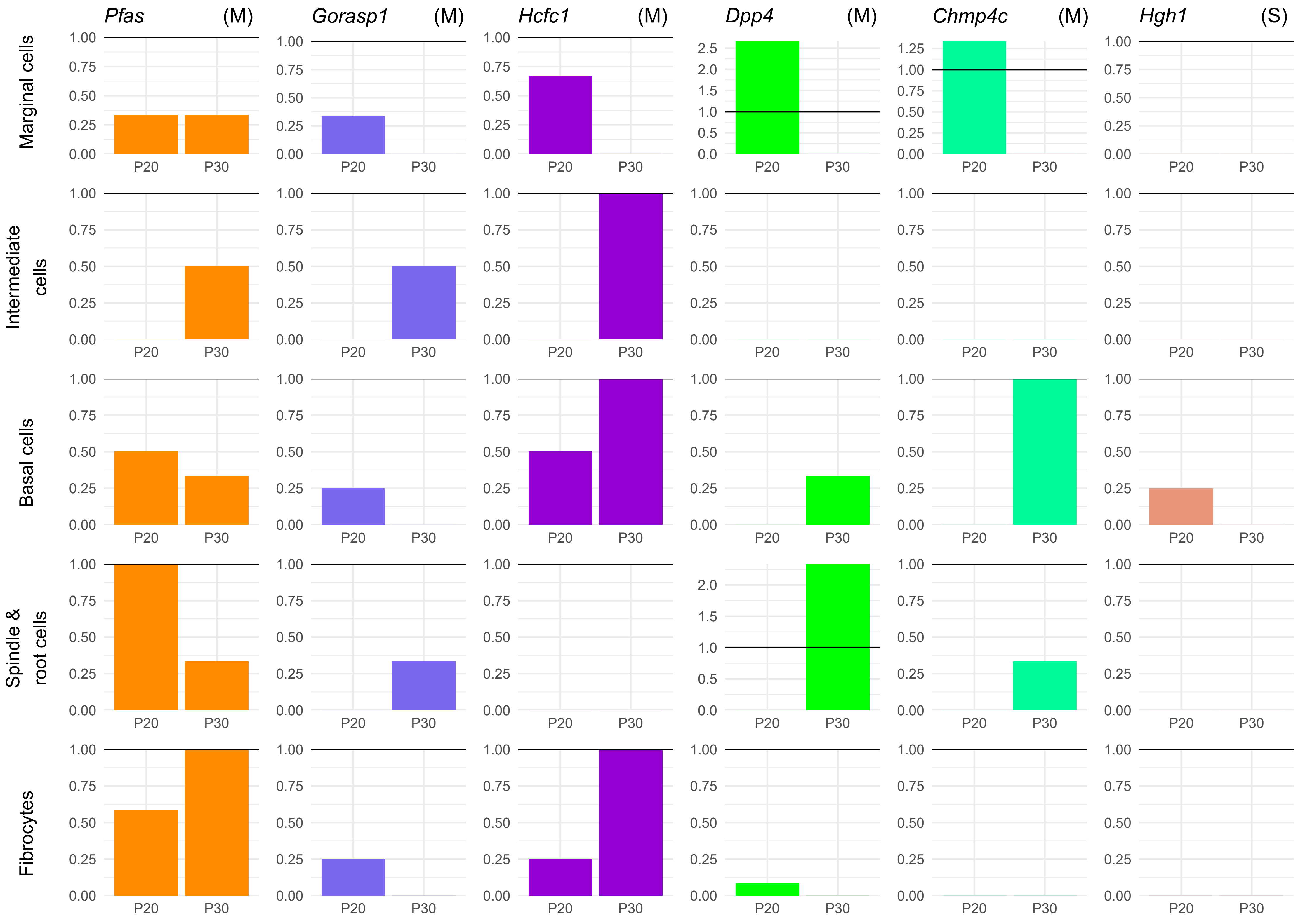
Pfas (M)
Gorasp1 (M)
Hcfc1 (M)
Dpp4 (M)
Chmp4c (M)
Hgh1 (S)
Marginal cells
Intermediate cells
Basal cells
Spindle & root cells
Fibrocytes
P20
P30

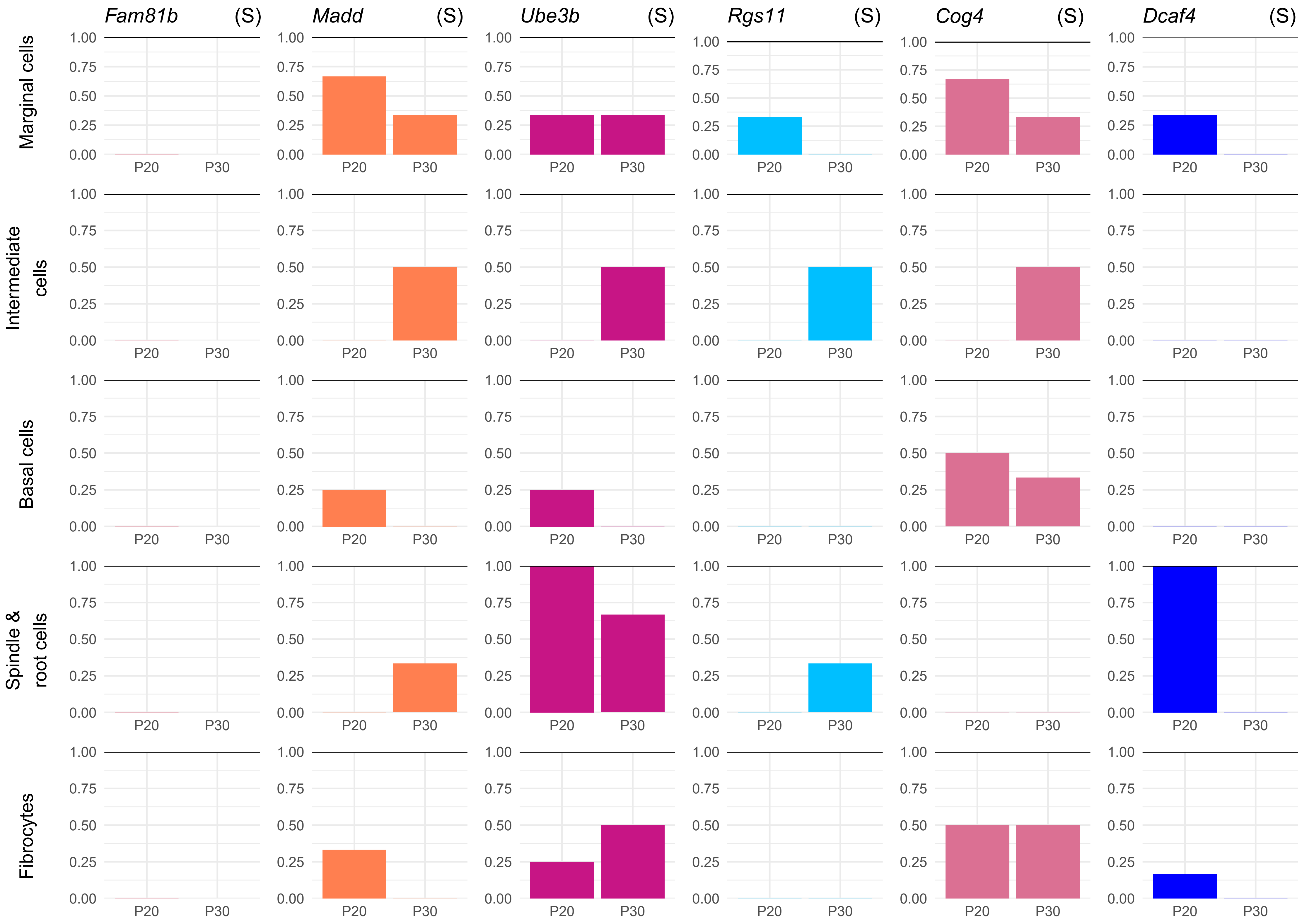

Fam81b (S)
Madd (S)
Ube3b (S)
Rgs11 (S)
Cog4 (S)
Dcaf4 (S)
Marginal cells
Intermediate cells
Basal cells
Spindle & root cells
Fibrocytes
1.00
0.75
0.50
0.25
0.00
P20
P30

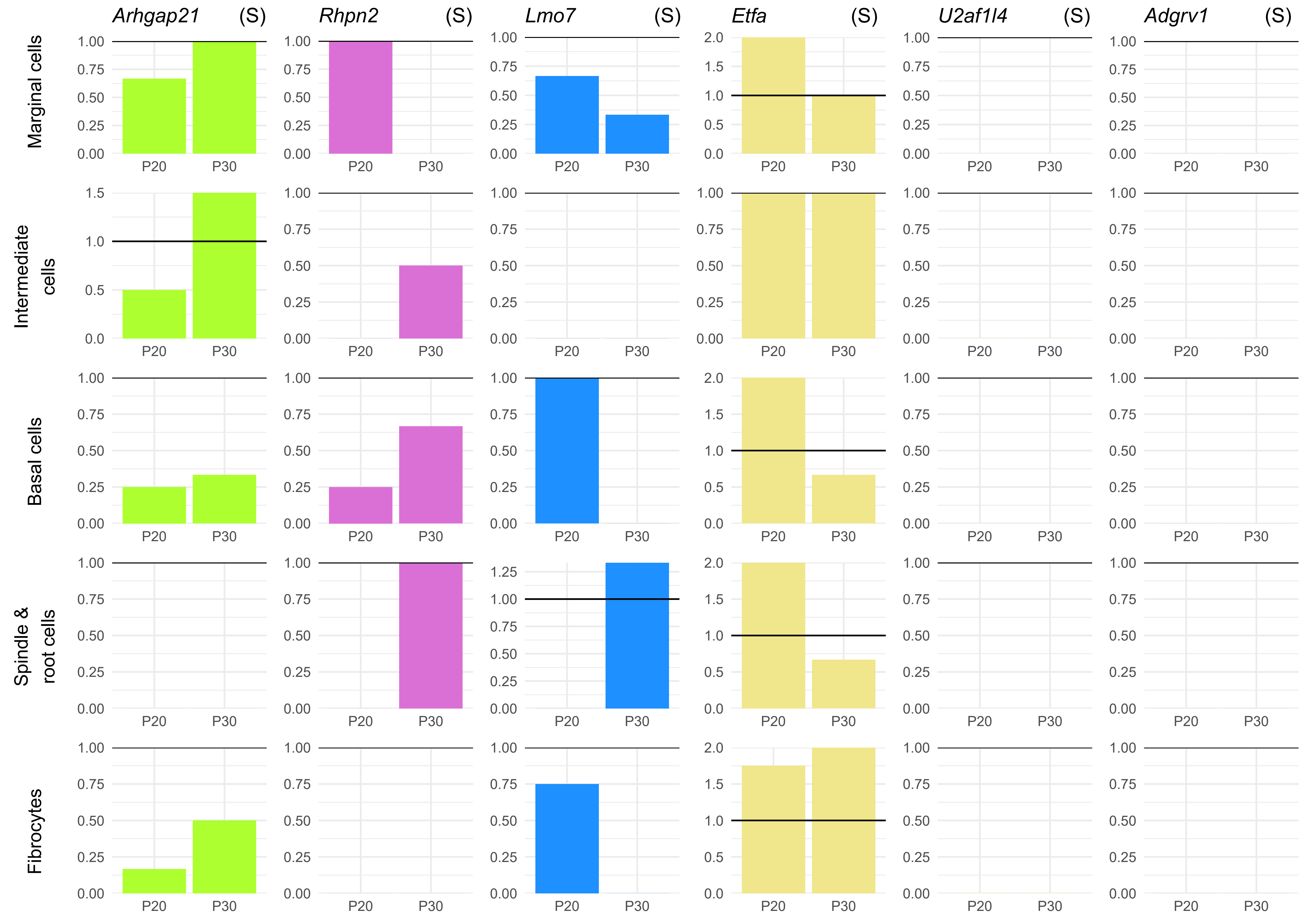

Arhgap21 (S)
Rhpn2 (S)
Lmo7 (S)
Etfa (S)
U2af1l4 (S)
Adgrv1 (S)
Marginal cells
Intermediate cells
Basal cells
Spindle & root cells
Fibrocytes
P20
P30

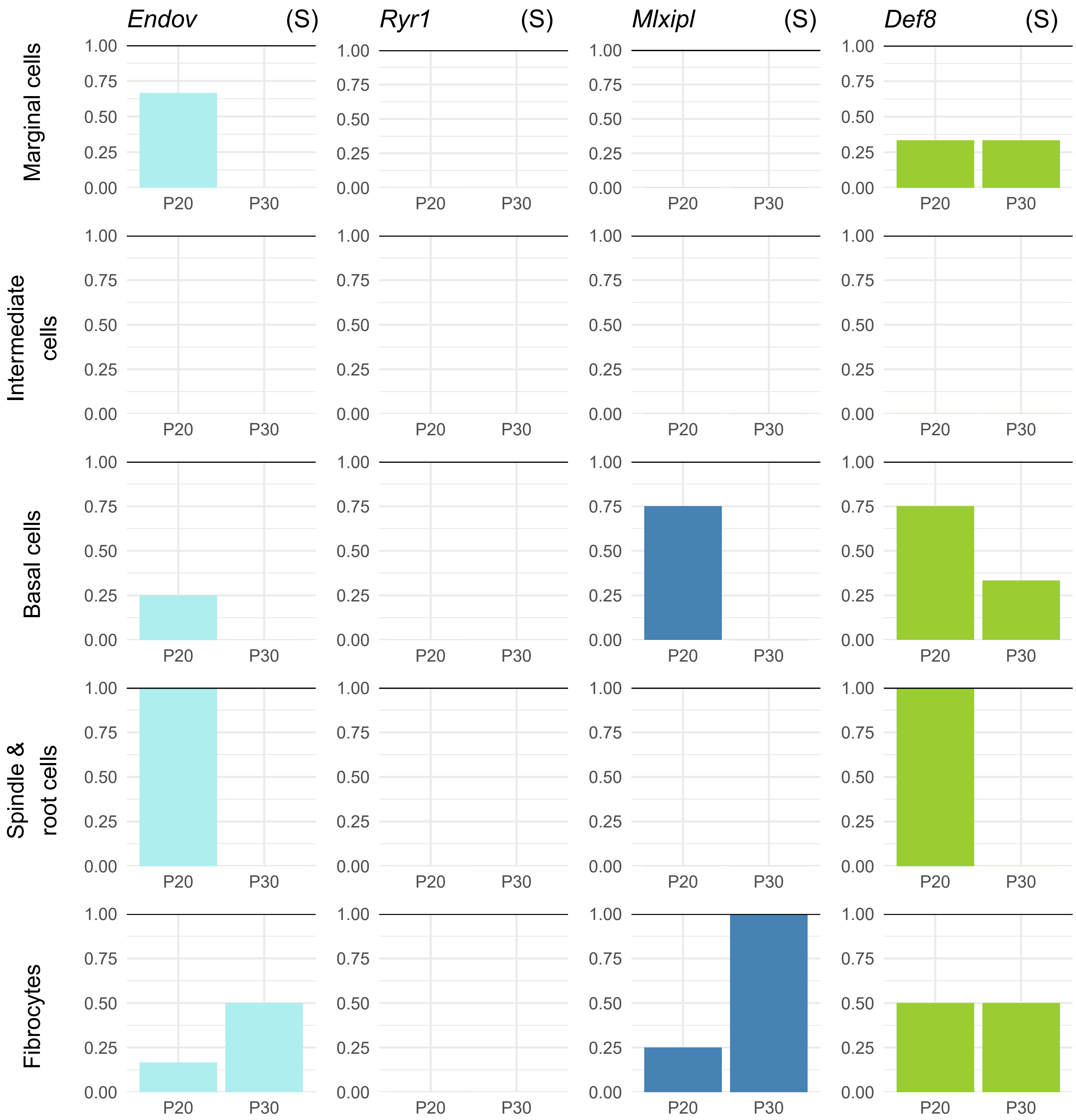
Endov
(S)
Ryr1
(S)
Mlxipl
(S)
Def8
(S)
Marginal cells
Intermediate cells
Basal cells
Spindle & root cells
Fibrocytes
1.00
0.75
0.50
0.25
0.00
P20
P30

**Suppl. Figure 6.** Plots of gene expression in different cell types of the mouse inner ear, from single cell RNAseq data. These are genes linked only to phenotypic subtypes in this study (M = Metabolic, S = Sensory). Expression was normalised to *Hprt* (represented by a horizontal line at y=1 on each plot). Marker genes have been included for comparison (*Myo7a* (hair cells), *Fgf8* (inner hair cells), *Slc26a5* (outer hair cells), *Sox2* (non-sensory cells), *S100b* (inner pillar cells), *Hes5* (Deiters' cells), *Kcne1* (marginal cells), *Met* (intermediate cells), *Cldn11* (basal cells), *Slc26a4* (spindle and root cells), *Gm525* (fibrocytes)). Datasets were obtained from the gEAR (http://umgear.org) (Kolla et al. 2020; Korrapati et al. 2019; Ranum et al. 2019; Xue et al. 2021).

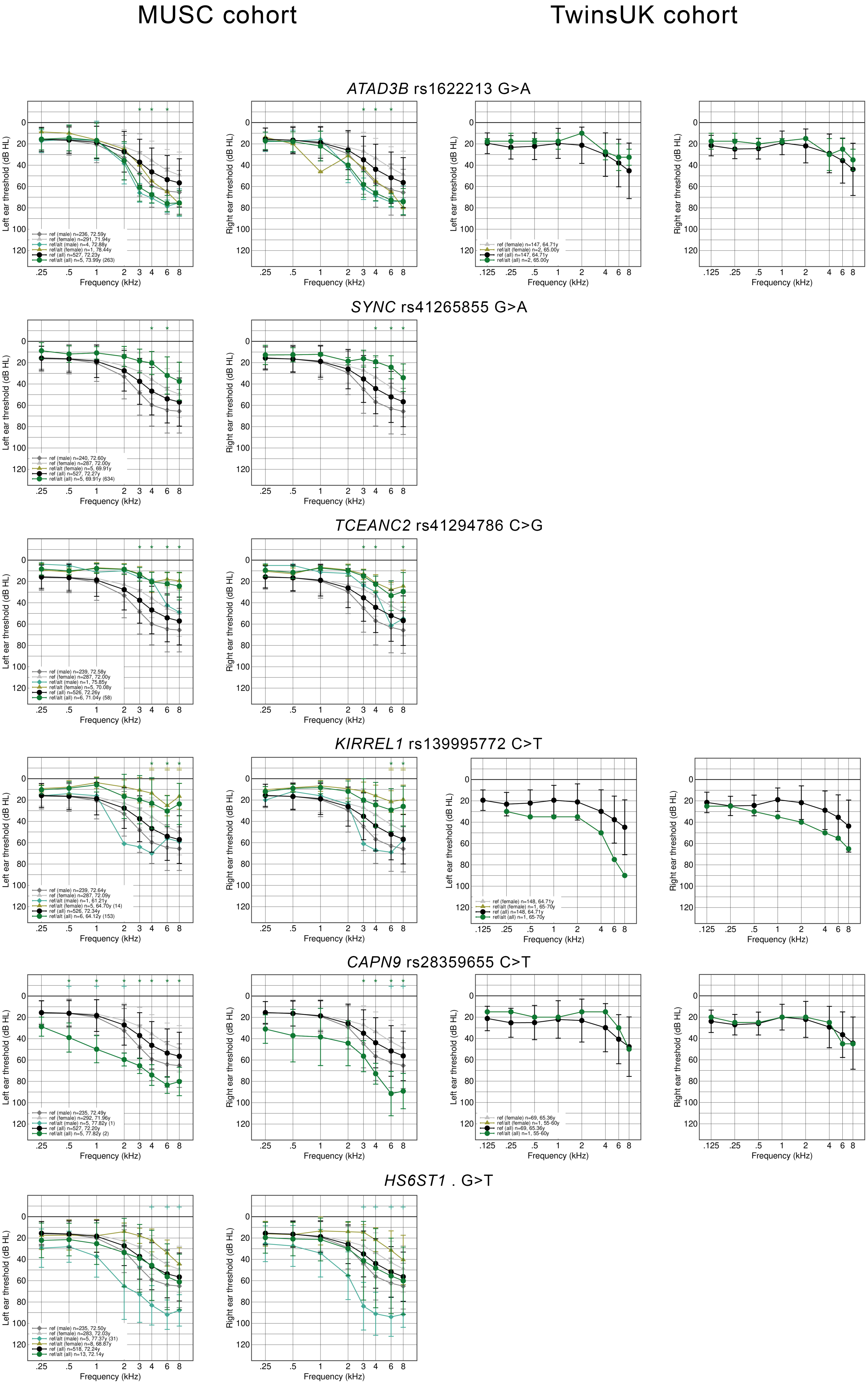
MUSC cohort
TwinsUK cohort
ATAD3B rs1622213 G>A
ref (male) n=236, 72.59y
ref (female) n=291, 71.94y
ref/alt (male) n=4, 72.88y
ref/alt (female) n=1, 78.44y
ref (all) n=527, 72.23y
ref/alt (all) n=5, 73.99y (263)
ref (female) n=147, 64.71y
ref/alt (female) n=2, 65.00y
ref (all) n=147, 64.71y
ref/alt (all) n=2, 65.00y
SYNC rs41265855 G>A
ref (male) n=240, 72.60y
ref (female) n=287, 72.00y
ref/alt (female) n=5, 69.91y
ref (all) n=527, 72.27y
ref/alt (all) n=5, 69.91y (634)
TCEANC2 rs41294786 C>G
ref (male) n=239, 72.58y
ref (female) n=287, 72.00y
ref/alt (male) n=1, 75.85y
ref/alt (female) n=5, 70.08y
ref (all) n=526, 72.26y
ref/alt (all) n=6, 71.04y (58)
KIRREL1 rs139995772 C>T
ref (male) n=239, 72.64y
ref (female) n=287, 72.09y
ref/alt (male) n=1, 61.21y
ref/alt (female) n=5, 64.70y (14)
ref (all) n=526, 72.34y
ref/alt (all) n=6, 64.12y (153)
ref (female) n=148, 64.71y
ref/alt (female) n=1, 65-70y
ref (all) n=148, 64.71y
ref/alt (all) n=1, 65-70y
CAPN9 rs28359655 C>T
ref (male) n=235, 72.49y
ref (female) n=292, 71.96y
ref/alt (male) n=5, 77.82y (1)
ref (all) n=527, 72.20y
ref/alt (all) n=5, 77.82y (2)
ref (female) n=69, 65.36y
ref/alt (male) n=1, 55-60y
ref (all) n=69, 65.36y
ref/alt (all) n=1, 55-60y
HS6ST1 . G>T
ref (male) n=235, 72.50y
ref (female) n=283, 72.03y
ref/alt (male) n=5, 77.37y (31)
ref/alt (female) n=8, 68.87y
ref (all) n=518, 72.24y
ref/alt (all) n=13, 72.14y
Left ear threshold (dB HL)
Right ear threshold (dB HL)
Frequency (kHz)
0
20
40
60
80
100
120
.125
.25
.5
1
2
3
4
6
8
*

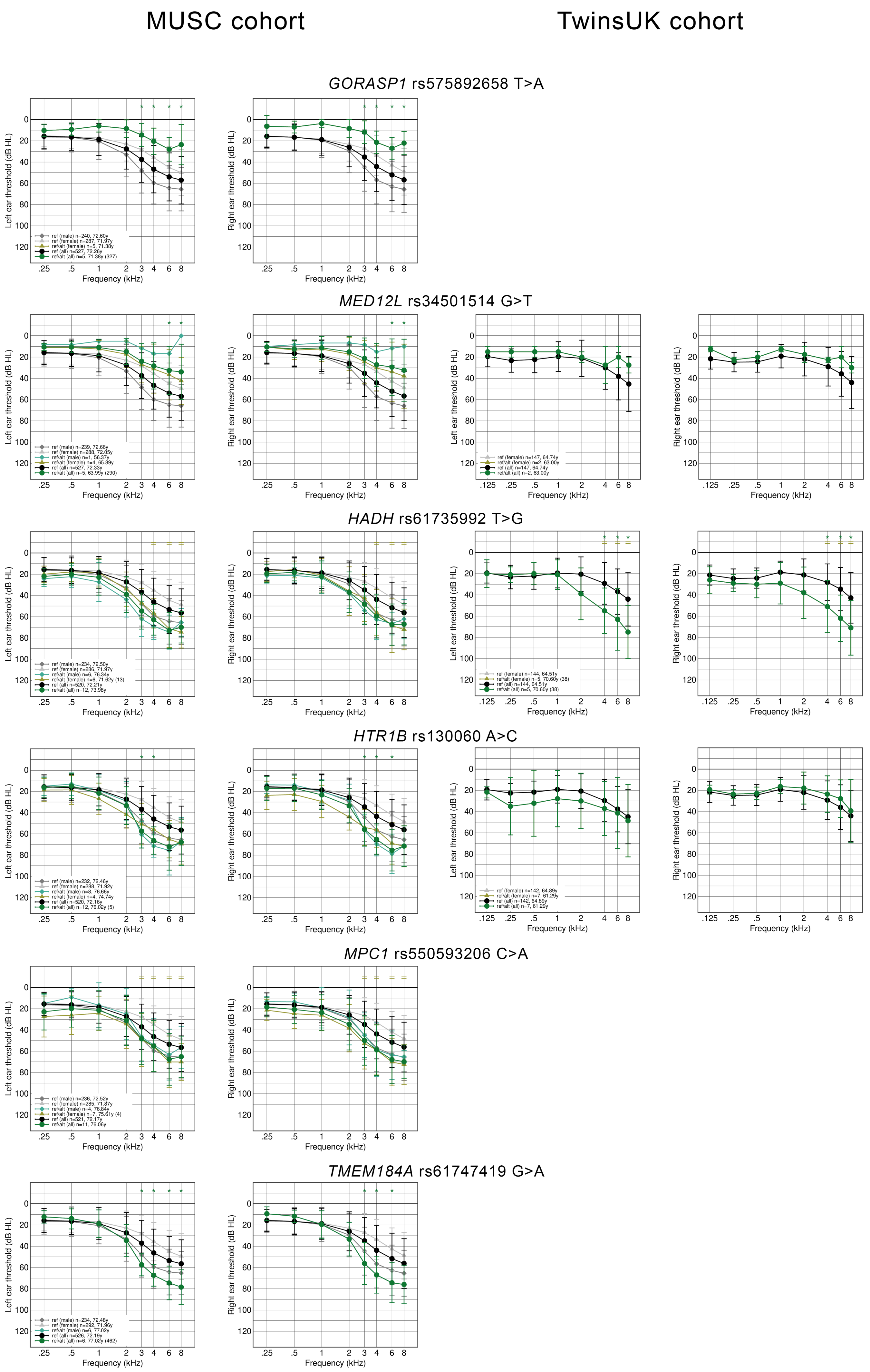
MUSC cohort
TwinsUK cohort
GORASP1 rs575892658 T>A
MED12L rs34501514 G>T
HADH rs61735992 T>G
HTR1B rs130060 A>C
MPC1 rs550593206 C>A
TMEM184A rs61747419 G>A
Left ear threshold (dB HL)
Right ear threshold (dB HL)
Frequency (kHz)

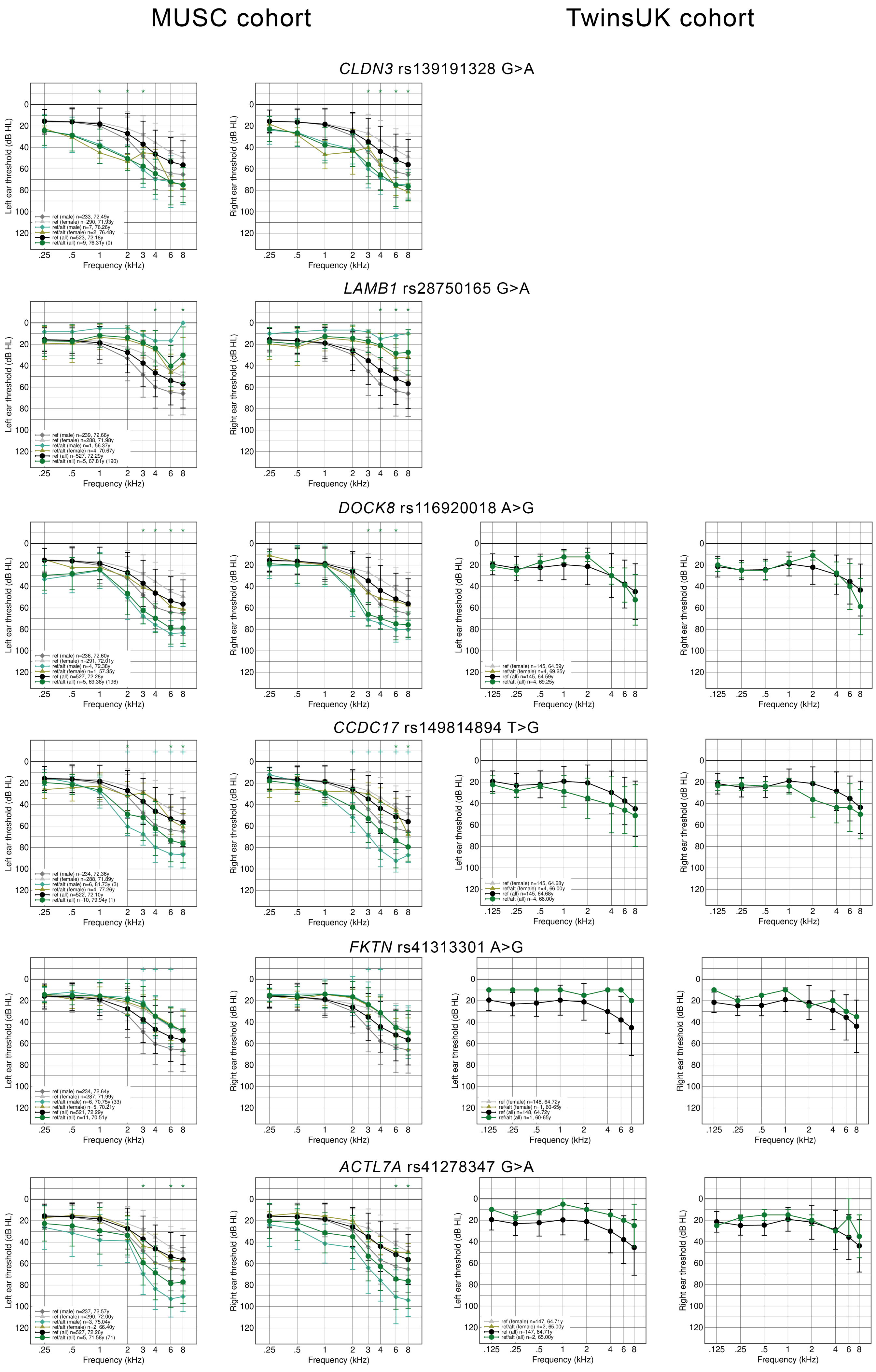

MUSC cohort
TwinsUK cohort
CLDN3 rs139191328 G>A
LAMB1 rs28750165 G>A
DOCK8 rs116920018 A>G
CCDC17 rs149814894 T>G
FKTN rs41313301 A>G
ACTL7A rs41278347 G>A
Left ear threshold (dB HL)
Right ear threshold (dB HL)
Frequency (kHz)

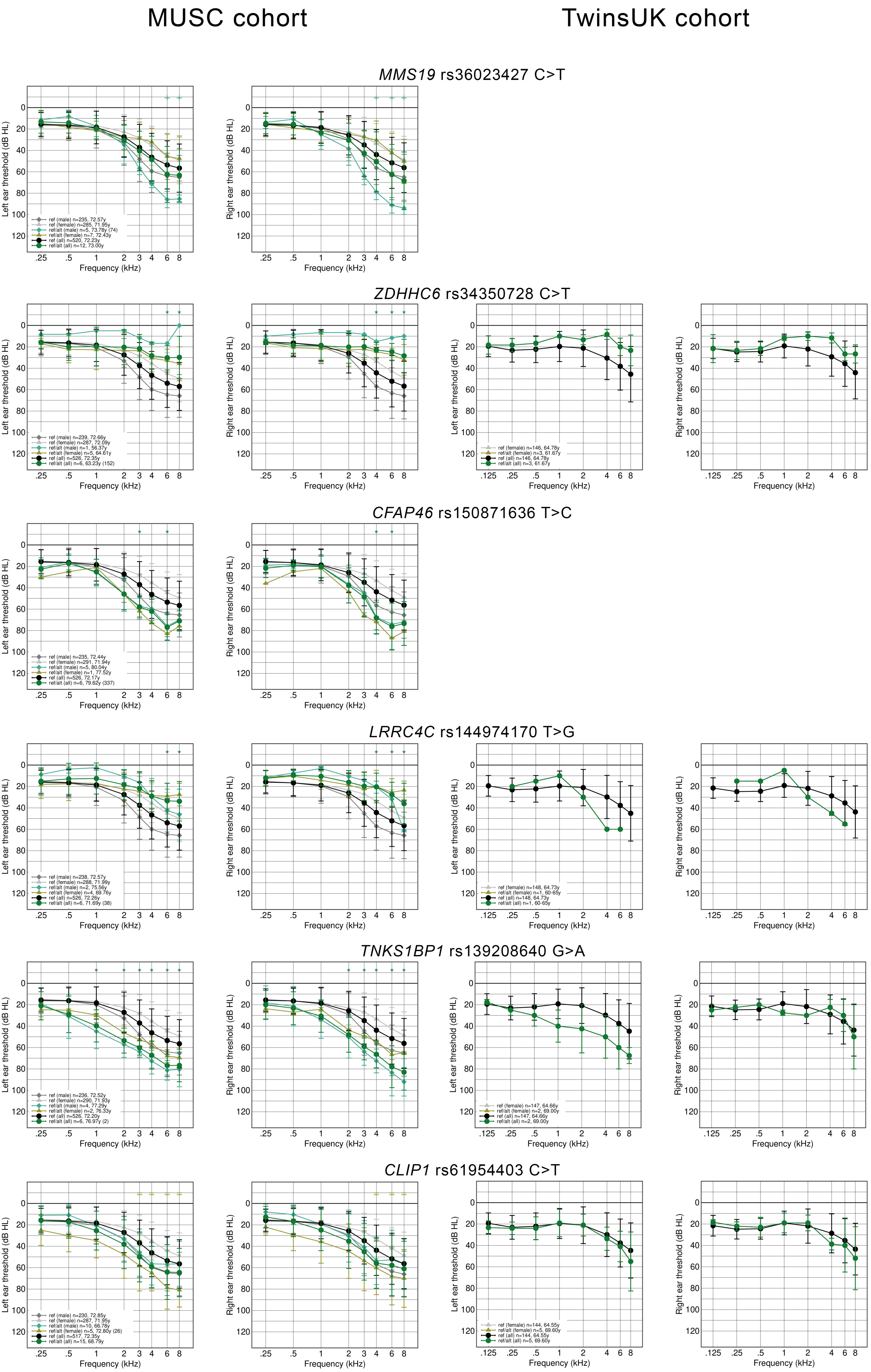

MUSC cohort
TwinsUK cohort
MMS19 rs36023427 C>T
ref (male) n=235, 72.57y
ref (female) n=285, 71.95y
ref/alt (male) n=5, 73.78y (74)
ref/alt (female) n=7, 72.43y
ref (all) n=520, 72.23y
ref/alt (all) n=12, 73.00y
ZDHHC6 rs34350728 C>T
ref (male) n=239, 72.66y
ref (female) n=287, 72.09y
ref/alt (male) n=1, 56.37y
ref/alt (female) n=5, 64.61y
ref (all) n=526, 72.35y
ref/alt (all) n=6, 63.23y (152)
ref (female) n=146, 64.78y
ref/alt (female) n=3, 61.67y
ref (all) n=146, 64.78y
ref/alt (all) n=3, 61.67y
CFAP46 rs150871636 T>C
ref (male) n=235, 72.44y
ref (female) n=291, 71.94y
ref/alt (male) n=5, 80.04y
ref/alt (female) n=1, 77.52y
ref (all) n=526, 72.17y
ref/alt (all) n=6, 79.62y (337)
LRRC4C rs144974170 T>G
ref (male) n=238, 72.57y
ref (female) n=288, 71.99y
ref/alt (male) n=2, 75.56y
ref/alt (female) n=4, 69.76y
ref (all) n=526, 72.26y
ref/alt (all) n=6, 71.69y (38)
ref (female) n=148, 64.73y
ref/alt (female) n=1, 60-65y
ref (all) n=148, 64.73y
ref/alt (all) n=1, 60-65y
TNKS1BP1 rs139208640 G>A
ref (male) n=236, 72.52y
ref (female) n=290, 71.93y
ref/alt (male) n=4, 77.29y
ref/alt (female) n=2, 76.33y
ref (all) n=526, 72.20y
ref/alt (all) n=6, 76.97y (2)
ref (female) n=147, 64.66y
ref/alt (female) n=2, 69.00y
ref (all) n=147, 64.66y
ref/alt (all) n=2, 69.00y
CLIP1 rs61954403 C>T
ref (male) n=230, 72.85y
ref (female) n=287, 71.95y
ref/alt (male) n=10, 66.78y
ref/alt (female) n=5, 72.80y (26)
ref (all) n=517, 72.35y
ref/alt (all) n=15, 68.79y
ref (female) n=144, 64.55y
ref/alt (female) n=5, 69.60y
ref (all) n=144, 64.55y
ref/alt (all) n=5, 69.60y
Left ear threshold (dB HL)
Right ear threshold (dB HL)
Frequency (kHz)

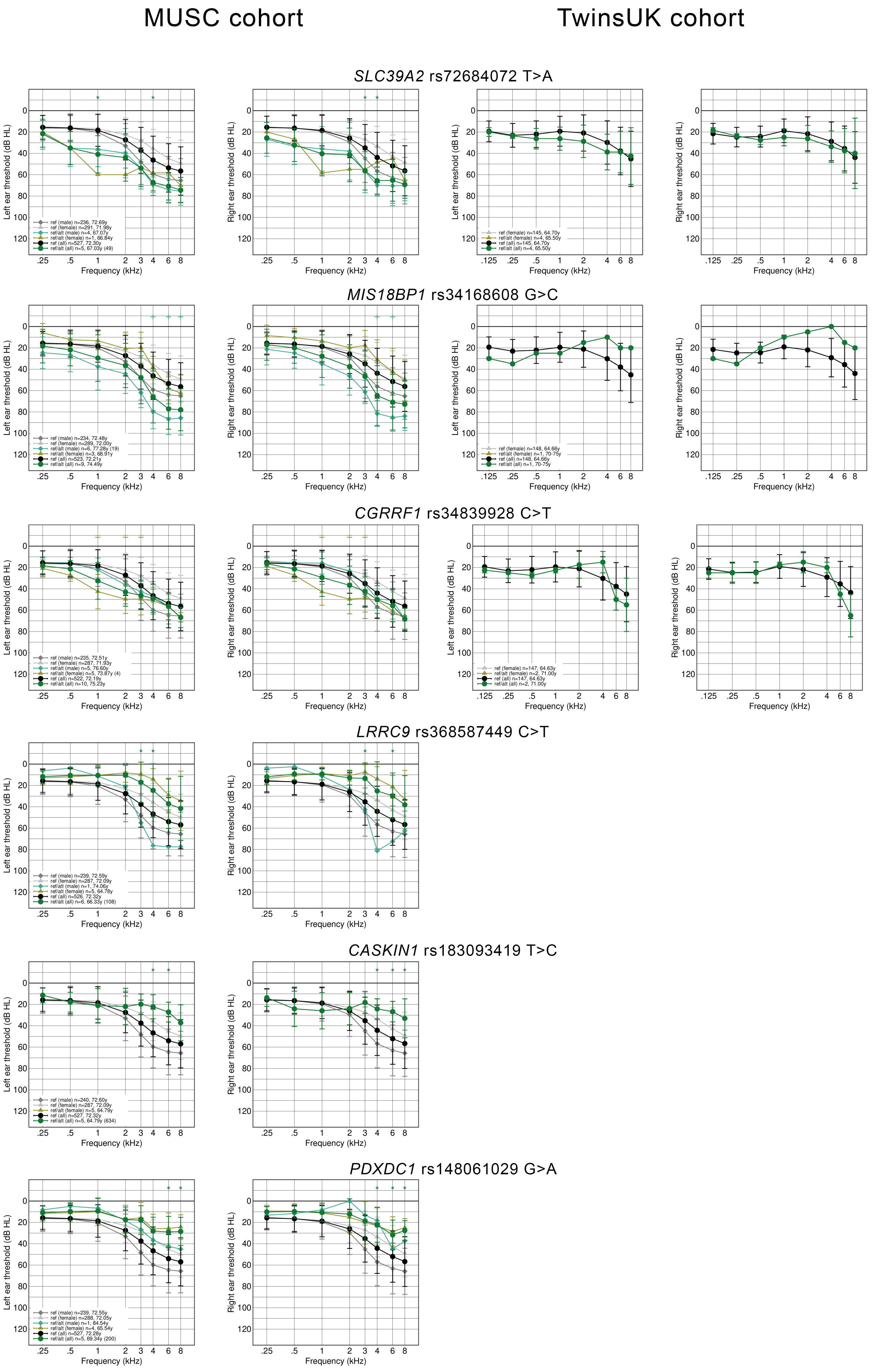
MUSC cohort
TwinsUK cohort
SLC39A2 rs72684072 T>A
MIS18BP1 rs34168608 G>C
CGRRF1 rs34839928 C>T
LRRC9 rs368587449 C>T
CASKIN1 rs183093419 T>C
PDXDC1 rs148061029 G>A
Left ear threshold (dB HL)
Right ear threshold (dB HL)
Frequency (kHz)

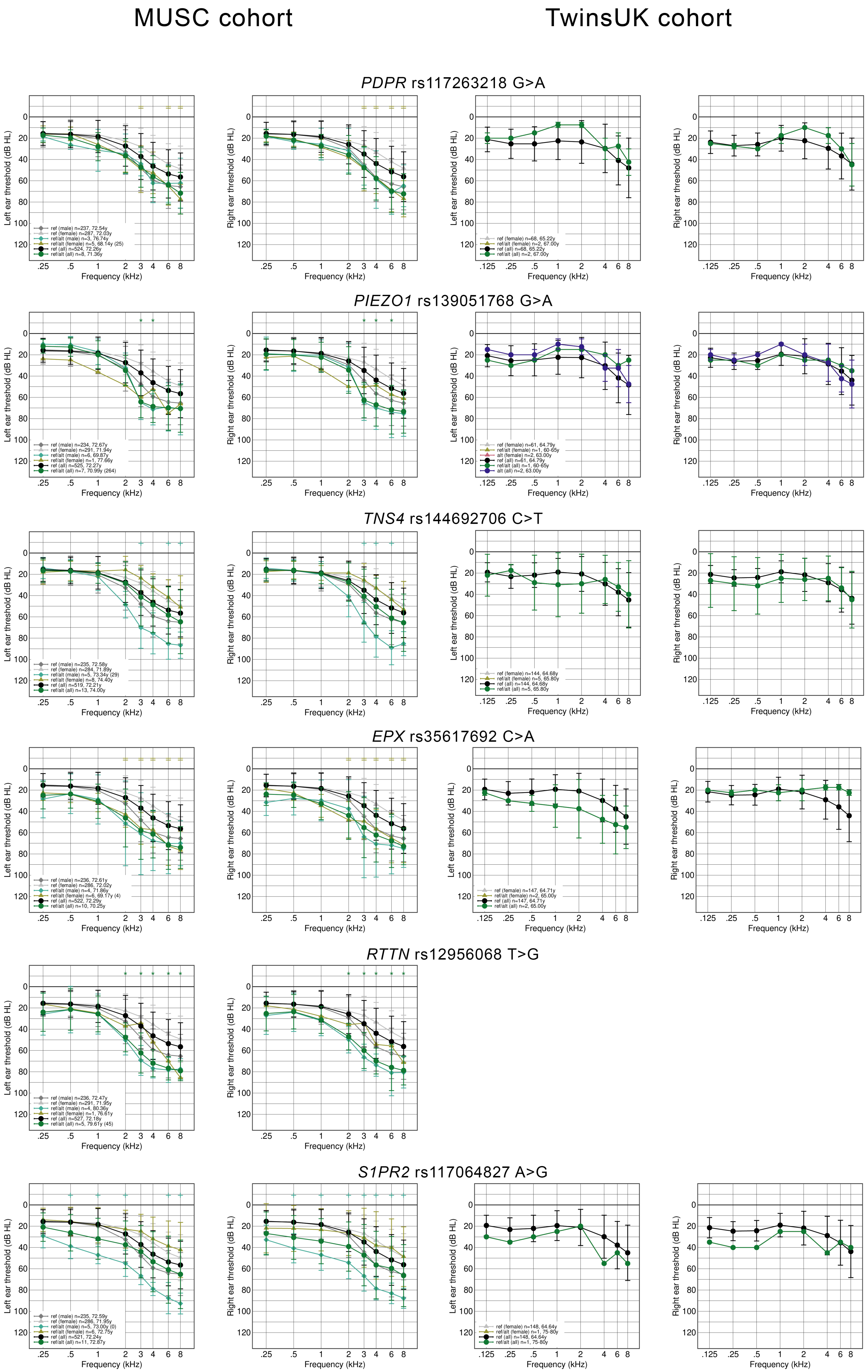

MUSC cohort
TwinsUK cohort
PDPR rs117263218 G>A
PIEZO1 rs139051768 G>A
TNS4 rs144692706 C>T
EPX rs35617692 C>A
RTTN rs12956068 T>G
S1PR2 rs117064827 A>G
Left ear threshold (dB HL)
Right ear threshold (dB HL)
Frequency (kHz)

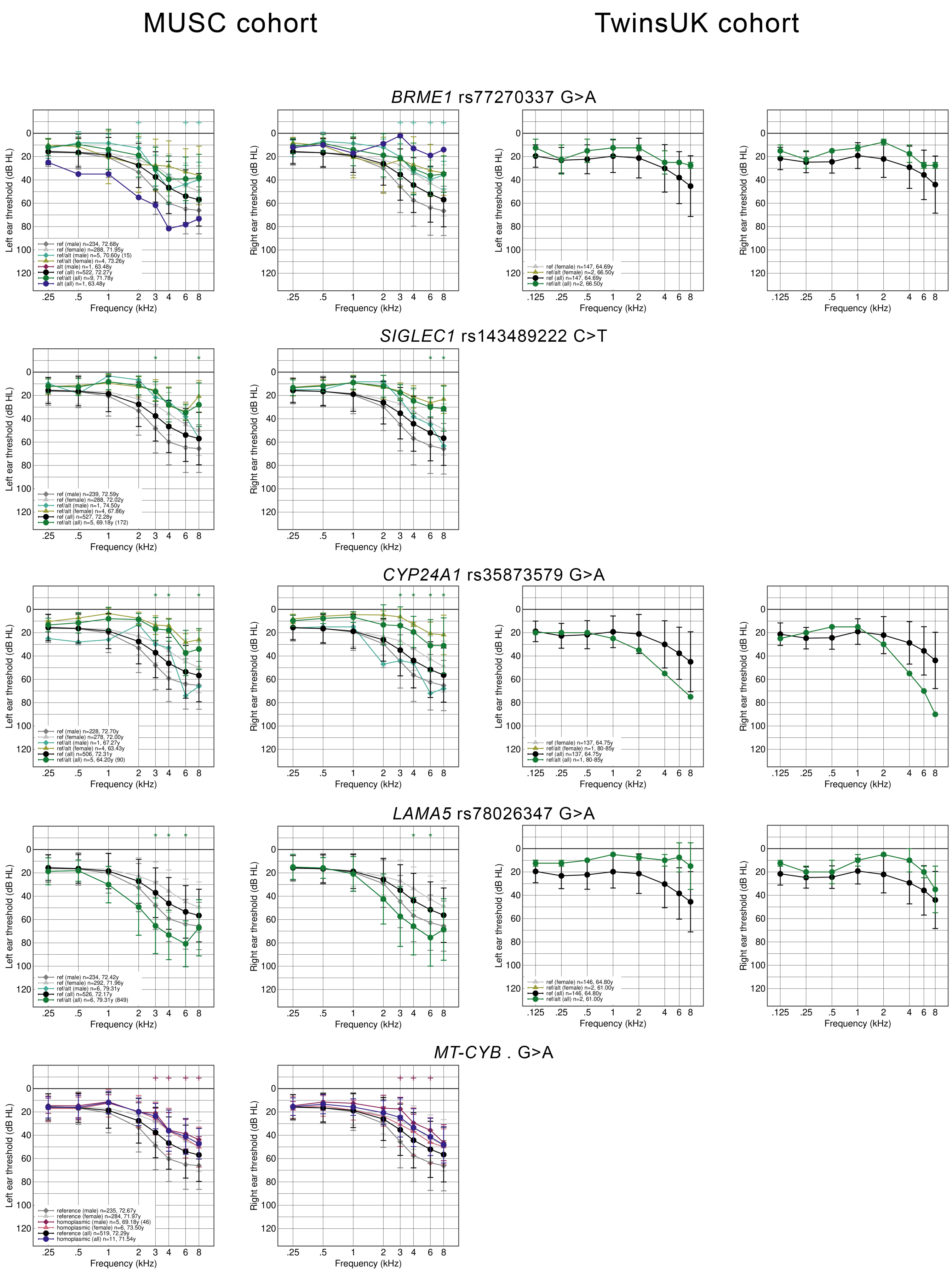
MUSC cohort
TwinsUK cohort
BRME1 rs77270337 G>A
ref (male) n=234, 72.68y
ref (female) n=288, 71.95y
ref/alt (male) n=5, 70.60y (15)
ref/alt (female) n=4, 73.26y
alt (male) n=1, 63.48y
ref (all) n=522, 72.27y
ref/alt (all) n=9, 71.78y
alt (all) n=1, 63.48y
ref (female) n=147, 64.69y
ref/alt (female) n=2, 66.50y
ref (all) n=147, 64.69y
ref/alt (all) n=2, 66.50y
SIGLEC1 rs143489222 C>T
ref (male) n=239, 72.59y
ref (female) n=288, 72.02y
ref/alt (male) n=1, 74.50y
ref/alt (female) n=4, 67.86y
ref (all) n=527, 72.28y
ref/alt (all) n=5, 69.18y (172)
CYP24A1 rs35873579 G>A
ref (male) n=228, 72.70y
ref (female) n=278, 72.00y
ref/alt (male) n=1, 67.27y
ref/alt (female) n=4, 63.43y
ref (all) n=506, 72.31y
ref/alt (all) n=5, 64.20y (90)
ref (female) n=137, 64.75y
ref/alt (female) n=1, 80-85y
ref (all) n=137, 64.75y
ref/alt (all) n=1, 80-85y
LAMA5 rs78026347 G>A
ref (male) n=234, 72.42y
ref (female) n=292, 71.96y
ref/alt (male) n=6, 79.31y
ref (all) n=526, 72.17y
ref/alt (all) n=6, 79.31y (849)
ref (female) n=146, 64.80y
ref/alt (female) n=2, 61.00y
ref (all) n=146, 64.80y
ref/alt (all) n=2, 61.00y
MT-CYB . G>A
reference (male) n=235, 72.67y
reference (female) n=284, 71.97y
homoplasmic (male) n=5, 69.18y (46)
homoplasmic (female) n=6, 73.50y
reference (all) n=519, 72.29y
homoplasmic (all) n=11, 71.54y
Left ear threshold (dB HL)
Right ear threshold (dB HL)
Frequency (kHz)

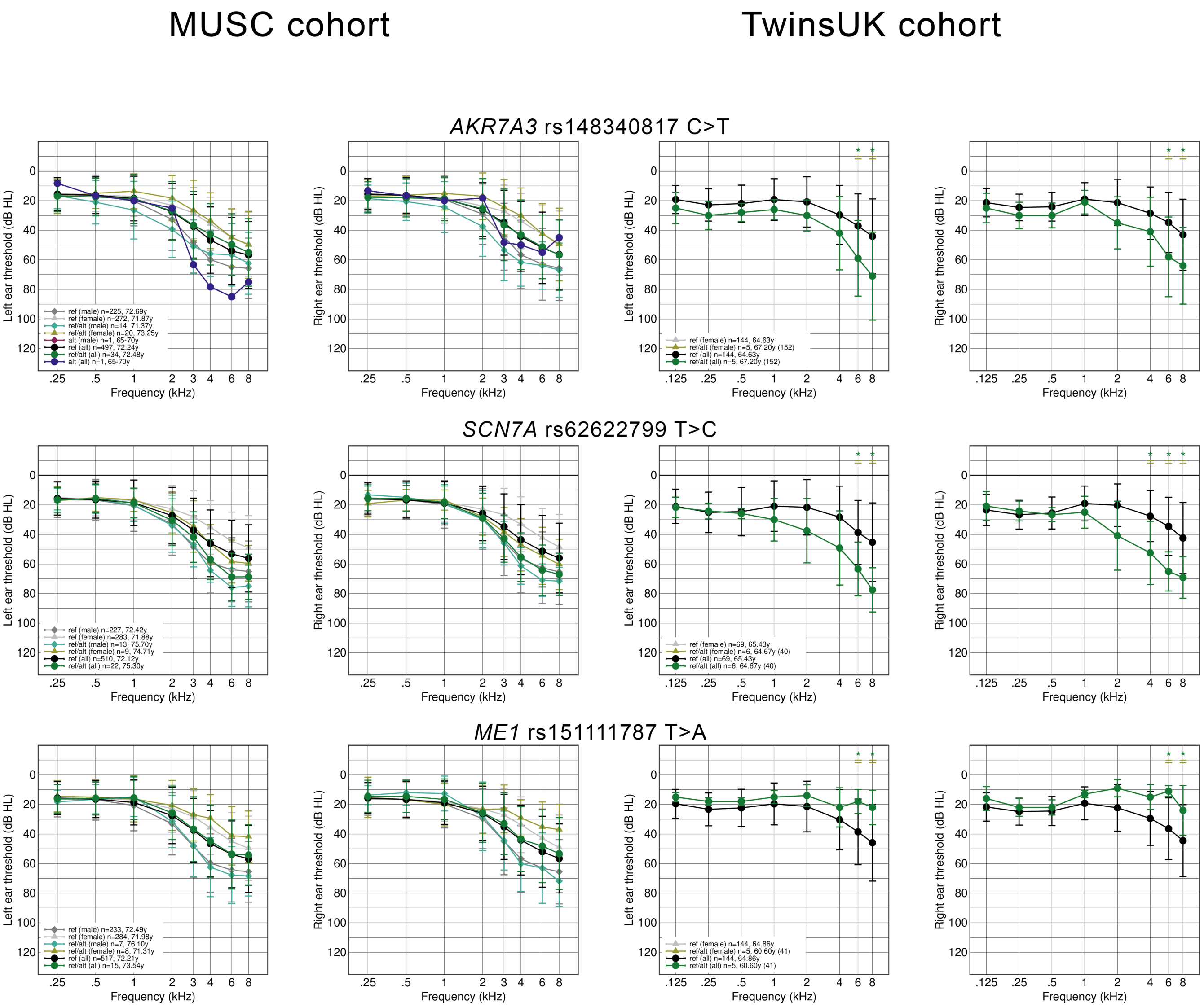
MUSC cohort
TwinsUK cohort
AKR7A3 rs148340817 C>T
SCN7A rs62622799 T>C
ME1 rs151111787 T>A
Left ear threshold (dB HL)
Right ear threshold (dB HL)
Frequency (kHz)
ref (male) n=225, 72.69y
ref (female) n=272, 71.87y
ref/alt (male) n=14, 71.37y
ref/alt (female) n=20, 73.25y
alt (male) n=1, 65-70y
ref (all) n=497, 72.24y
ref/alt (all) n=34, 72.48y
alt (all) n=1, 65-70y
ref (female) n=144, 64.63y
ref/alt (female) n=5, 67.20y (152)
ref (all) n=144, 64.63y
ref/alt (all) n=5, 67.20y (152)
ref (male) n=227, 72.42y
ref (female) n=283, 71.88y
ref/alt (male) n=13, 75.70y
ref/alt (female) n=9, 74.71y
ref (all) n=510, 72.12y
ref/alt (all) n=22, 75.30y
ref (female) n=69, 65.43y
ref/alt (female) n=6, 64.67y (40)
ref (all) n=69, 65.43y
ref/alt (all) n=6, 64.67y (40)
ref (male) n=233, 72.49y
ref (female) n=284, 71.98y
ref/alt (male) n=7, 76.10y
ref/alt (female) n=8, 71.31y
ref (all) n=517, 72.21y
ref/alt (all) n=15, 73.54y
ref (female) n=144, 64.86y
ref/alt (female) n=5, 60.60y (41)
ref (all) n=144, 64.86y
ref/alt (all) n=5, 60.60y (41)

**Suppl. Figure 7**. Audiograms from the MUSC cohort (left pair) and TwinsUK cohort (right pair) showing threshold differences for carriers of the 41 variants identified in the MUSC cohort and the 4 found in the TwinsUK cohort. The corresponding audiogram from the other cohort is shown where there were carriers of that variant. The numbers in brackets in the keys show the number of permutations giving a similar result for that group. Two audiograms are shown for each variant in each cohort; the thresholds from the left ear are shown on the left, and those from the right ear on the right.
